# Interventions for Cognitive Impairment in Type 2 Diabetes Mellitus: An Umbrella Review of Pharmacological and Lifestyle Strategies, Mechanisms, and Clinical Implications

**DOI:** 10.64898/2026.10.23.26366288

**Authors:** Herbert Izo Ninsiima, Eze Ejike Daniel, Irene Nakito Kisakye, Immaculate Mandera, Herbert Elvis Ainamani, Mercy Akambasisa, Geofrey Ayebazibwe, Daniel Matovu

## Abstract

**Background:** Type 2 diabetes mellitus (T2DM) is one of the fastest-growing epidemics associated with cognitive impairment and dementia. This reduces quality of life among patients with diabetes. Several trials on the management of cognitive impairment have been done although their effectiveness and mechanisms are still unclear.

**Aim:** To synthesize evidence from systematic reviews and meta-analyses of RCTs examining interventions for cognitive impairment in T2DM, stratified by intervention category, comparator, patient characteristics, mechanisms and clinical outcomes.

**Methods:** We identified eleven systematic reviews (n = 213,604 participants) reporting effects of cognitive impairment interventions through comprehensive literature search. Using random effect models, a meta-meta-analysis was conducted and heterogeneity assessed using I² statistics. AMSTAR and GRADE were used to assess for quality of reviews. Subgroup analyses were conducted for intervention category, cognitive severity, age, diabetes duration, sex, and follow-up duration.

**Result:** Sodium glucose co-transporter inhibitors showed the strongest cognitive improvement effect (pooled OR = 0.44, 95% CI: 0.35–0.55; SUCRA = 94.5%), followed by GLP-1 receptor agonists (RR/OR = 0.60, 95% CI: 0.50–0.72; SUCRA = 92.1%). Among lifestyle interventions, multi-component exercise was the most effective (SMD = 0.69–1.04). The lower baseline cognition (MMSE < 23) benefited more from GLP-1 receptor agonists (Cohen’s d = -3.00, p < 0.001). Advanced age (≥60 years) and prolonged intervention time (≥24 weeks for medications; ≥6 months for exercise) were associated with greater effects. Female gender showed greater benefit from GLP-1 receptor agonists (HR = 0.63 vs 0.81 for male gender).

**Conclusions:** SGLT2 inhibitors and GLP-1 receptor agonists have the greatest effect on cognition in T2DM, and the multi-component exercise as the most effective lifestyle component. The review therefore recommends individualized intervention selection based on patient characteristics and baseline cognitive status. This calls for high quality trials with prolonged follow-up to strengthen the evidence base.

**PROSPERO Registration:** The trial was registered in PROSPERO under number CRD420261453178

## Introduction

Type 2 Diabetes Mellitus (T2DM) is a major global public health problem affecting over 500 million people worldwide (Duncan et al., 2021). Beyond its well-recognized microvascular and macrovascular complications, increasing evidence suggests that T2DM is associated with accelerated cognitive decline, mild cognitive impairment, and dementia (Bao et al., 2025). Individuals with T2DM have an estimated 1.5–2 times greater risk of developing cognitive impairment compared to non-diabetic populations (Biessels et al., 2006).

Several mechanisms have been proposed to explain diabetes-associated cognitive dysfunction, including chronic hyperglycemia, insulin resistance, neuroinflammation, oxidative stress, endothelial dysfunction, cerebral microvascular disease, mitochondrial dysfunction, and neurodegenerative processes such as amyloid-β accumulation and tau hyperphosphorylation (Bao et al., 2025; Biessels et al., 2006). Consequently, numerous pharmacological and non-pharmacological interventions have been investigated to improve cognitive outcomes among individuals with T2DM (Allen et al., 2013; An et al., 2025; Gupta et al., 2011)

These interventions include antidiabetic medications (e.g., metformin, GLP-1 receptor agonists, SGLT2 inhibitors) (Khan et al., 2026; Osman et al., 2025; Youssef et al., 2023) nutritional supplements (e.g., zinc, omega-3 fatty acids, vitamins) (Ghaedi et al., 2024; Jafarnejad et al., 2019)physical exercise, dietary modification, cognitive training, and multidomain lifestyle interventions (Margara-Escudero et al., 2025; Marlon E Cerf, 2021; Omolade Komolafe & Anthonia Okafor, 2024). While multiple systematic reviews and meta-analyses have synthesized evidence from randomized controlled trials (RCTs), findings remain fragmented and vary in methodological quality.

An umbrella review is therefore needed to comprehensively evaluate and compare evidence from existing systematic reviews and meta-analyses. It is also important in examining the biological mechanisms through which interventions may improve cognitive outcomes in adults with T2DM.

### Rationale

Type 2 Diabetes Mellitus (T2DM)-induced cognitive impairment is a recognized complication among T2DM patients. Cognitive impairment adversely affects diabetic self-care, treatment adherence, quality of life, and the risk of dementia. Although numerous randomized controlled trials have evaluated pharmacological and non-pharmacological interventions to improve cognitive function, the evidence has been synthesized in multiple systematic reviews with varying scope, quality, and conclusions. This fragmentation makes it difficult to identify the most effective interventions and their overall strength of evidence.

Most existing systematic reviews focus on individual interventions, such as antidiabetic drugs, nutritional supplements, exercise, or lifestyle modification, rather than comparing evidence across intervention types. In addition, relatively few reviews comprehensively examine the biological mechanisms through which these interventions influence cognition, including improvements in glycemic control, insulin sensitivity, oxidative stress, neuroinflammation, and vascular function. A broader synthesis is therefore needed to integrate both clinical effectiveness and mechanistic evidence.

An umbrella review offers the highest level of evidence synthesis by integrating findings from systematic reviews and meta-analyses of randomized controlled trials. It enables comparison of intervention effectiveness, assessment of the quality and consistency of evidence, and identification of research gaps. The findings will inform clinical practice, guide future research, and support the development of evidence-based strategies to prevent or reduce cognitive impairment among adults with T2DM.

### Review Question

What is the effectiveness of interventions evaluated in randomized controlled trials for improving cognitive impairment in adults with Type 2 Diabetes Mellitus, and through which biological mechanisms do these interventions exert their effects?

### Objectives

#### Primary Objective

To synthesize evidence from systematic reviews and meta-analyses of randomized controlled trials evaluating interventions aimed at improving cognitive impairment among adults with Type 2 Diabetes Mellitus.

### Secondary Objectives

1. To compare the effectiveness of pharmacological and non-pharmacological interventions on cognitive outcomes among adults with T2DM, stratified by intervention category, comparator, and patient characteristics.
2. To assess the quality and strength of evidence supporting these interventions.
3. To identify and summarize the biological mechanisms through which these interventions improve cognitive function in individuals with T2DM.

## Methods

### Study Design

Umbrella review of systematic reviews and meta-analyses of randomized controlled trials.

The review was conducted according to the recommendations of the Joanna Briggs Institute (JBI) Manual for Evidence Synthesis and reported following the Preferred Reporting Items for Overviews of Reviews (PRIOR) statement (Pollock et al., 2019).

### Eligibility Criteria

This umbrella review reviewed systematic reviews and meta-analyses (including network meta-analyses) that contain randomized controlled trials. The review focused on adults (≥18 years) diagnosed with Type 2 Diabetes Mellitus with any form of cognitive decline, including mild cognitive impairment, subjective cognitive decline, diabetes-associated cognitive dysfunction, or early dementia.

Only reviews that examined any intervention intended to improve cognitive function, including pharmacological agents (metformin, GLP-1 receptor agonists, SGLT2 inhibitors, DPP-4 inhibitors, thiazolidinediones, insulin-based interventions, and neuroprotective agents). The review also included those that assessed the nutritional interventions (zinc supplementation, vitamins, omega-3 fatty acids, antioxidants, and micronutrient supplementation). The other interventions considered were, lifestyle interventions (physical exercise, dietary interventions, weight reduction programs, and multidomain lifestyle programs), as well as cognitive and behavioral interventions (cognitive training, memory rehabilitation, and digital cognitive therapies).

Eligible comparators included placebo, standard of care, no intervention, or alternative interventions. The primary outcomes of interest measures were cognitive performance assessed using validated instruments such as the Mini-Mental State Examination (MMSE), Montreal Cognitive Assessment (MoCA), and Alzheimer’s Disease Assessment Scale-Cognitive Subscale (ADAS-Cog). The secondary outcomes included mechanisms including glycemic control (HbA1c and fasting glucose), insulin sensitivity, inflammatory biomarkers, oxidative stress biomarkers, neurotrophic factors, neuroimaging outcomes, and cerebral blood flow. Only Systematic reviews, meta-analyses, and network meta-analyses containing randomized controlled trials were included. We excluded narrative reviews, scoping reviews, editorials, commentaries, protocols, and reviews containing only observational studies.

The following databases were searched: MEDLINE (PubMed), Embase, Scopus and Web of Science.

### Search Strategy

A combination of controlled vocabulary and keywords used in study was: (“Type 2 Diabetes Mellitus” OR T2DM OR diabetes) AND (cognitive impairment OR cognitive decline OR cognition OR dementia OR memory) AND (systematic review OR meta-analysis) AND (randomized controlled trial OR RCT) search strategies were adapted for each database.

### Selection Process

All records were imported into Rayyan and duplicates removed. Two independent reviewers screened the titles, abstracts and full texts. The disagreements were resolved by through discussion and consultation with a third reviewer. The study selection process is presented using PRISMA flow diagram (figure 1).

**Figure 1:**
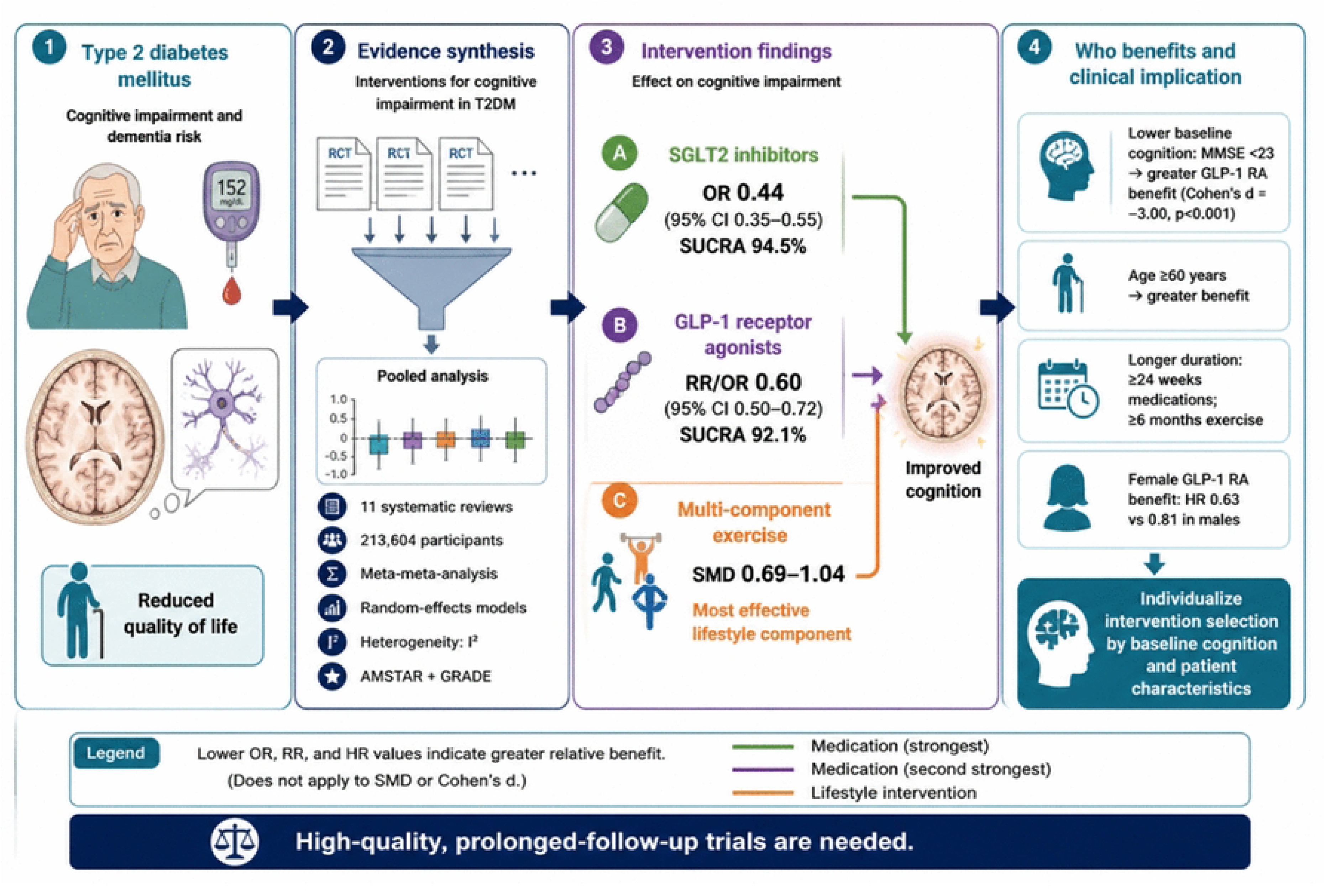
Graphical abstract.

### Data Collection

The data extraction process was guided by a standardized form developed specifically for this purpose. Two (NIH, GA) independent reviewers extracted all relevant information from each included systematic review. The extracted included bibliographic information (author, year, and country), the type of review, the number of randomized controlled trials (RCTs) included, and the total sample size. Furthermore, comprehensive descriptive data on population characteristics, the nature of the intervention and its comparator, as well as the specific cognitive outcomes measured were extracted. To facilitate a robust synthesis, we also captured quantitative data such as effect sizes, alongside qualitative insights like mechanistic findings, the conclusions drawn by the review authors, and the results of their methodological quality assessment.

### Data items

The primary outcome for which data were sought was cognitive performance assessed using validated instruments such as MMSE, MoCA, and ADAS-Cog. Secondary outcomes for which data were sought included mechanisms (glycemic control, insulin sensitivity, inflammatory biomarkers, oxidative stress biomarkers, neurotrophic factors, neuroimaging outcomes, and cerebral blood flow). All results compatible with each outcome domain in each study were sought, including all measures and time points reported.

Other variable included: bibliographic information (author, year, country), type of review, number of RCTs included, total sample size, population characteristics, comparator, cognitive outcomes measured, effect size, mechanistic findings, conclusions, and methodological quality assessment results. Where information was missing or unclear, this was noted and assumptions were documented.

### Study Risk of Bias Assessment

Methodological quality of the included reviews was appraised using the AMSTAR 2 (A Measurement Tool to Assess Systematic Reviews 2) to evaluate the rigor by two (NIH, GA) reviewers. Discrepancies were resolved by consensus with a third reviewer. This was used to categorize each review into one of four confidence ratings: high, moderate, low, or critically low. At the same time, the certainty of the evidence for the key findings was assessed. Where the included reviews had already reported risk of bias assessments (Cochrane RoB, RoB 2, NOS, PEDrp, JBI) these were extracted directly. Where such assessments were not reported, the certainty of the evidence was independently evaluated by the review team using established GRADE principles.

### Effect measures

For each outcome, the effect measures used in the synthesis or presentation of results were as follows: dementia incidence and cognitive decline were expressed as risk ratios (RR) or odds ratios (OR) with 95% confidence intervals; cognitive performance (MMSE, MoCA, ADAS-Cog, and domain-specific tests) was expressed as standardized mean differences (SMD) or mean differences (MD) with 95% confidence intervals; and mechanistic outcomes (HbA1c, fasting glucose, inflammatory biomarkers, etc.) were expressed as MD or SMD as reported in the included reviews.

### Synthesis methods

This umbrella review employed a narrative synthesis approach as the primary method of evidence synthesis, given the anticipated heterogeneity in interventions, outcomes, and populations. Where included reviews reported comparable outcomes and comparators, we additionally performed exploratory second-order meta-analysis (meta-meta-analysis) to generate pooled effect estimates. This approach was exploratory and intended to summarize the range of effect estimates rather than to provide definitive pooled estimates. When multiple reviews reported the same the same comparison, we selected the review with the highest AMSTAR 2 rating or, where equal, the most comprehensive review (largest number of primary studies).

Overlap among primary studies included in the 11 reviews was assessed using the Corrected Covered Area (CCA) method. The CCA was calculated as follows: CCA = (N − r) / (r × c − r), where N is the total number of primary study occurrences across all reviews, r is the number of primary studies, and c is the number of reviews. The calculated CCA was 2.11%, indicating slight overlap. For the exploratory meta-meta-analysis, we used a hierarchical random-effects model to account for within-review and between-review variance. When multiple reviews reported the same comparison, we selected the review with the highest AMSTAR 2 rating or, where equal, the most comprehensive review (largest number of primary studies).

Results of individual studies and syntheses were tabulated in tables 1–8 and displayed using forest plots and PRISMA flow diagram (Figure 2). A graphical abstract (Figure 1) and mechanistic pathway diagram (Figure 3) were also produced.

**FIGURE 2:**
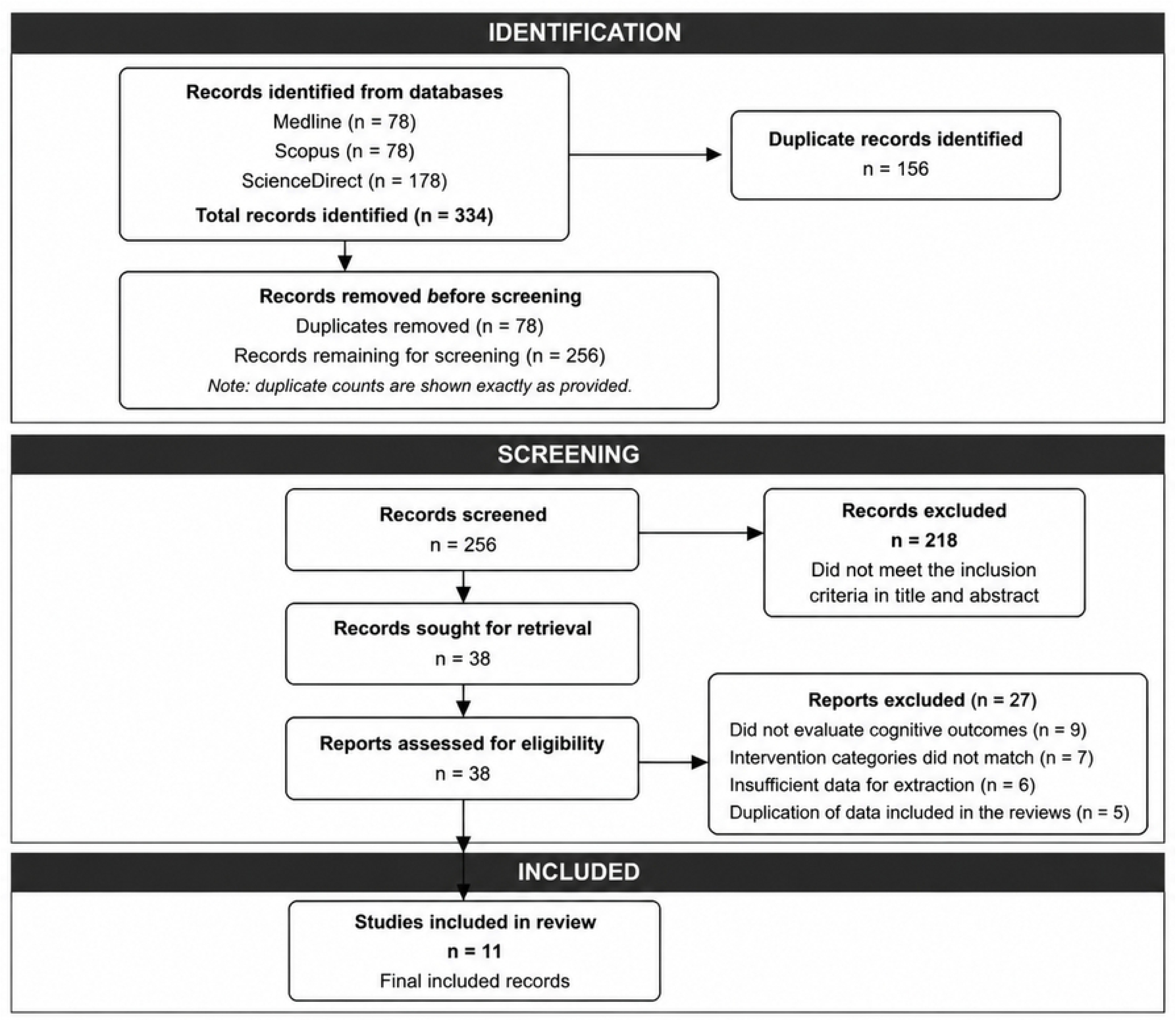
Preferred Reporting Items for Overviews of Reviews (PRISMA) flow chart.

**Fig 3:**
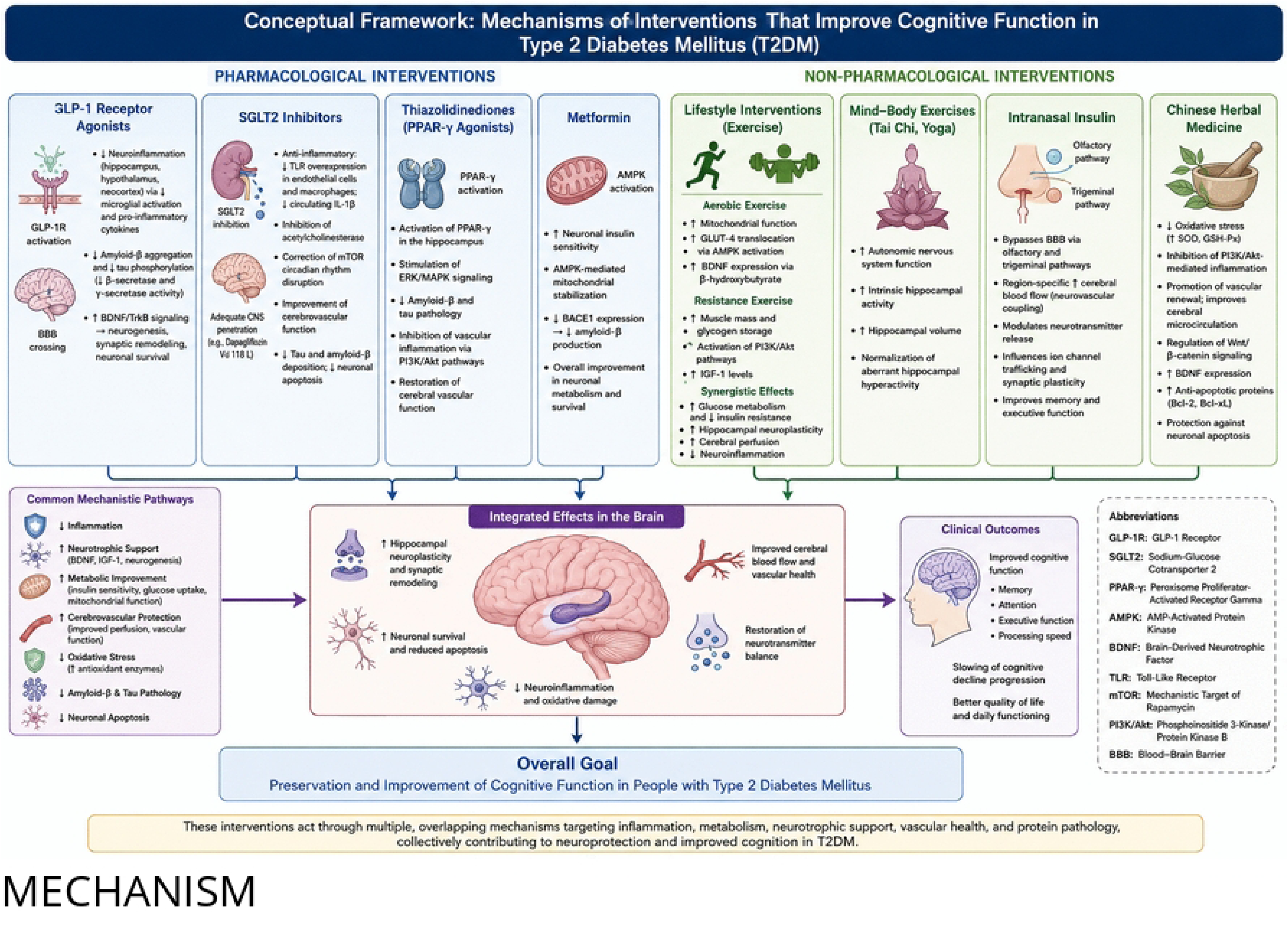
Mechanistic pathways.

For the exploratory meta-meta-analysis, random-effects models were fitted using the meta for package (version 4.0-1) in R (version 4.3.2). Restricted maximum likelihood (REML) was used to estimate between-study variance (τ²). Heterogeneity was quantified using the I² statistic and Cochran’s Q test. Subgroup analyses were conducted using mixed-effects models. SUCRA rankings were derived from network meta-analytic estimates reported in the original reviews; where SUCRA values were not reported, we did not generate them de novo.

Subgroup analyses were conducted using mixed-effect models. They were performed for intervention category, cognitive severity, age, diabetes duration, sex, and follow-up duration.

Sensitivity analyses were conducted as reported in the reviews to assess the robustness of the synthesized results. Where sensitivity analyses were reported by the original authors, these were extracted and summarized.

Risk of bias due to missing results in the synthesis (arising from reporting biases) was assessed. Where included reviews reported publication bias assessments (funnel plots, Egger’s test, Beggs test), these were extracted directly. The presence of publication bias was noted for each intervention category where reported.

The certainty of the evidence for the key findings was assessed. Where the included reviews had already reported GRADE assessments, these were extracted directly. Where such assessments were not reported, the certainty of evidence was independently evaluated by the review team using established GRADE principles. Two reviewers (HN and GA) independently applied GRADE principles; discrepancies were resolved by consensus with a third reviewer. Ratings ranged from very low certainty to high certainty across outcomes and reviews. Certainty was downgraded for risk of bias, inconsistency, indirectness, imprecision, and publication bias.

## 3.0 Results

The PRISMA flow chart (fig 2) illustrates the systematic literature search and screening process for this umbrella review. A total of 334 records were initially identified through database searching, consisting 78 records from Medline, 78 from Scopus, and 178 from Science Direct. After the removal of duplicate records (n = 158), 256 unique records were screened based on their titles and abstracts. In the initial screening phase, 218 records were excluded as they did not meet the predefined eligibility criteria, leaving 38 articles for full-text screening. Of these, 27 articles were excluded for the following reasons: studies did not specifically evaluate cognitive outcomes in T2DM populations (n = 9), intervention categories did not match the predefined framework (n = 7), insufficient data for extraction (n = 6), or duplication of data from included systematic reviews (n = 5). Finally, 11 systematic reviews met all inclusion criteria and were included in the data extraction and final qualitative synthesis of this umbrella review. This high attrition rate during full text screening reflects the application of rigorous methodological criteria to ensure that only high-quality, relevant evidence was synthesized across the five intervention categories examined in this umbrella review.

### Characteristics of included studies

Table 1 presents the characteristics of the 11 systematic reviews and meta-analyses included in this umbrella review. The systematic reviews and metanalyses were published between 2022 and 2026 and included a total of over 210,000 participants across in 279 RCTs. Five intervention categories: antidiabetic medications (n = 4 reviews), nutritional supplements (n = 1), lifestyle interventions including exercise and intranasal insulin (n = 5), cognitive interventions (n = 0), and multidomain interventions (n = 0) analyzed. The most comprehensive reviews were those investigating antidiabetic medications, with (Z. Li, Lin, et al., 2024) including over 3.4 million participants from 41 observational studies and 23 RCTs. The other review was from (E. Hui et al., 2025). synthesizing evidence from 42 studies across multiple drug classes. The largest sample sizes were in pharmacological reviews, while exercise interventions typically included smaller samples ranging from 884 to 3,201 participants.

**Table one:**
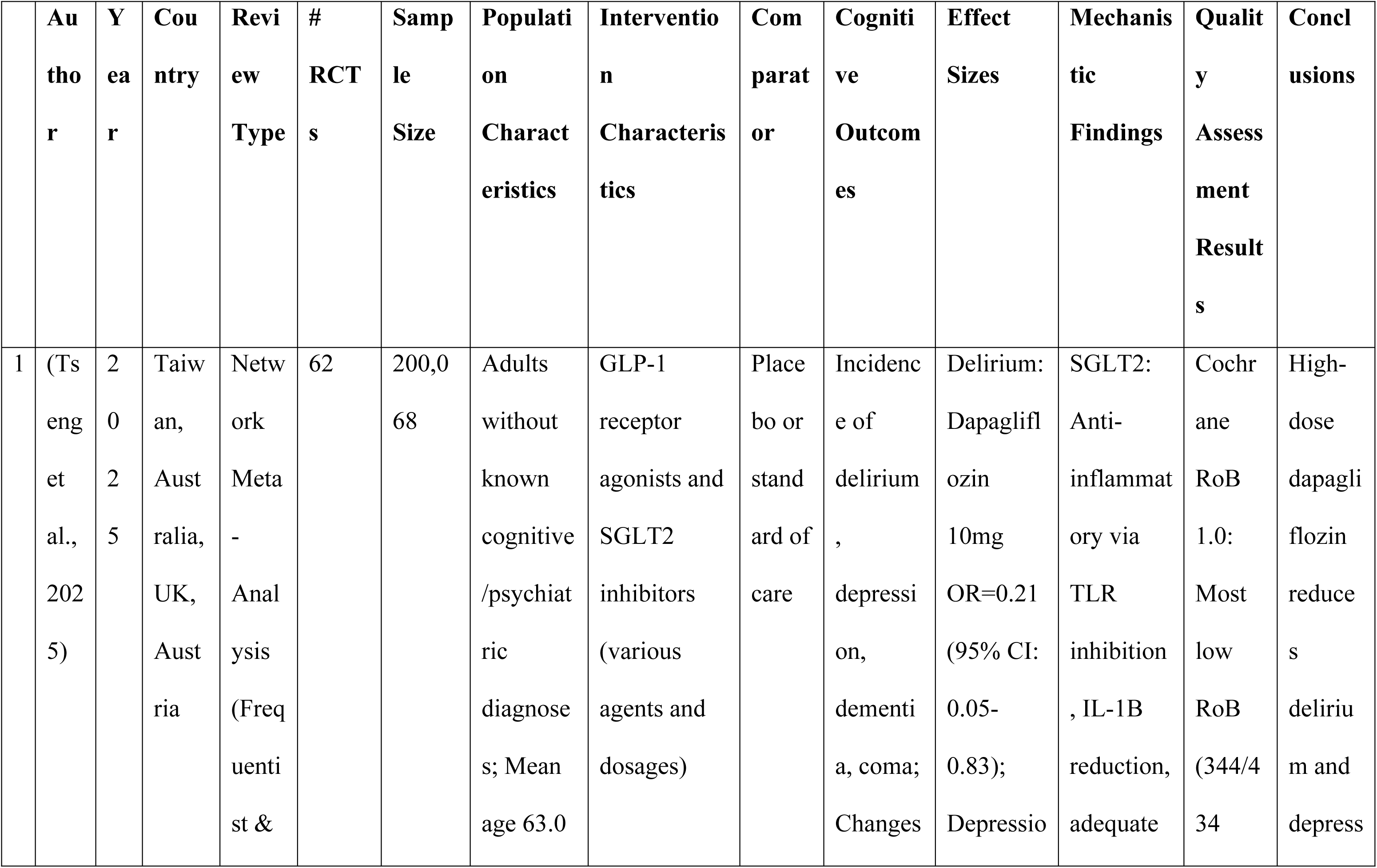
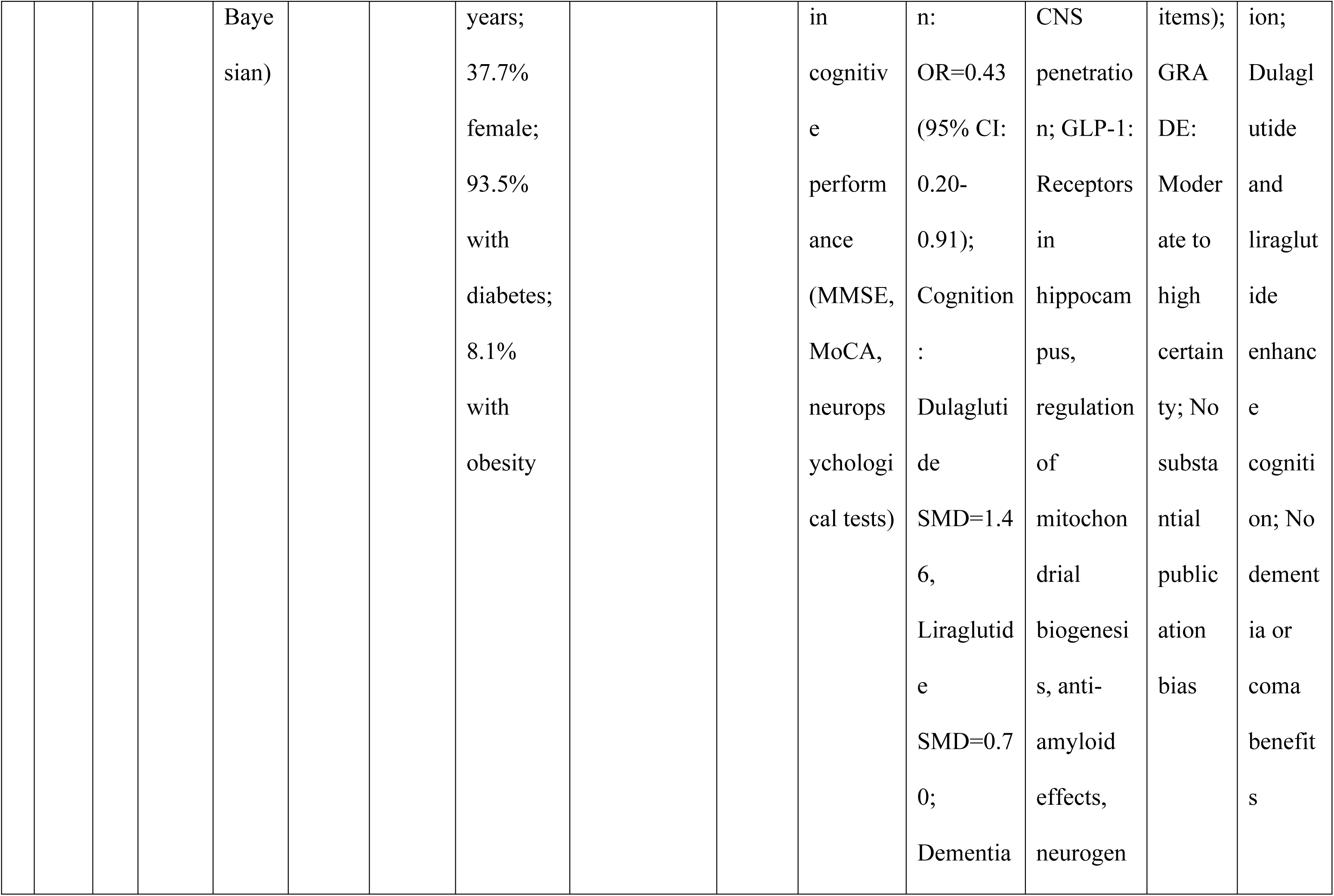
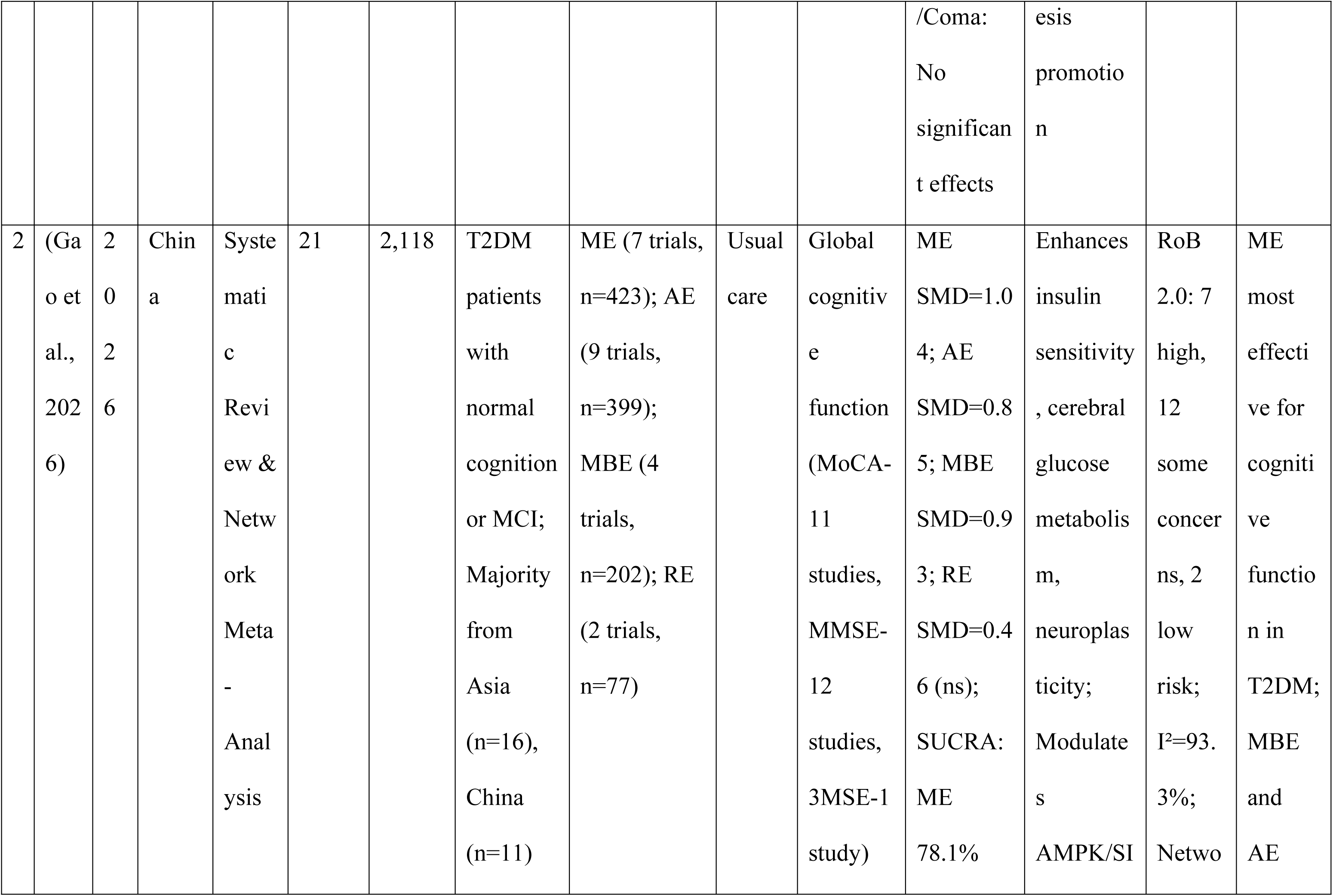
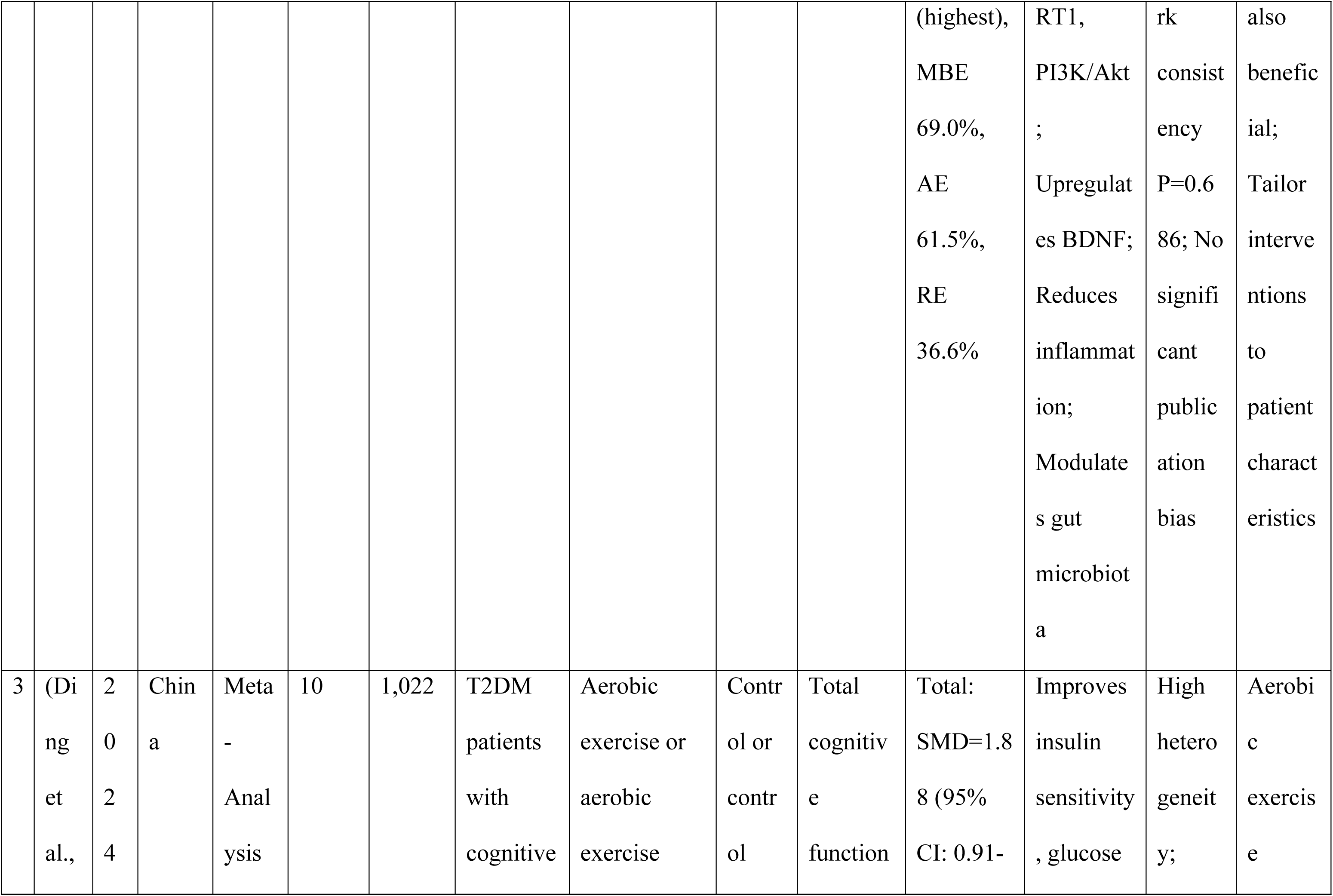
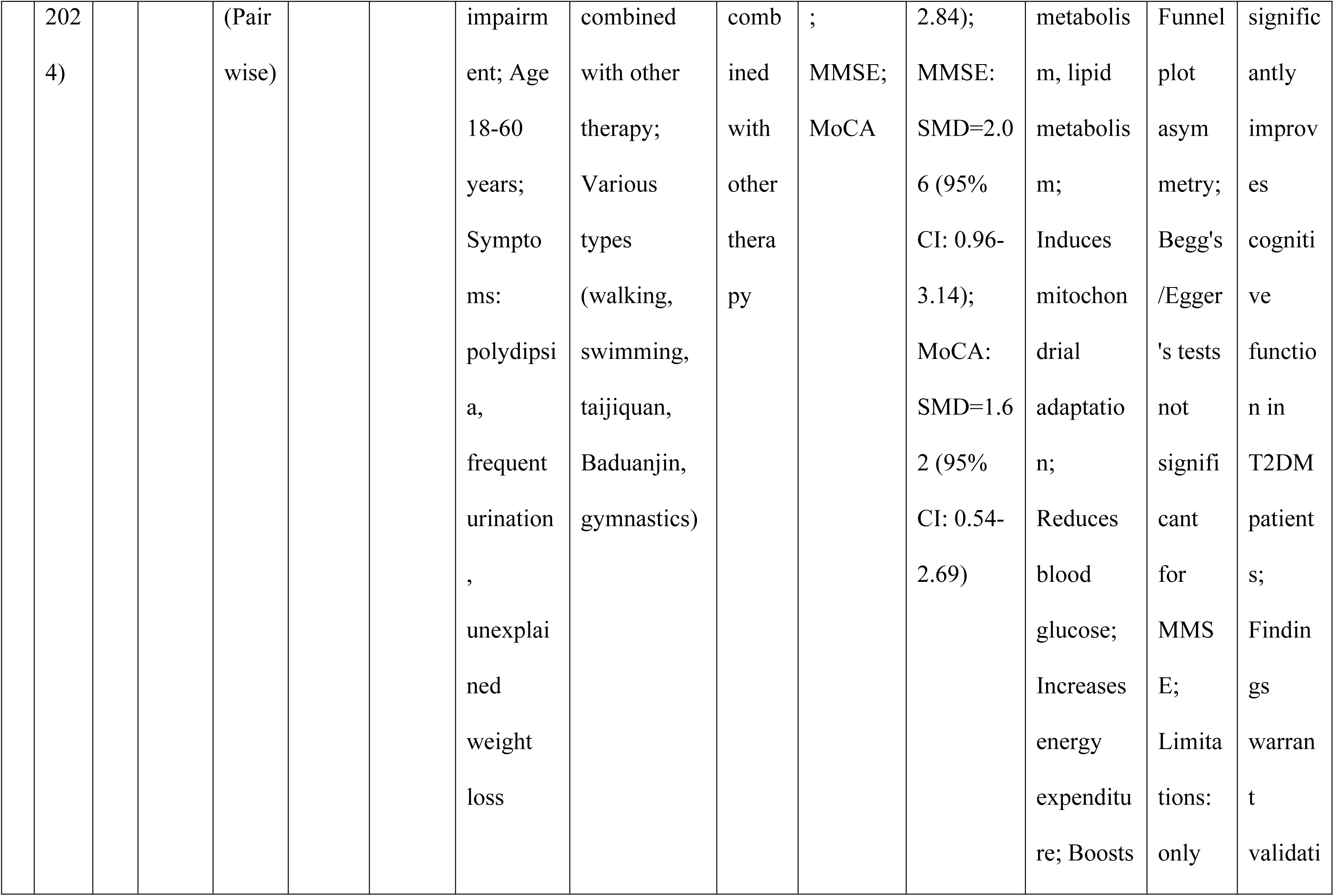
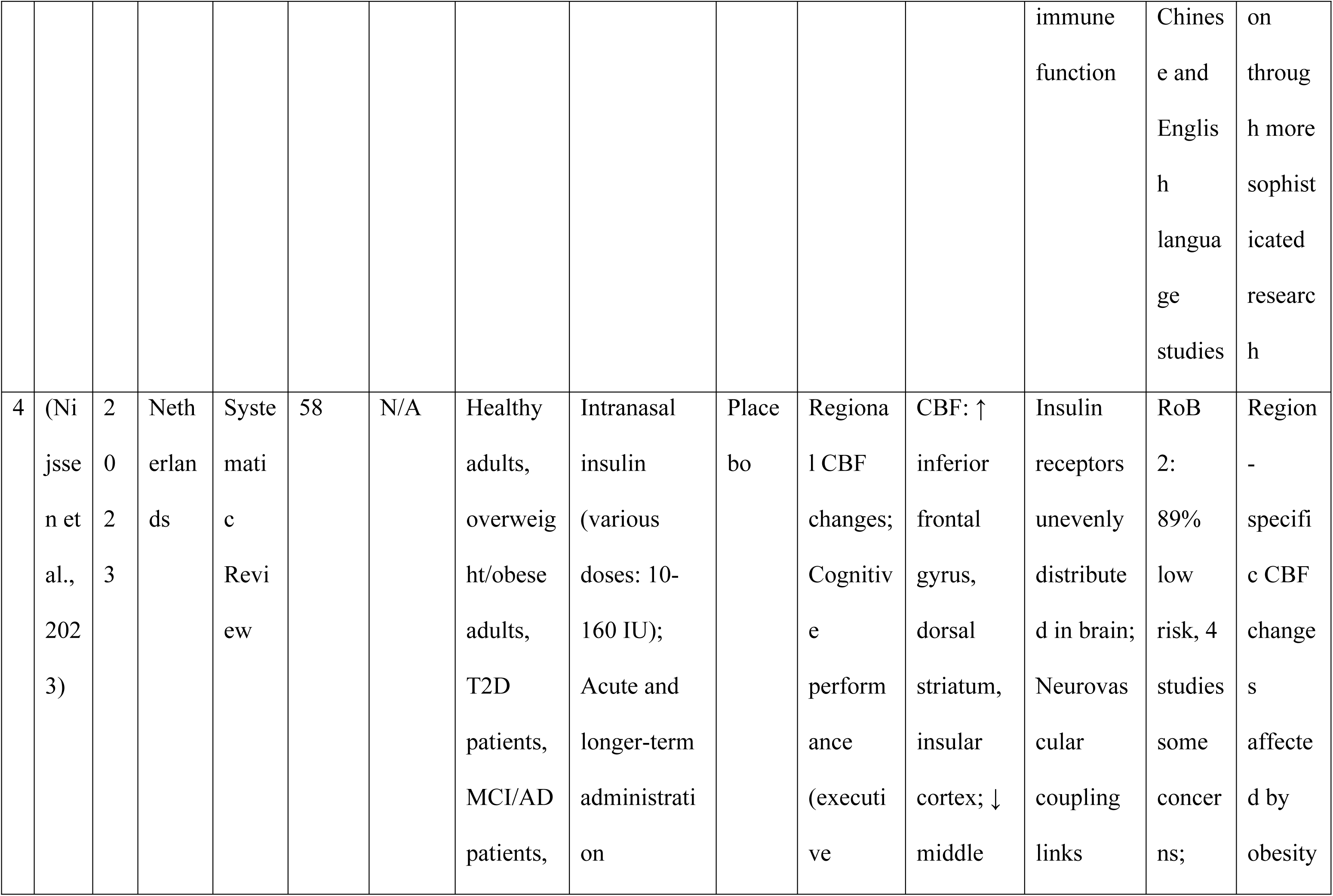
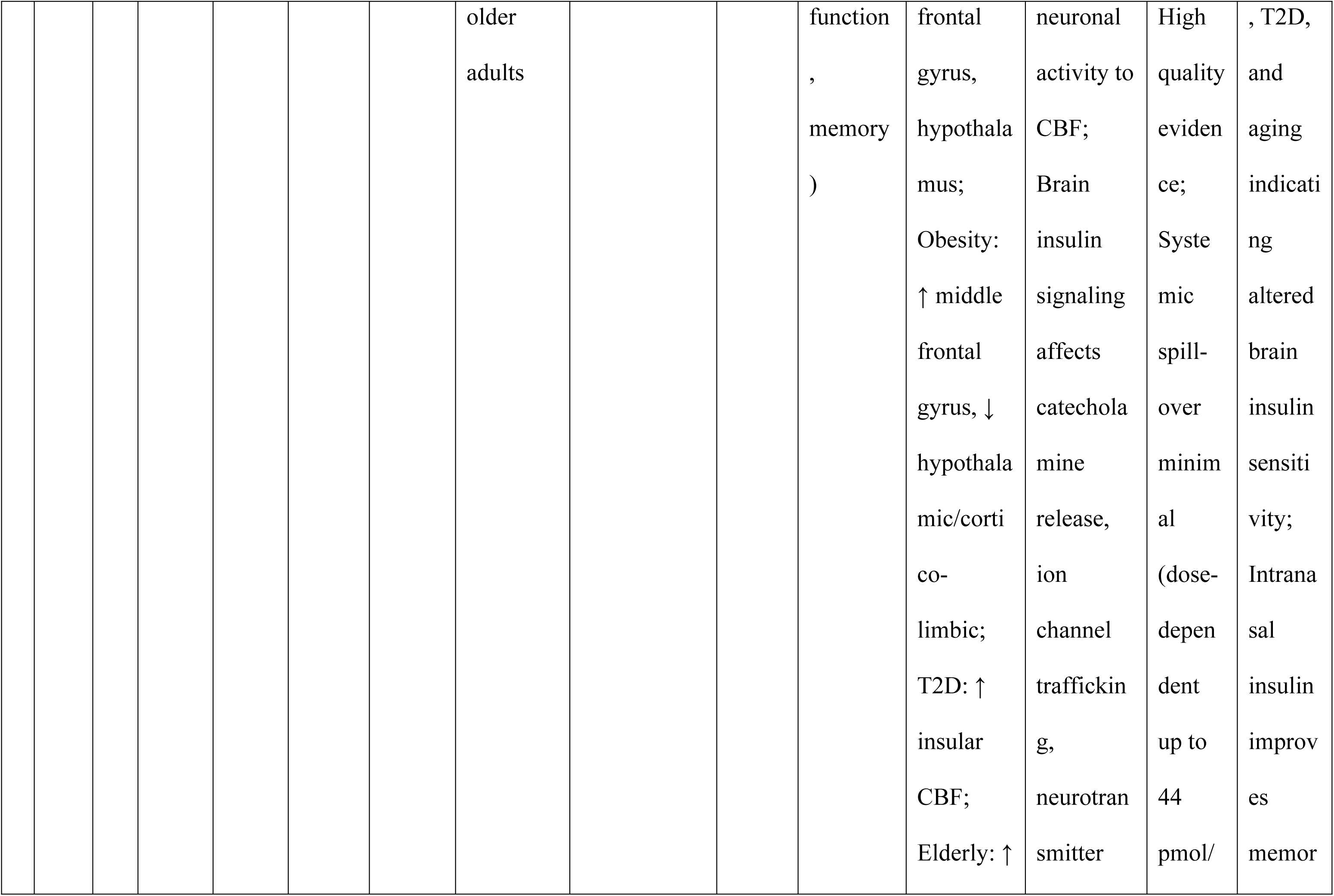
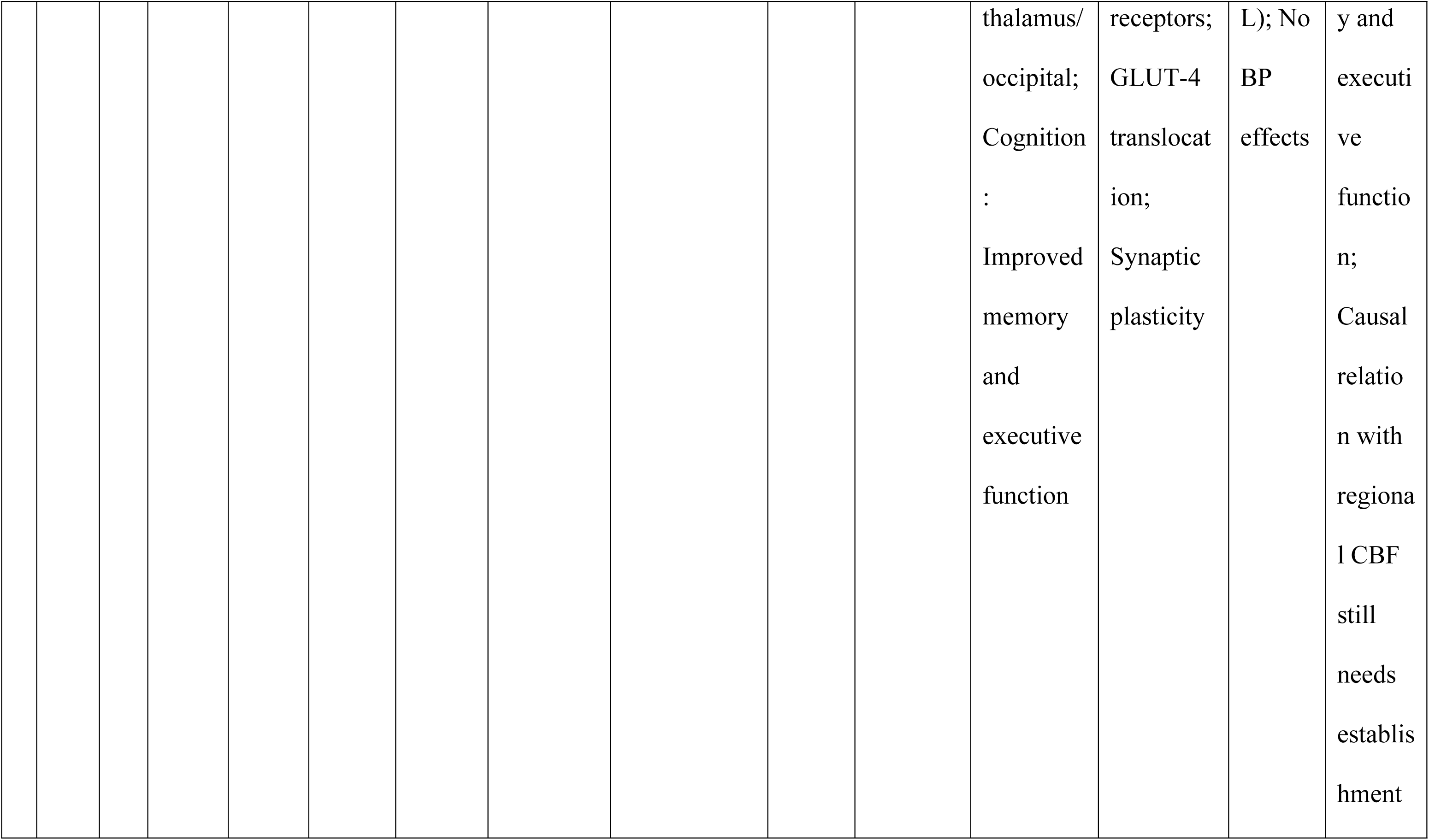
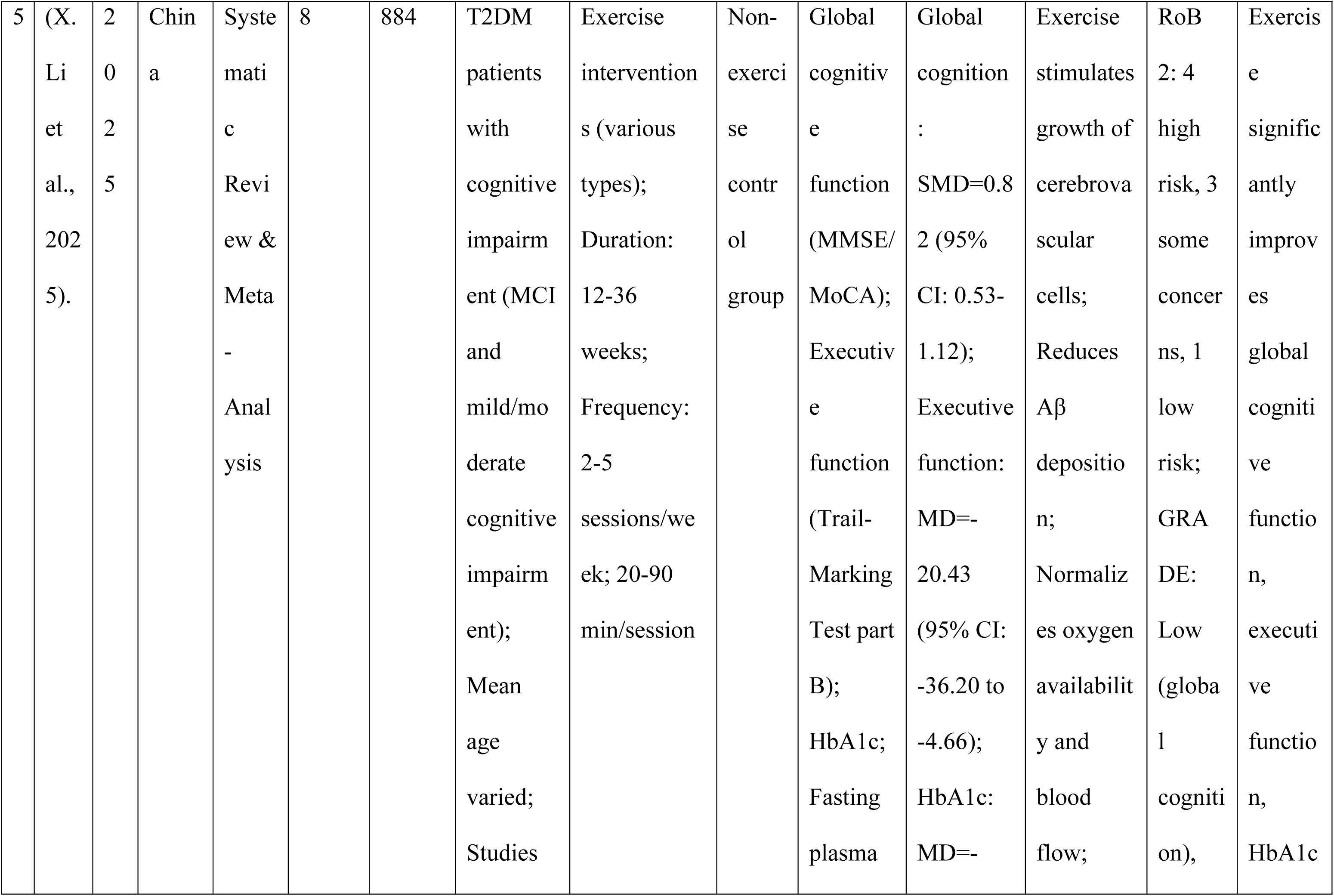
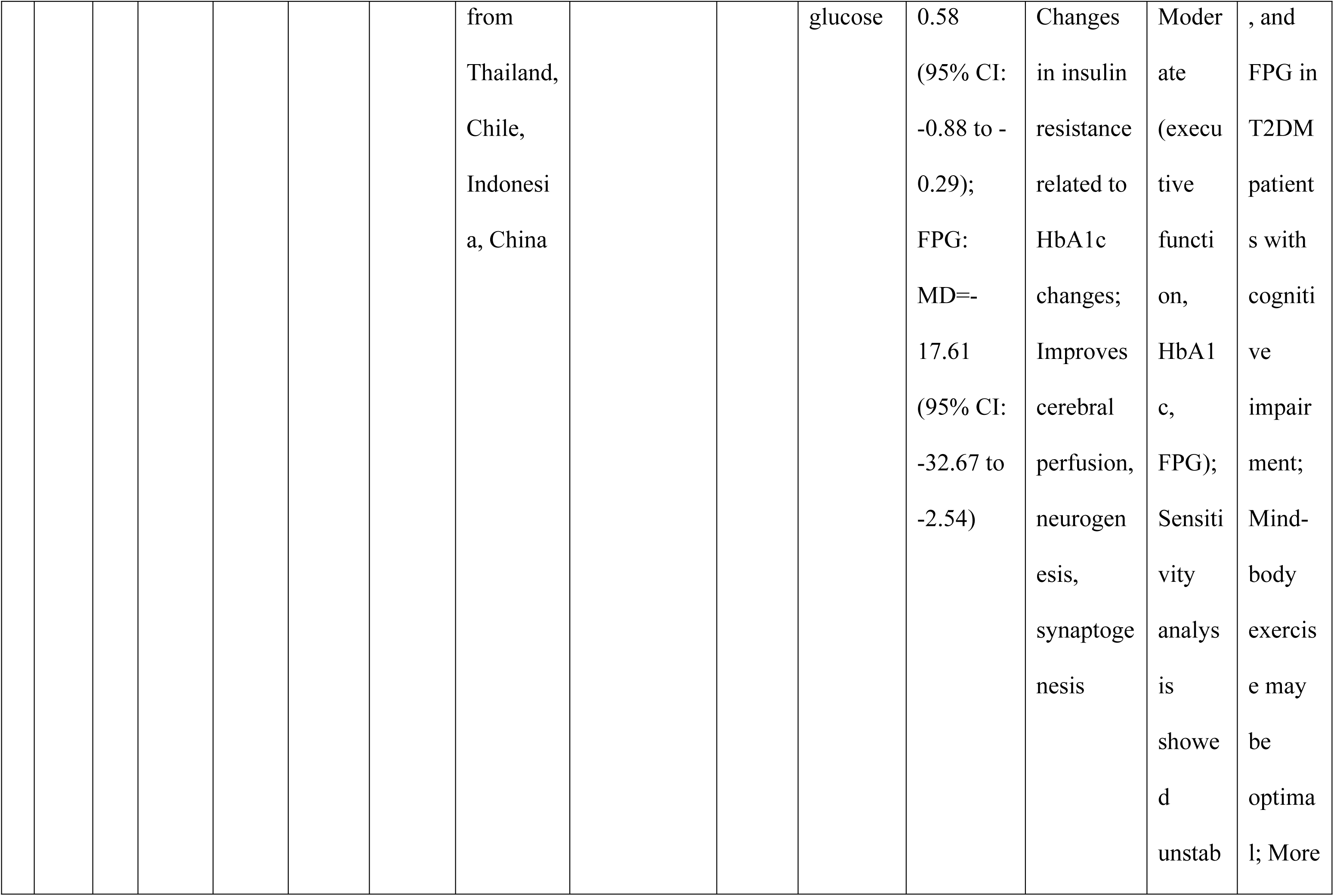
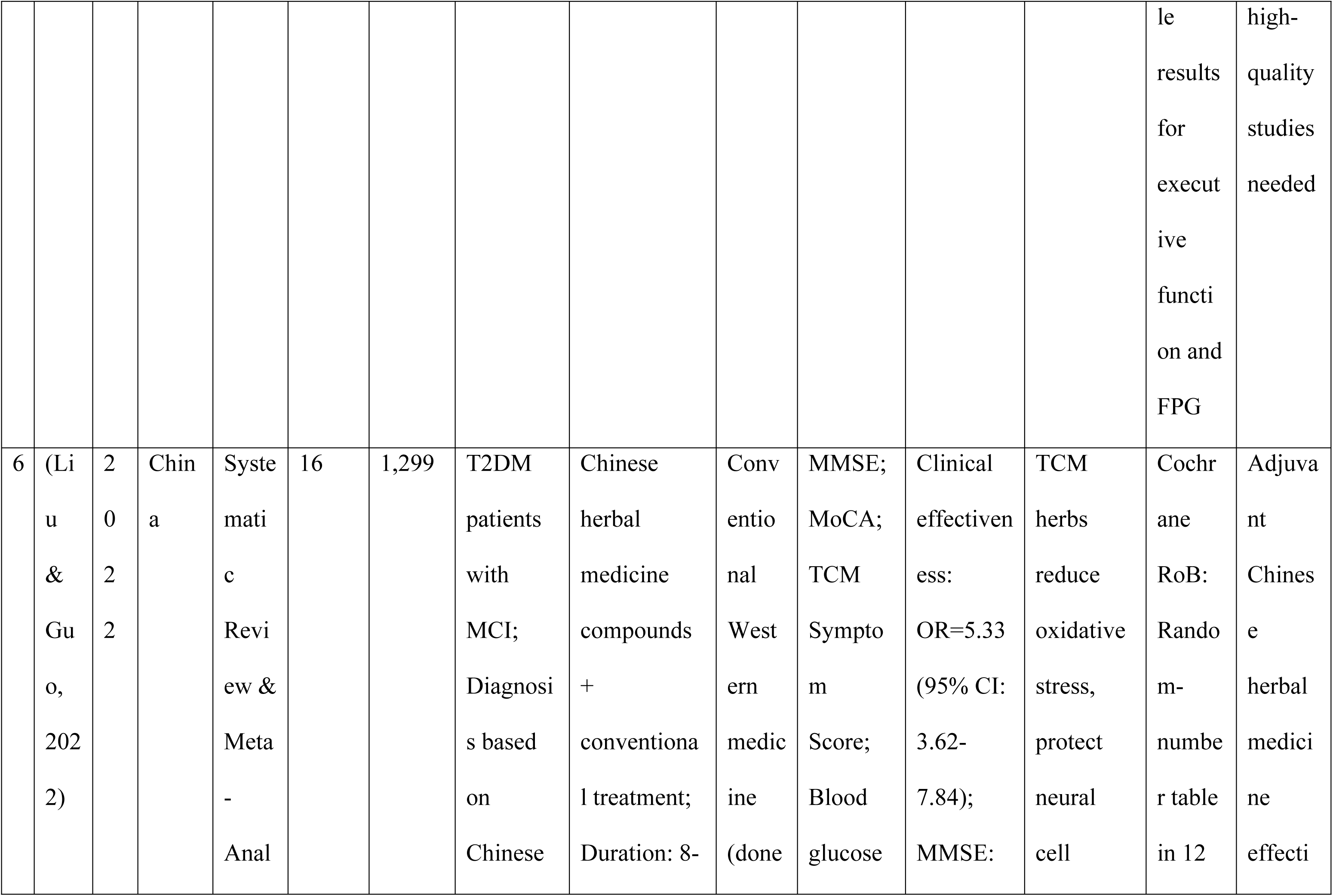
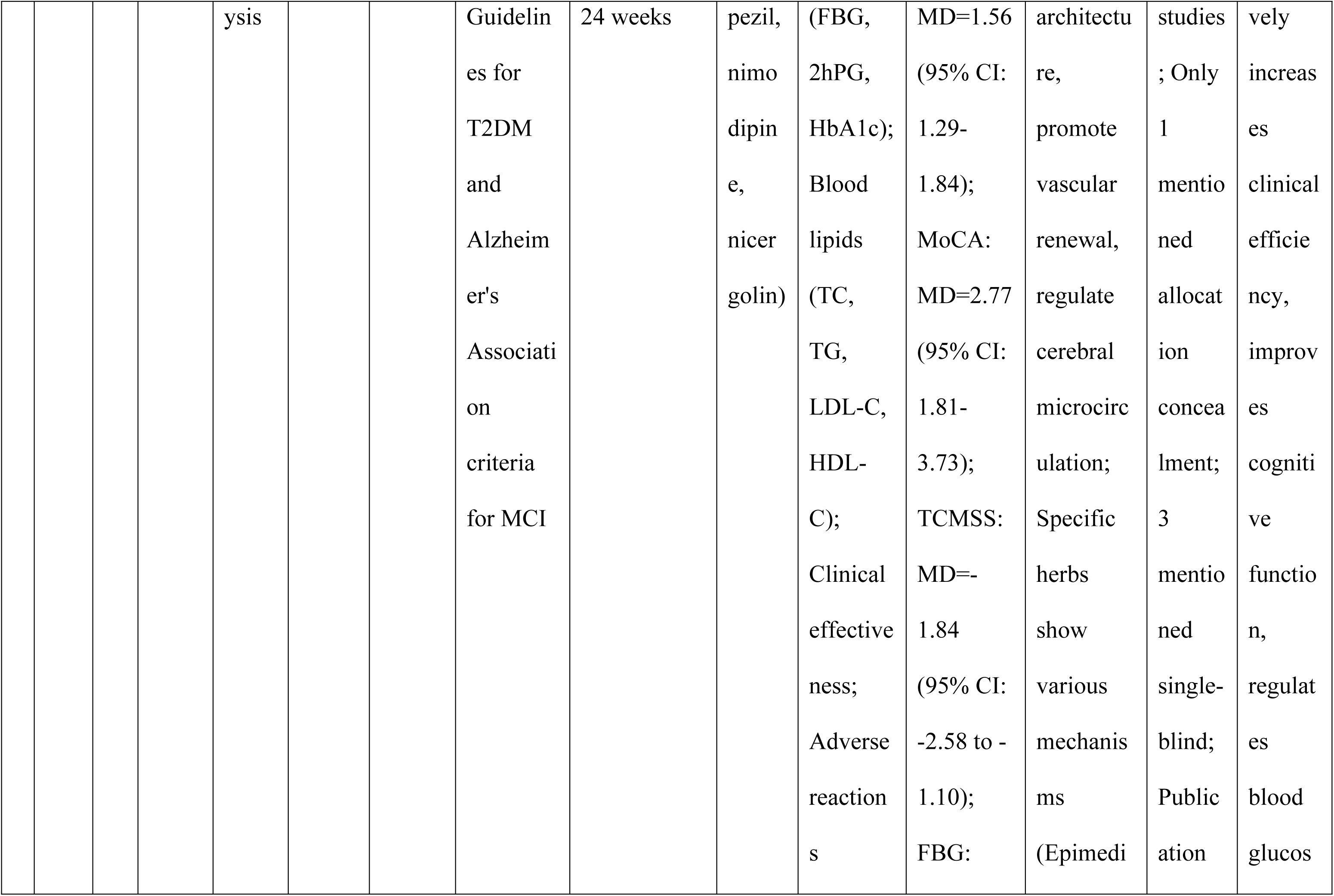
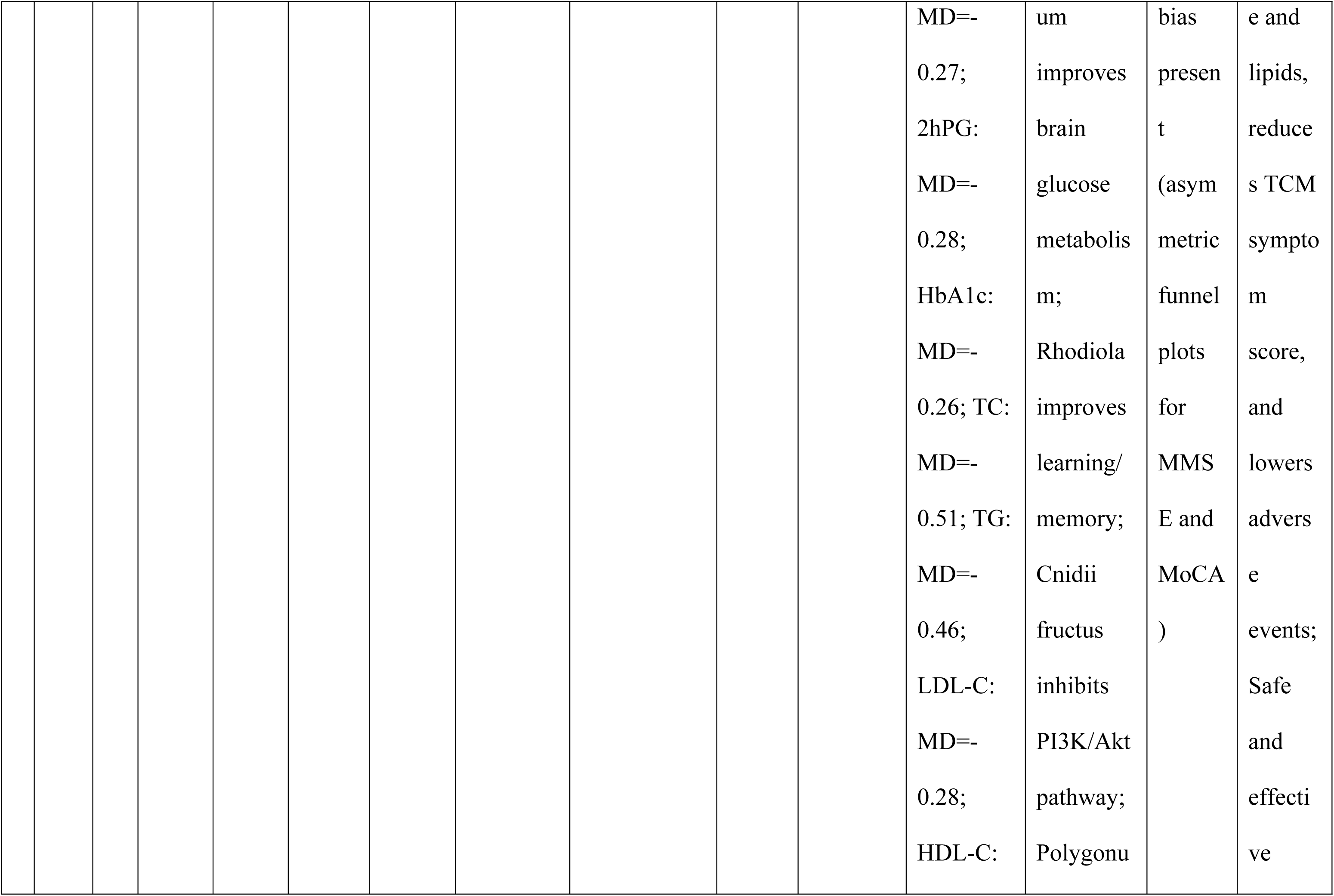
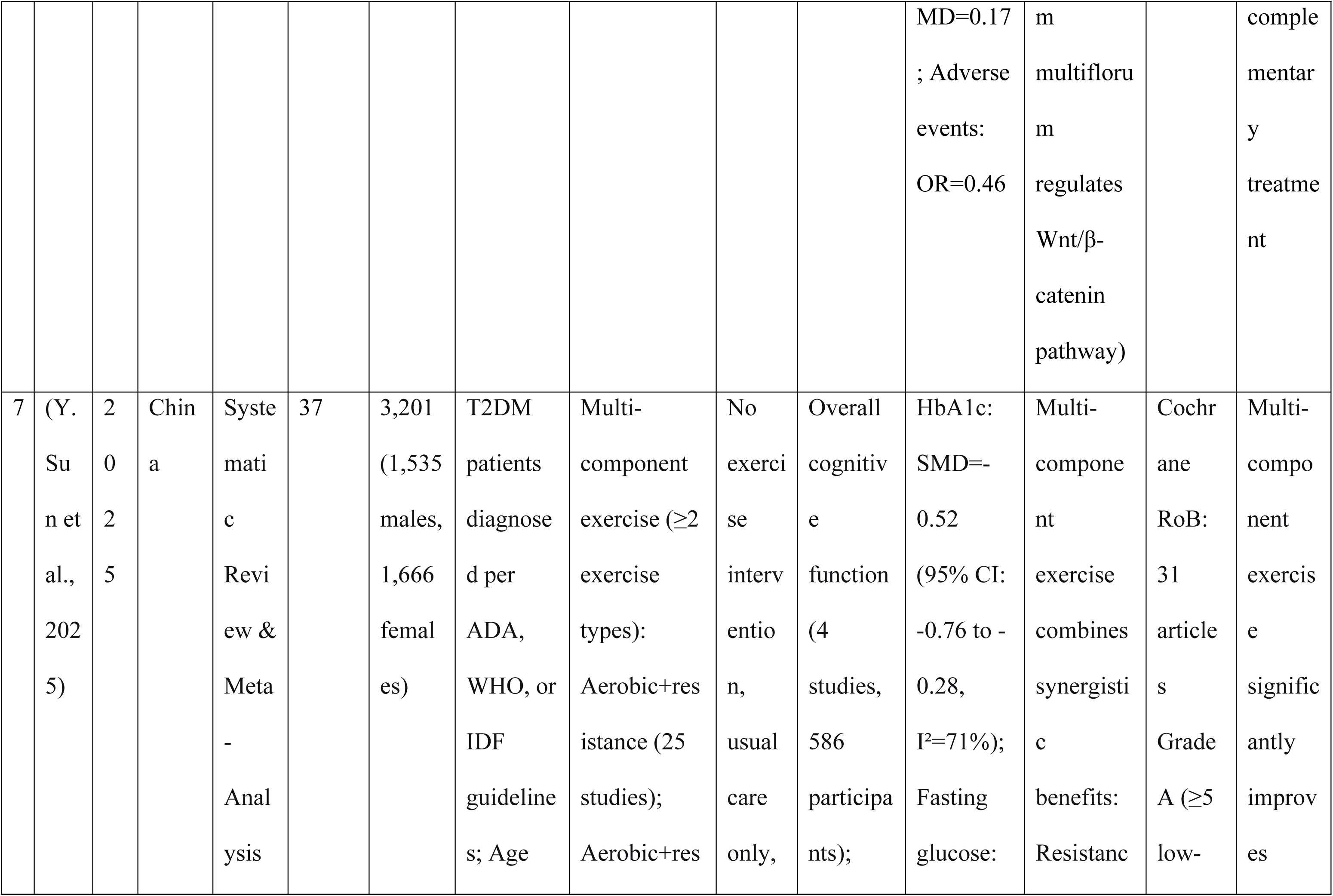
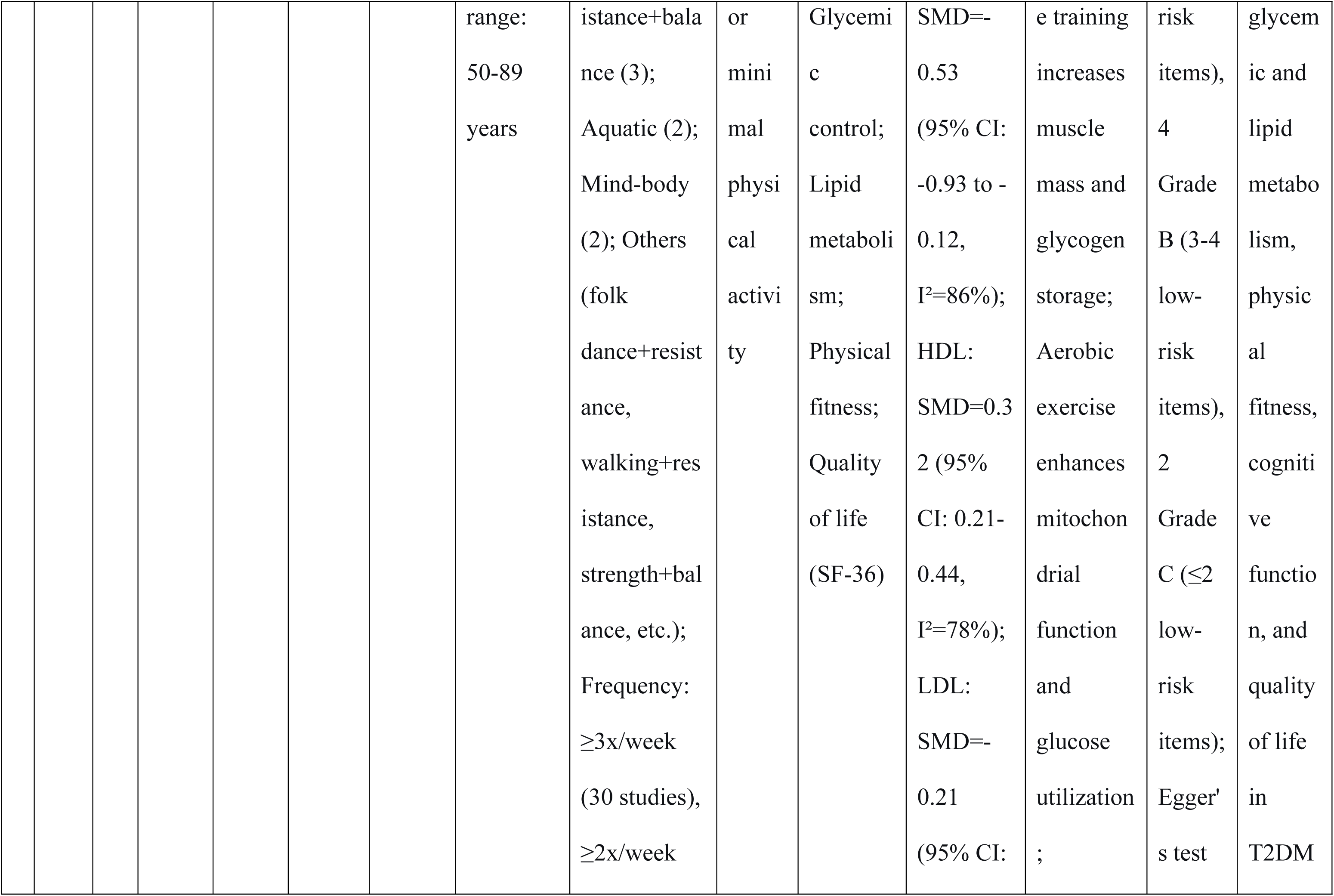
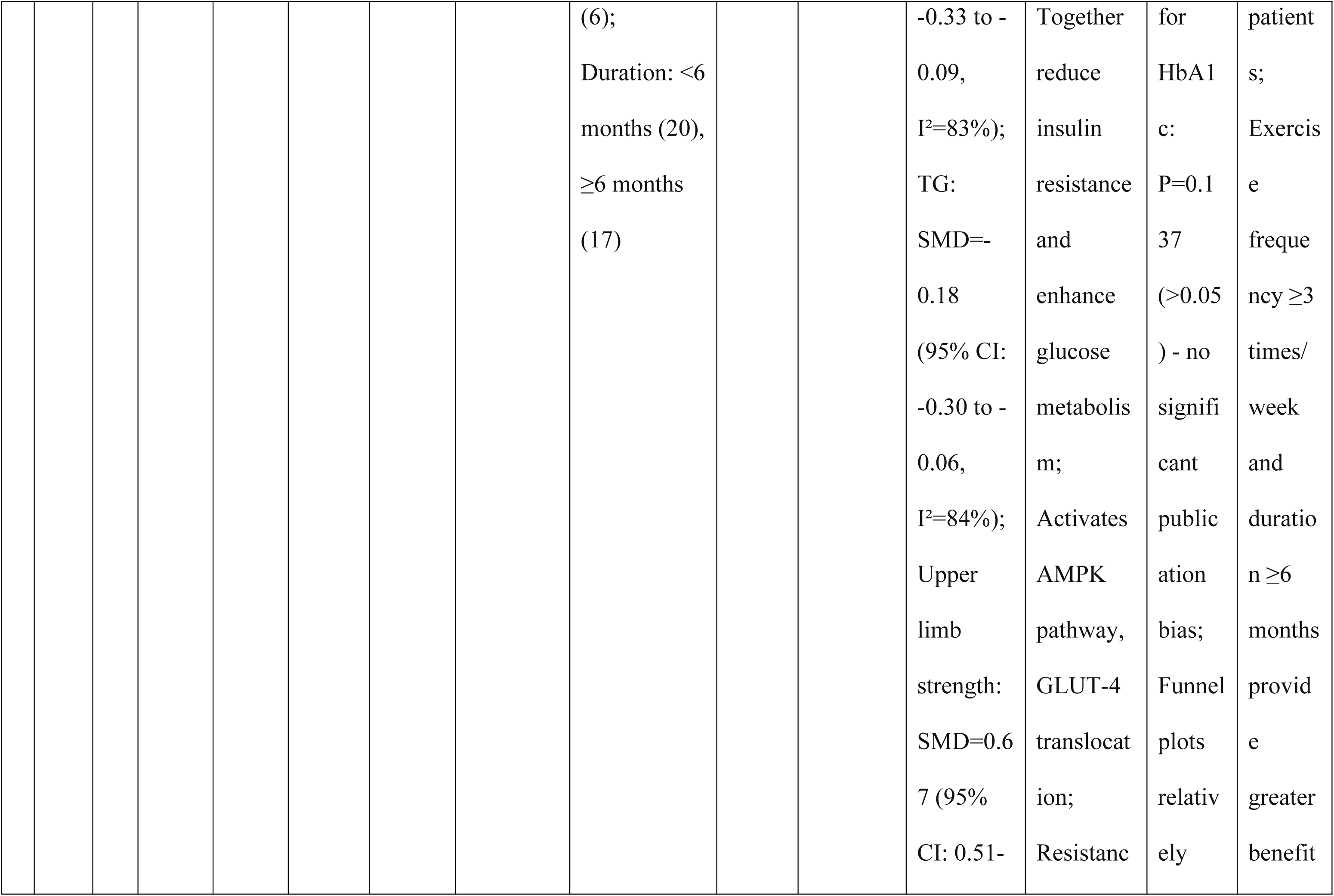
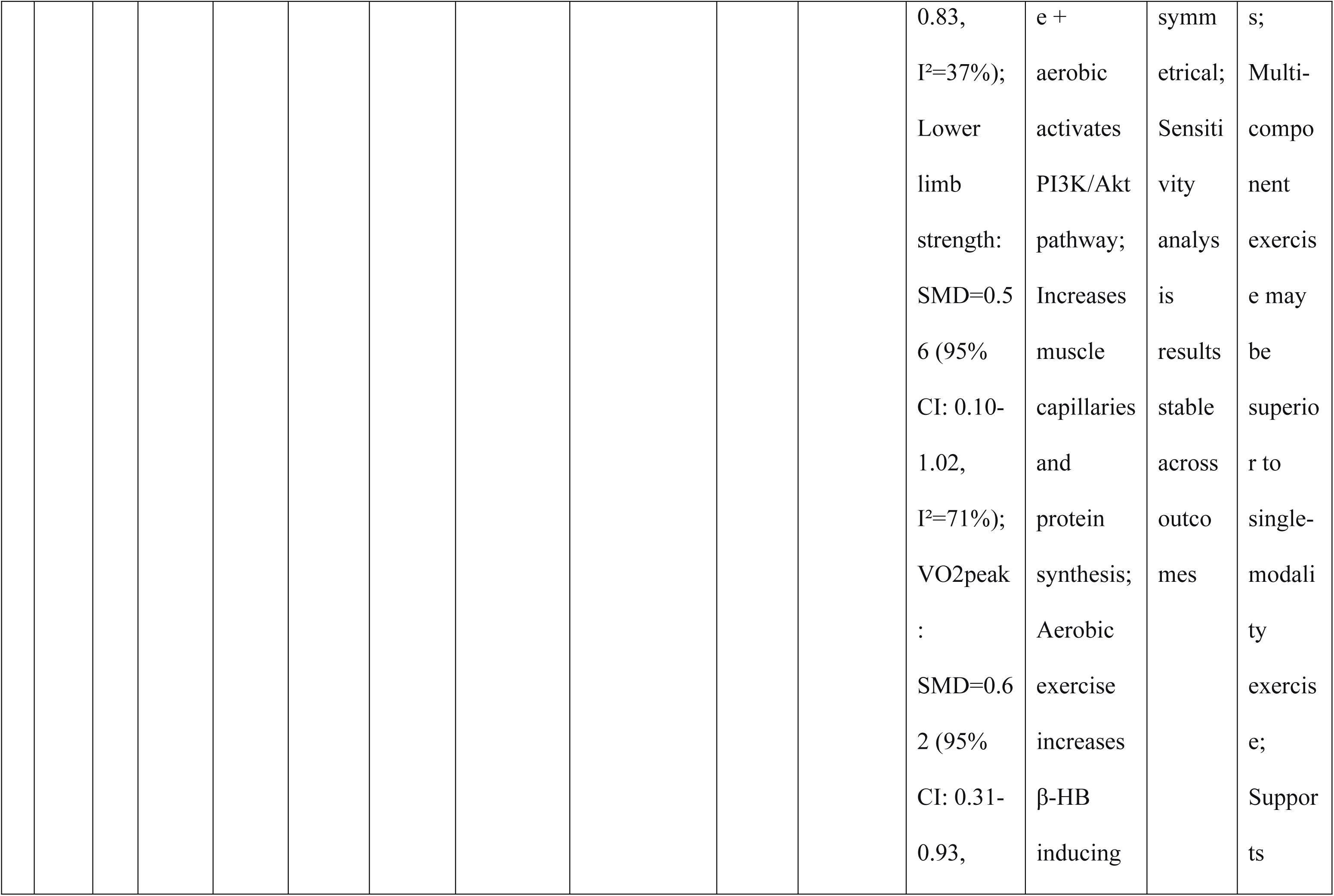
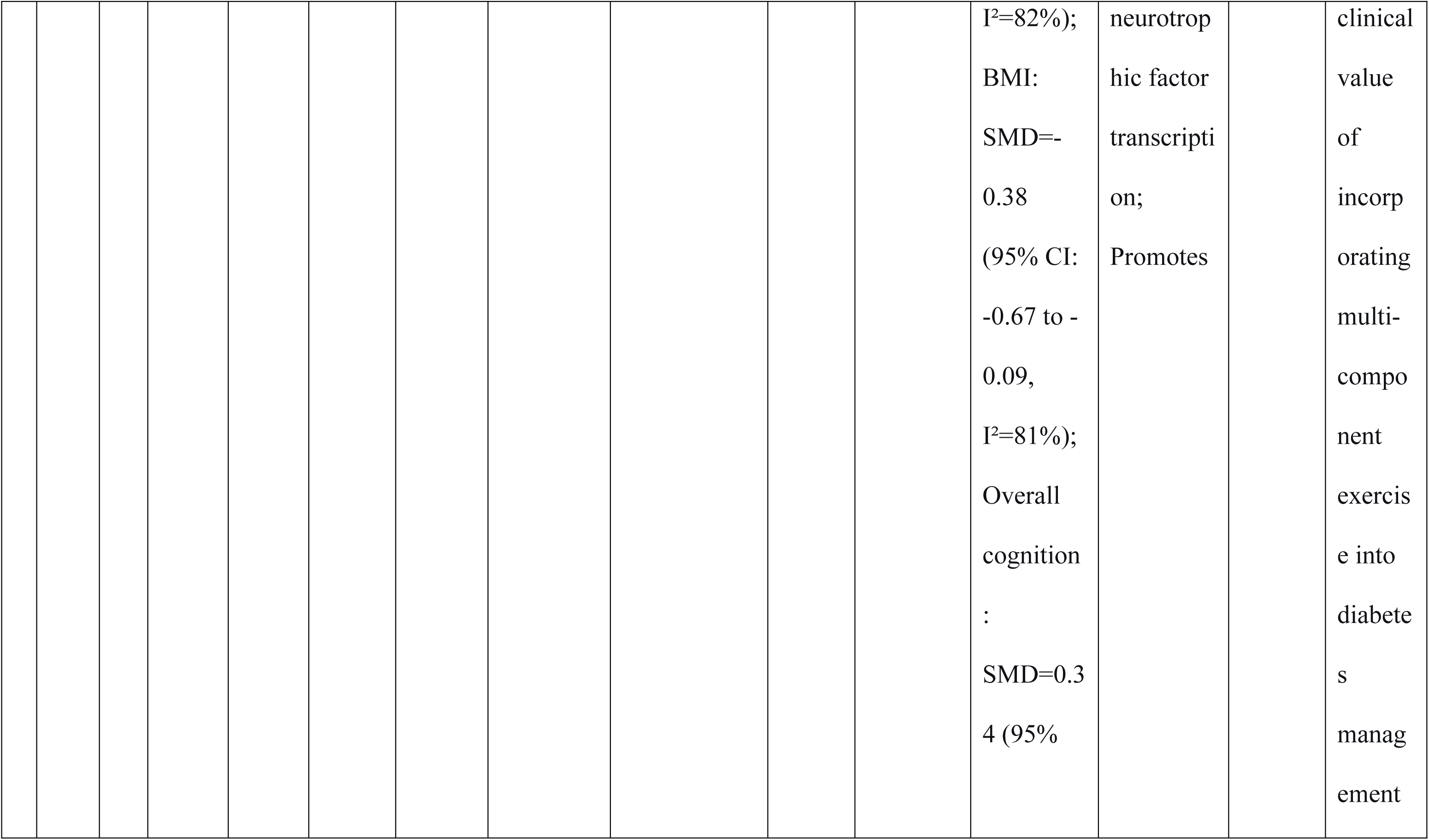
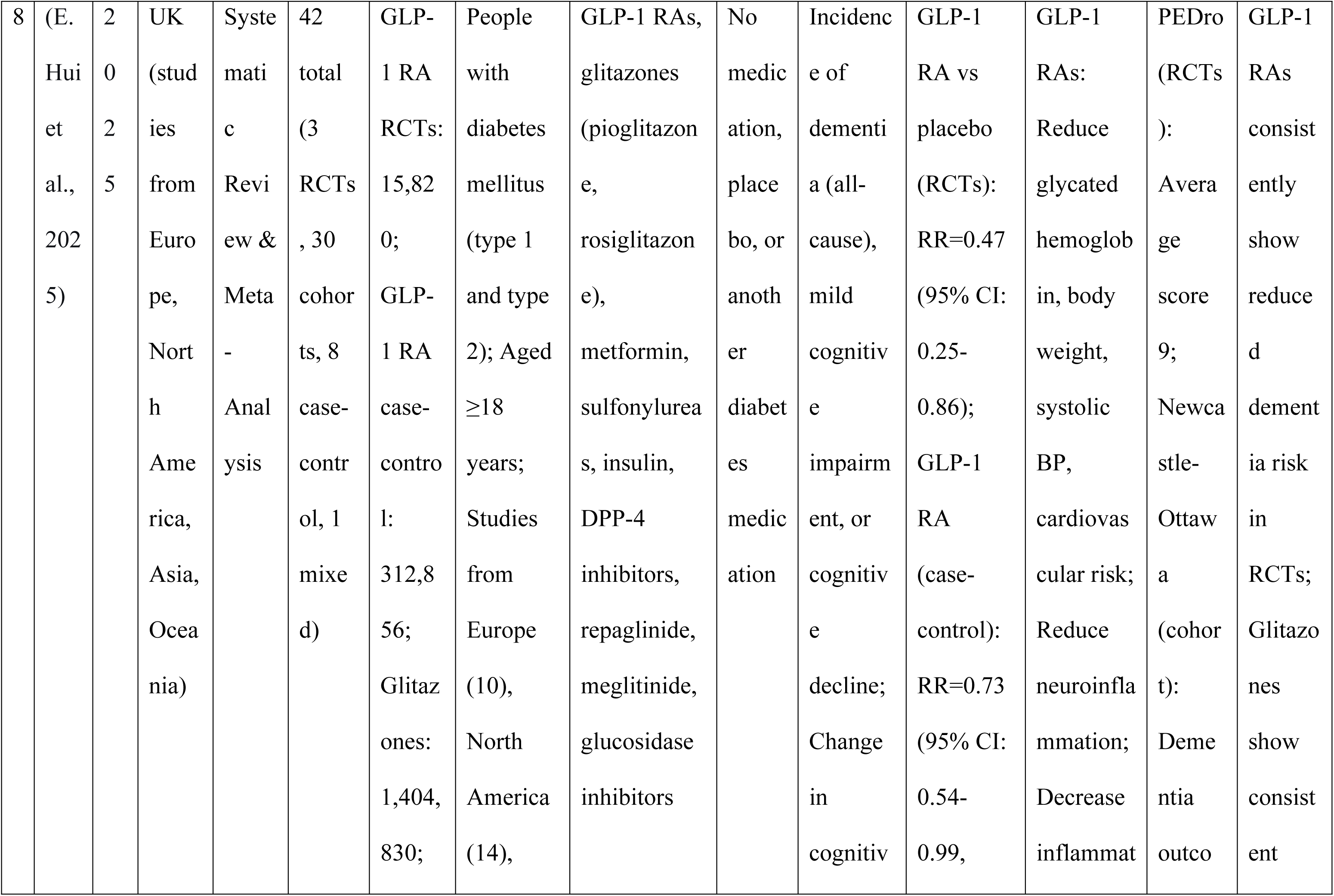
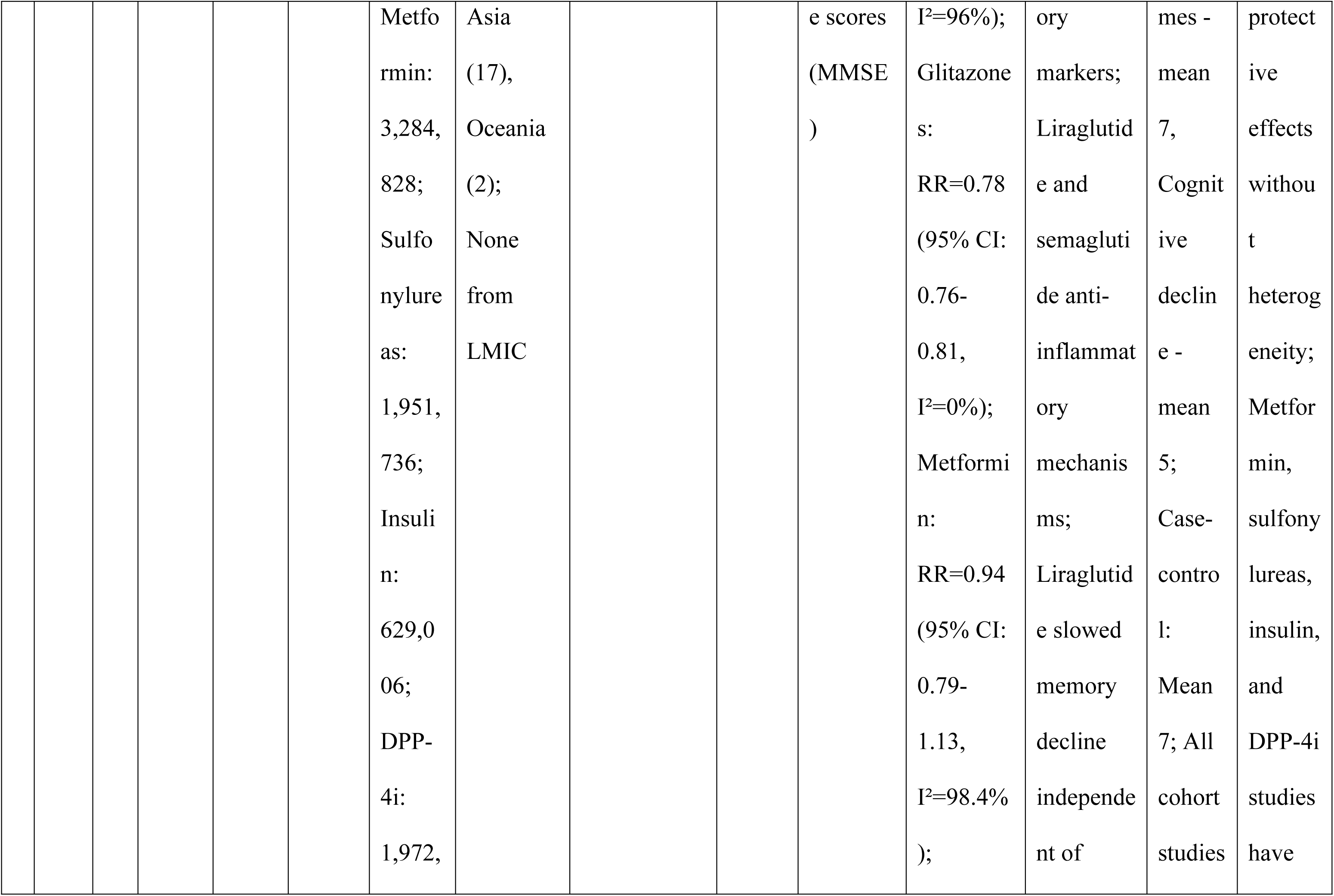
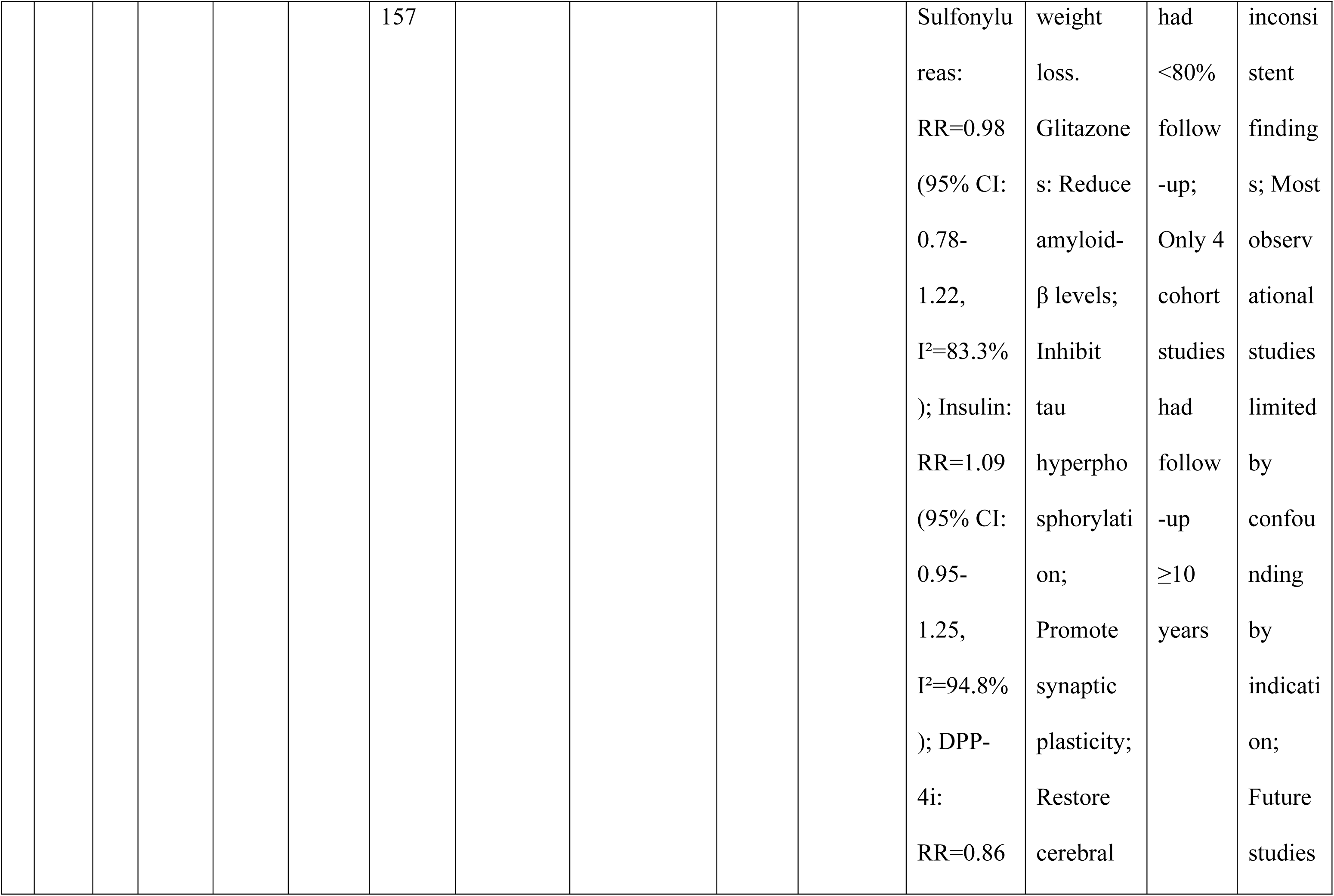
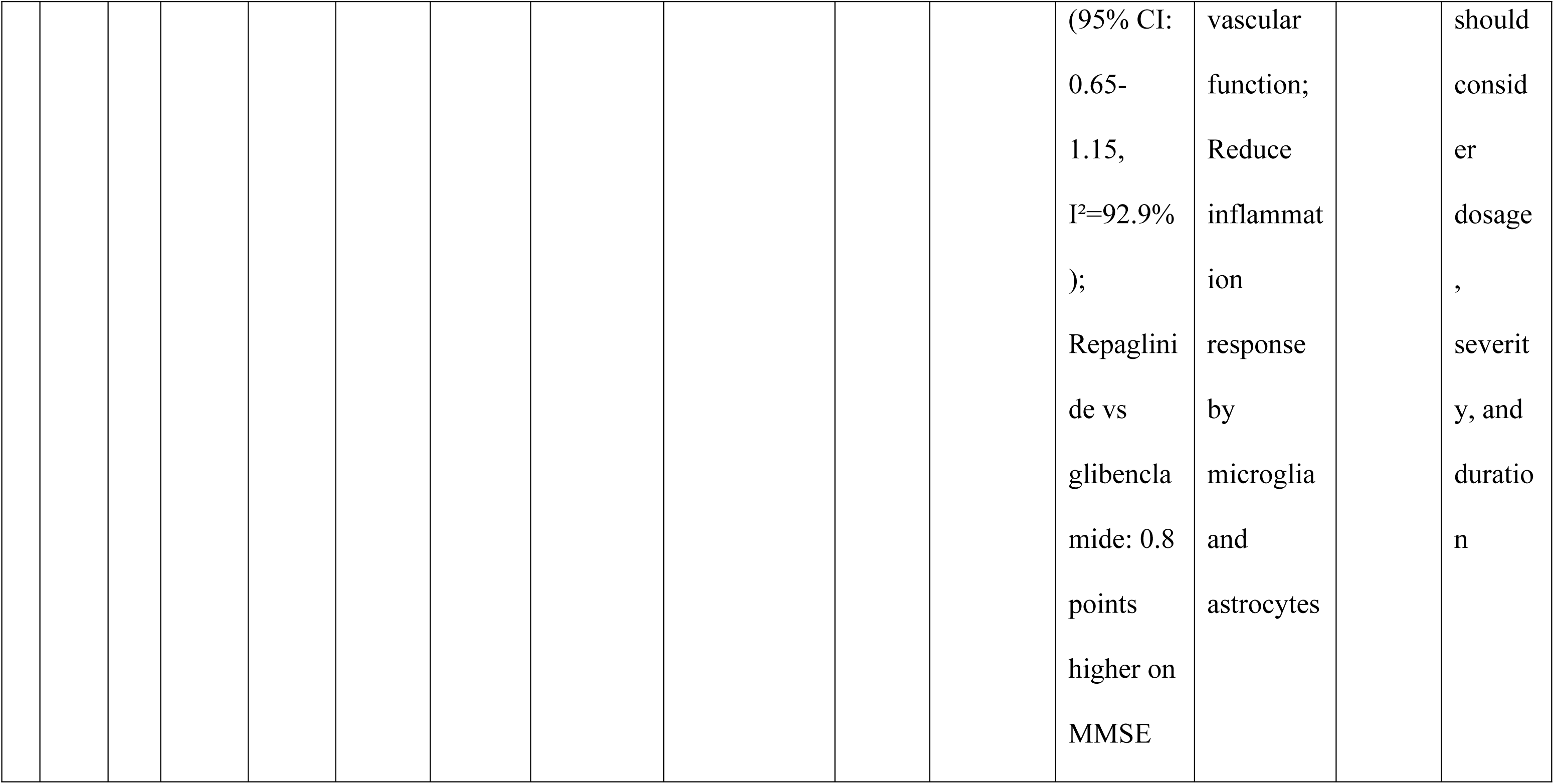
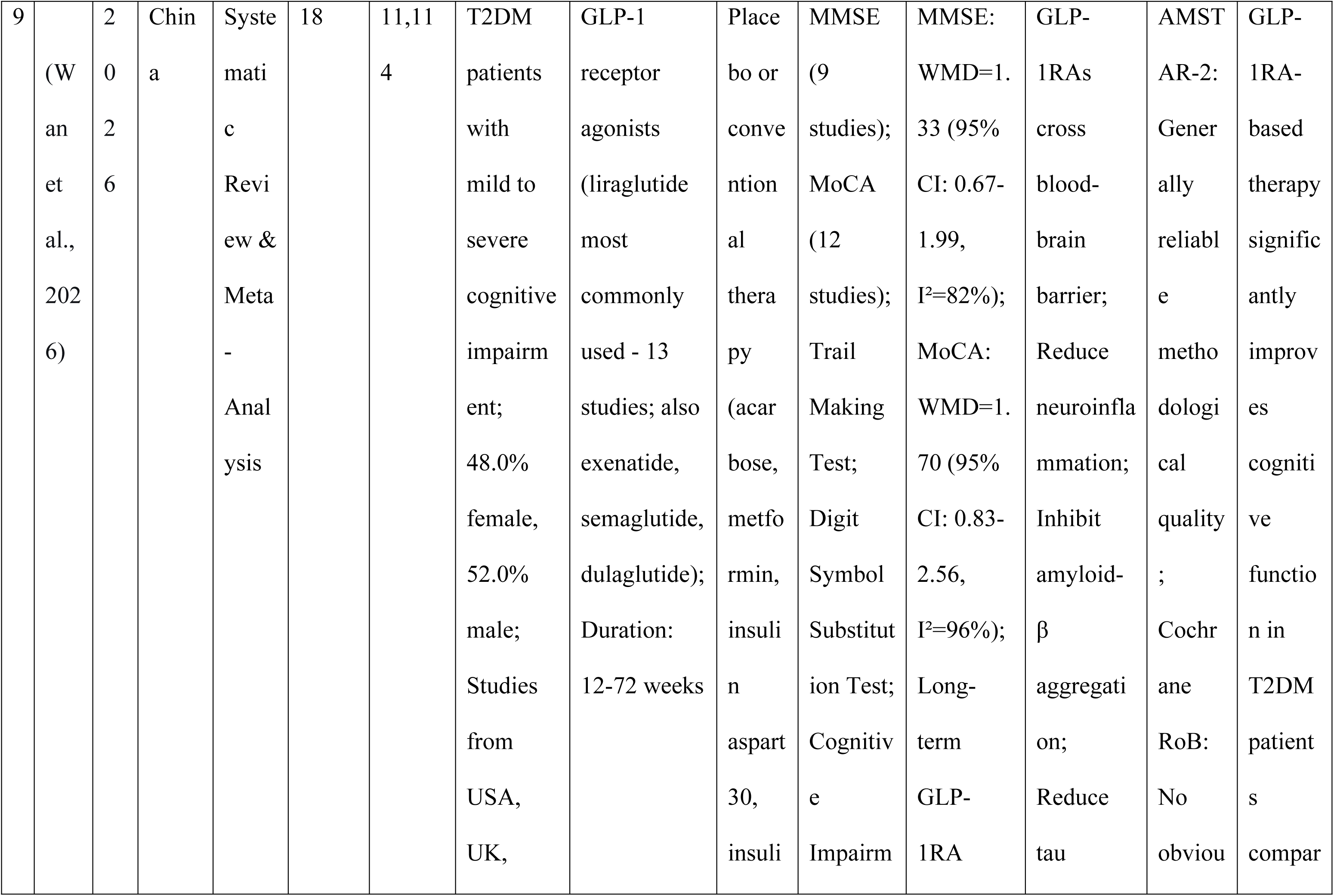
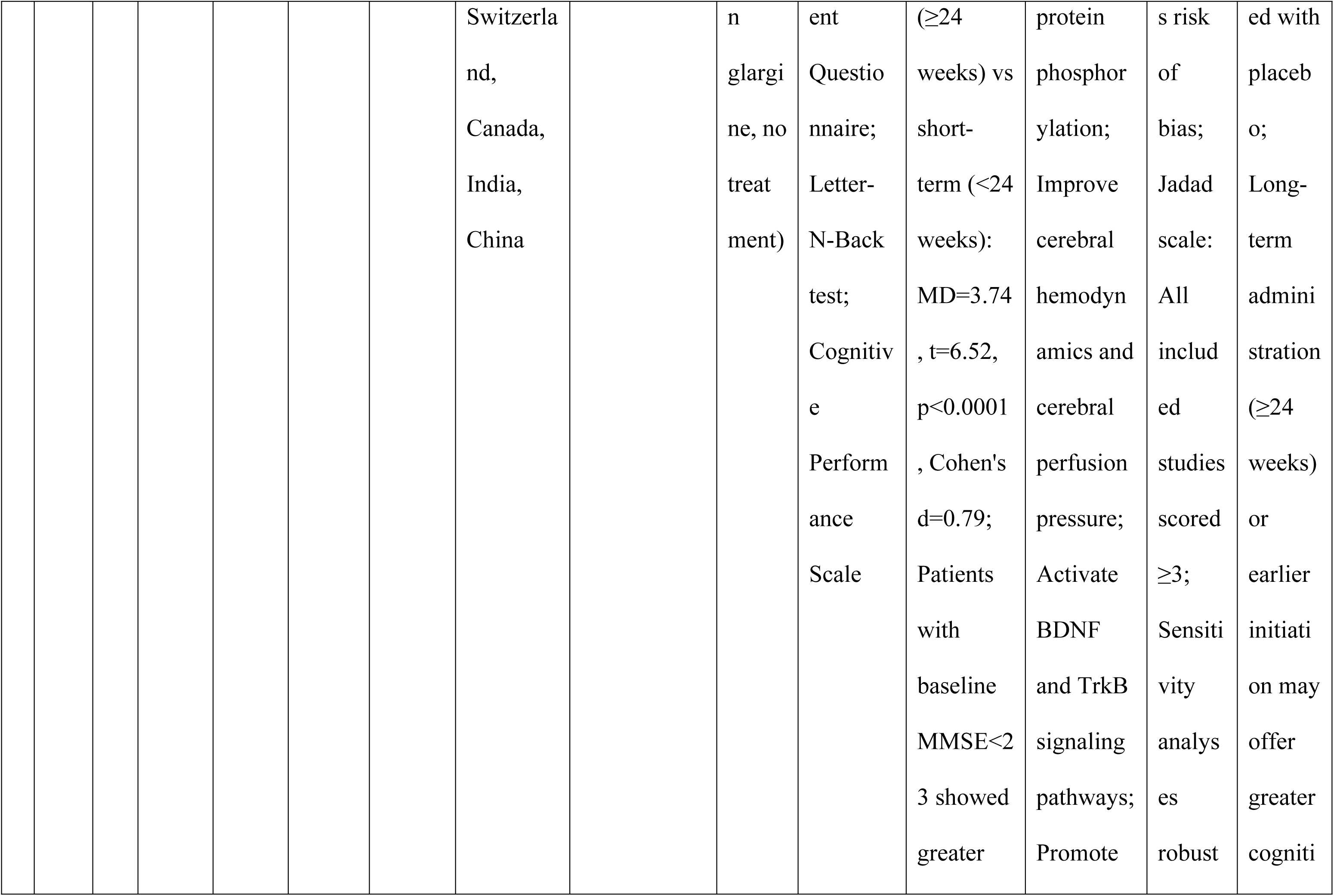
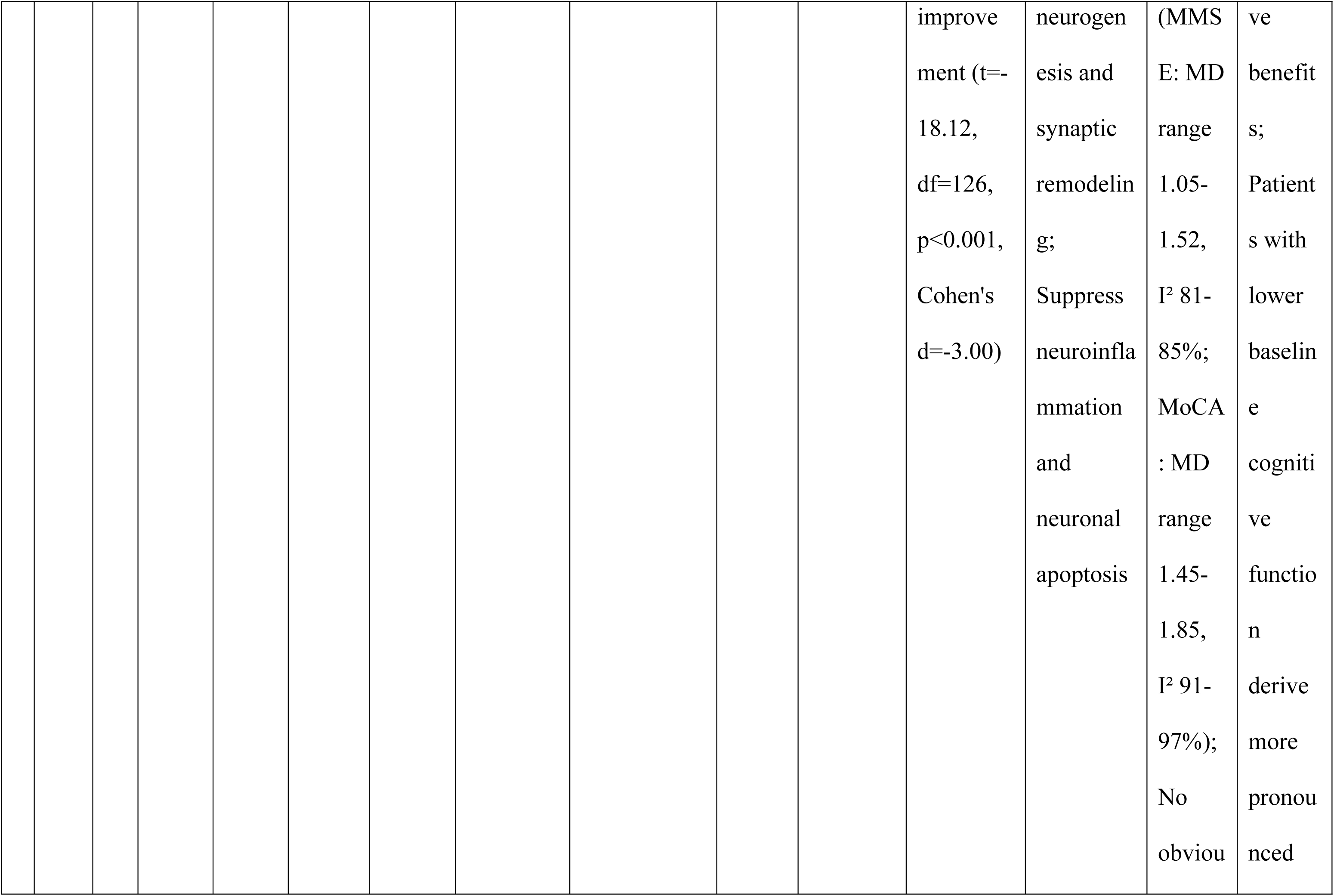
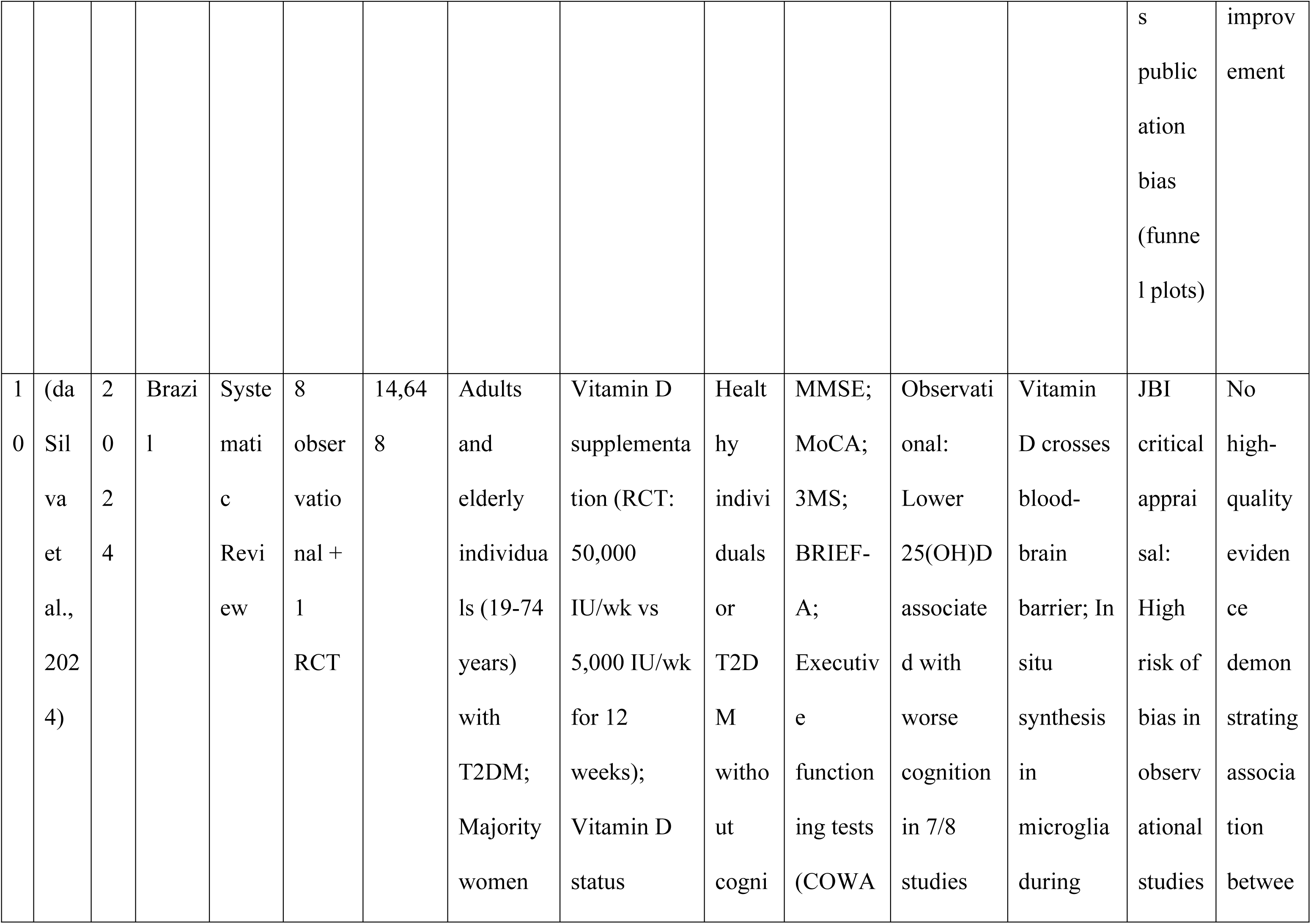
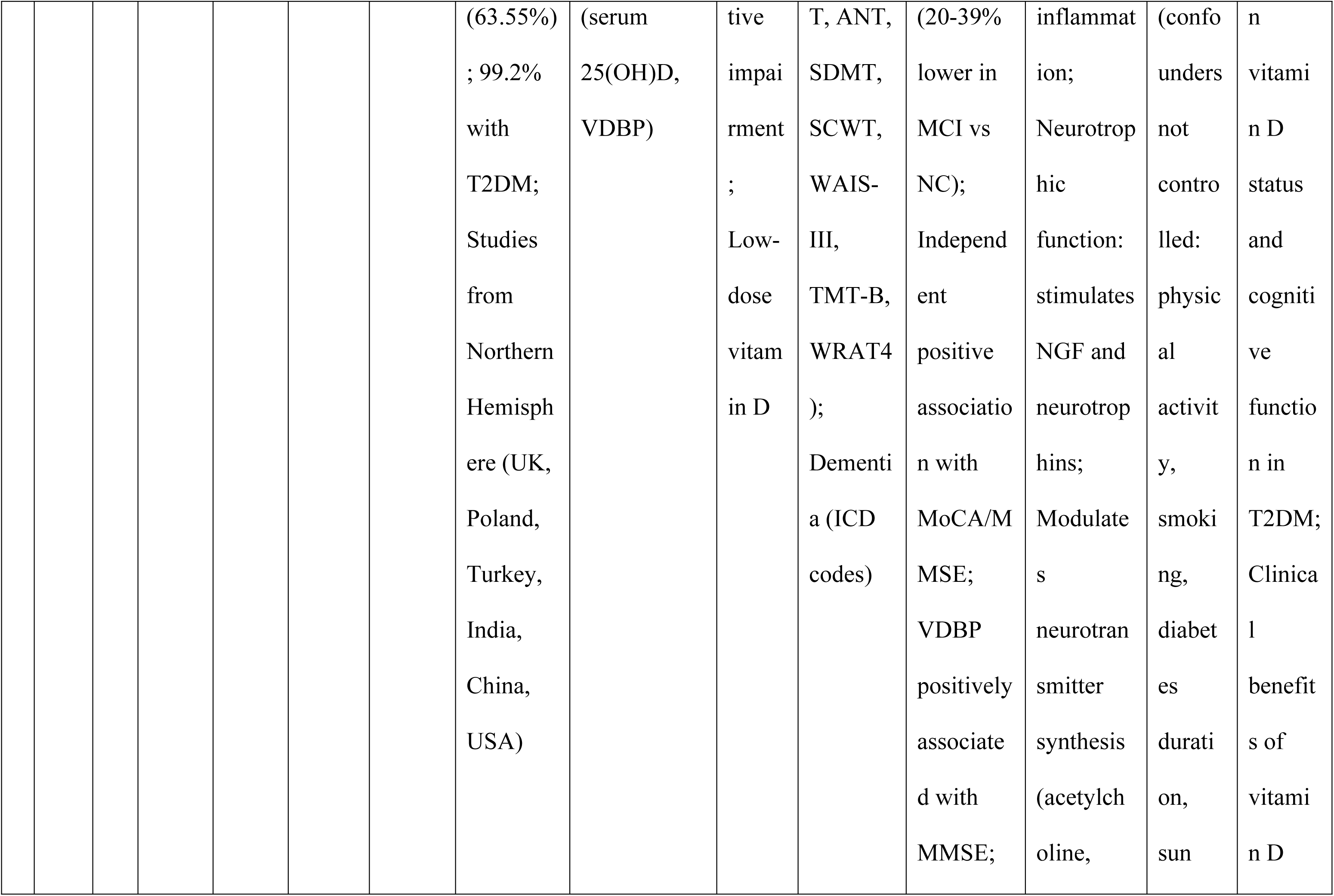
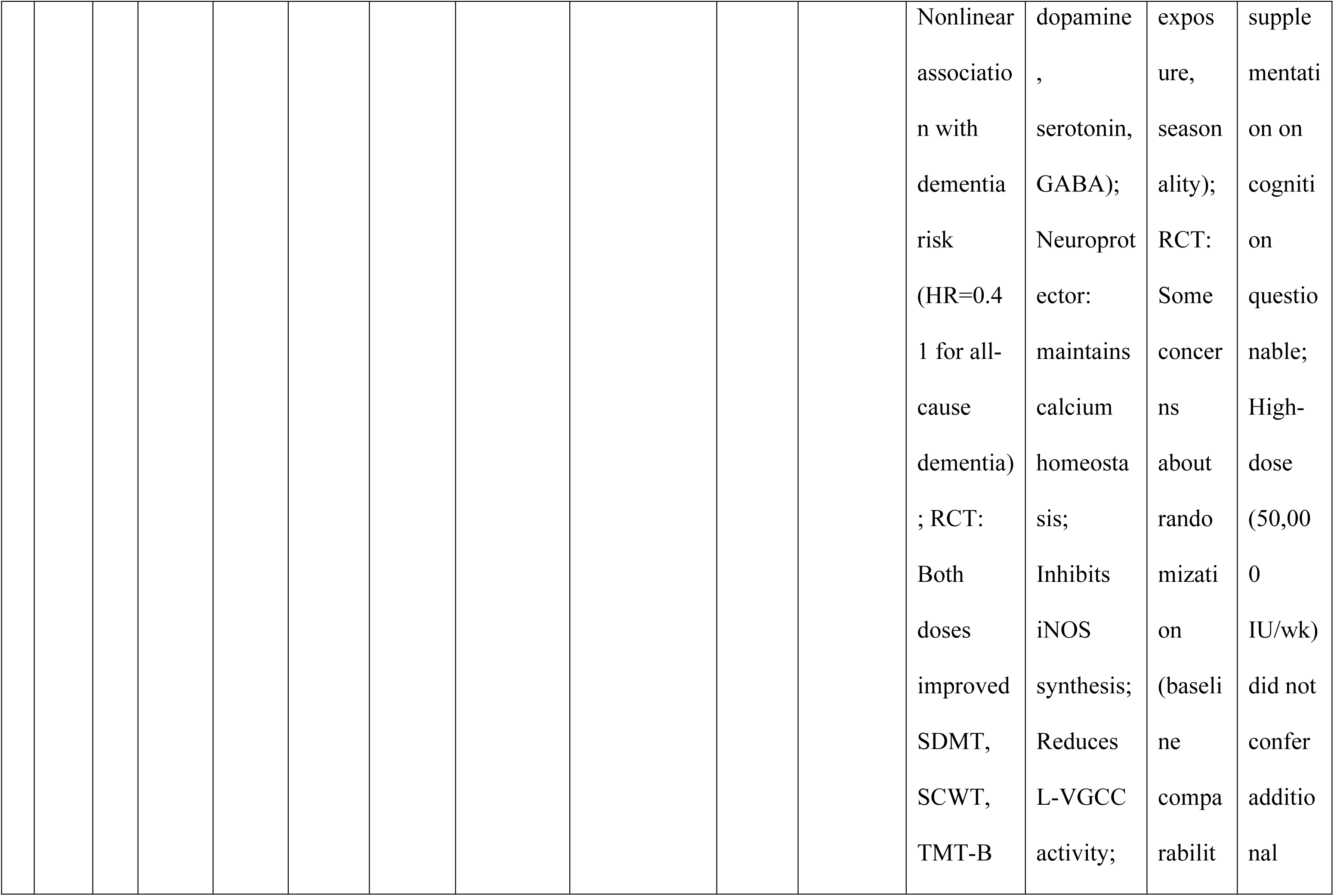
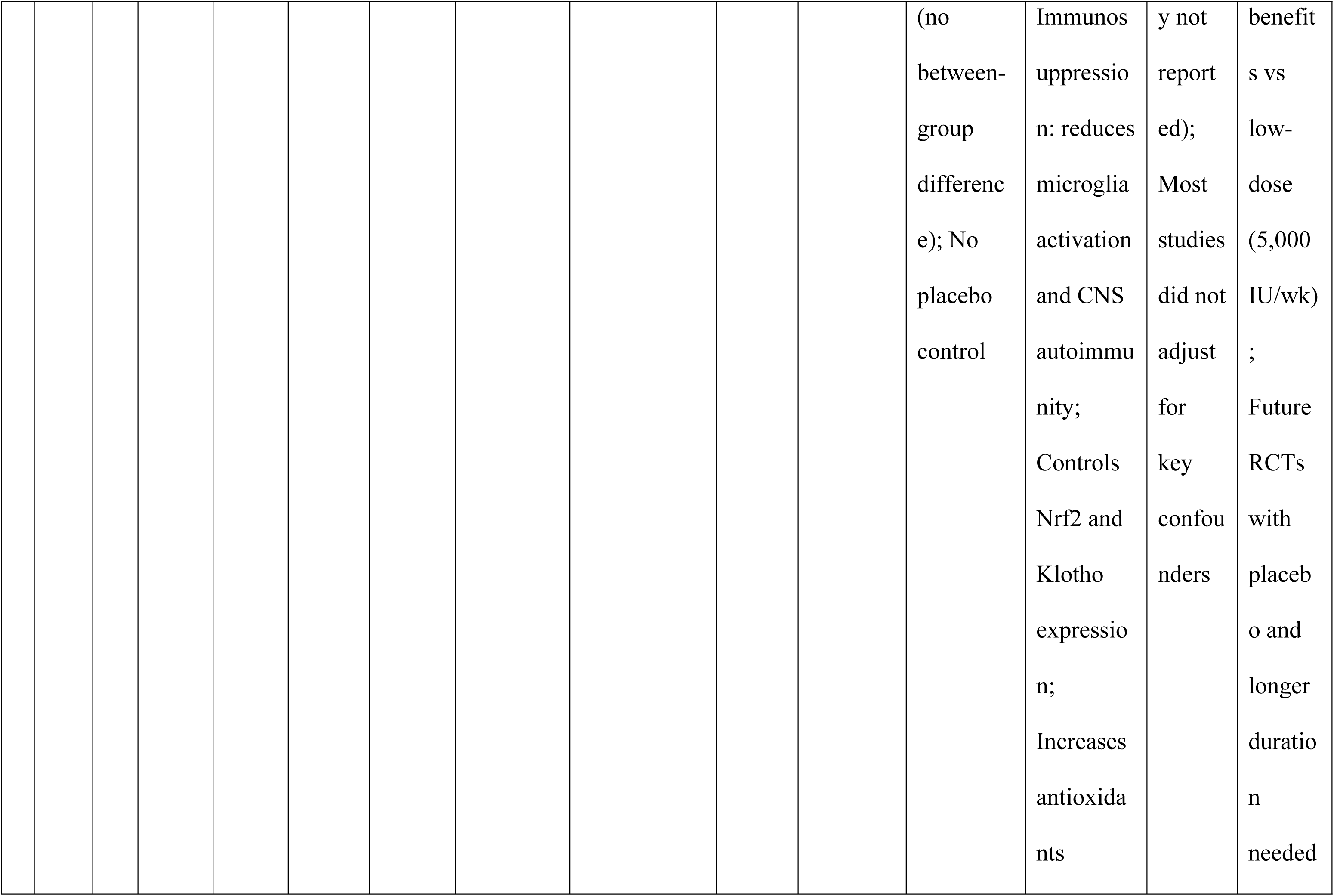
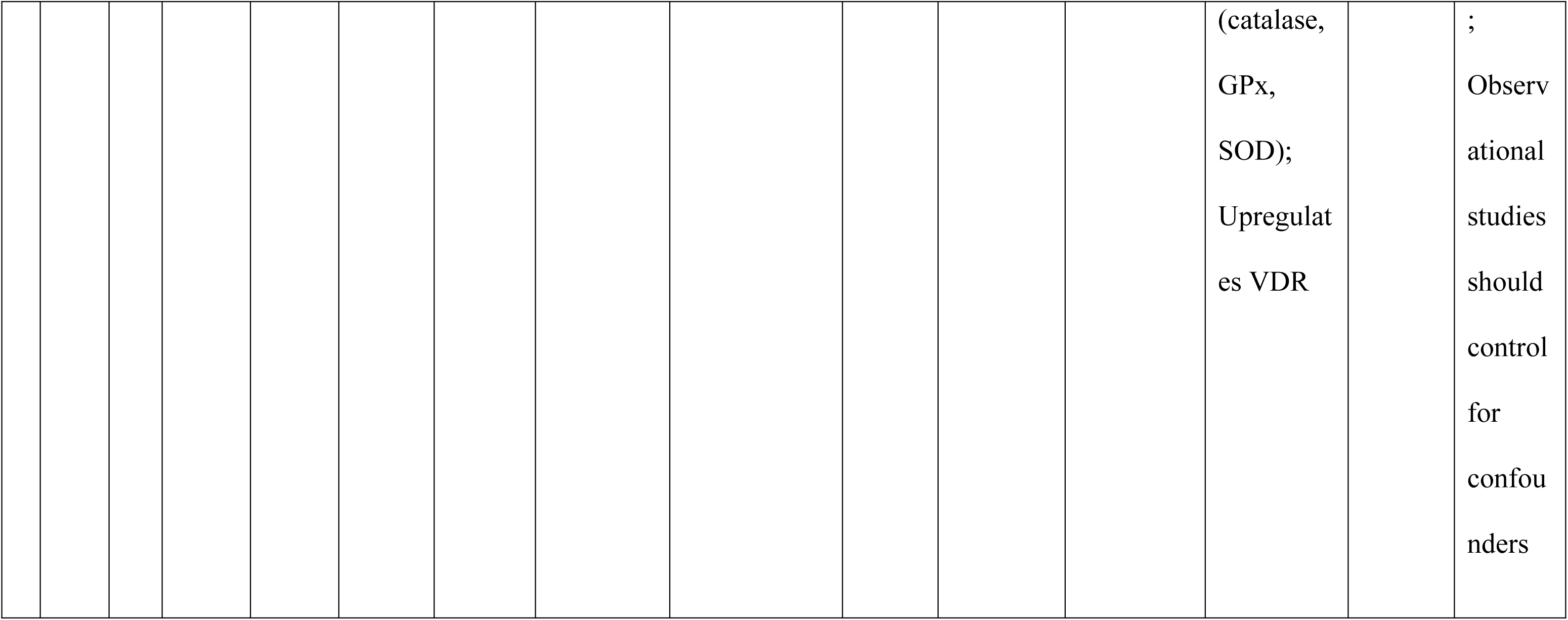
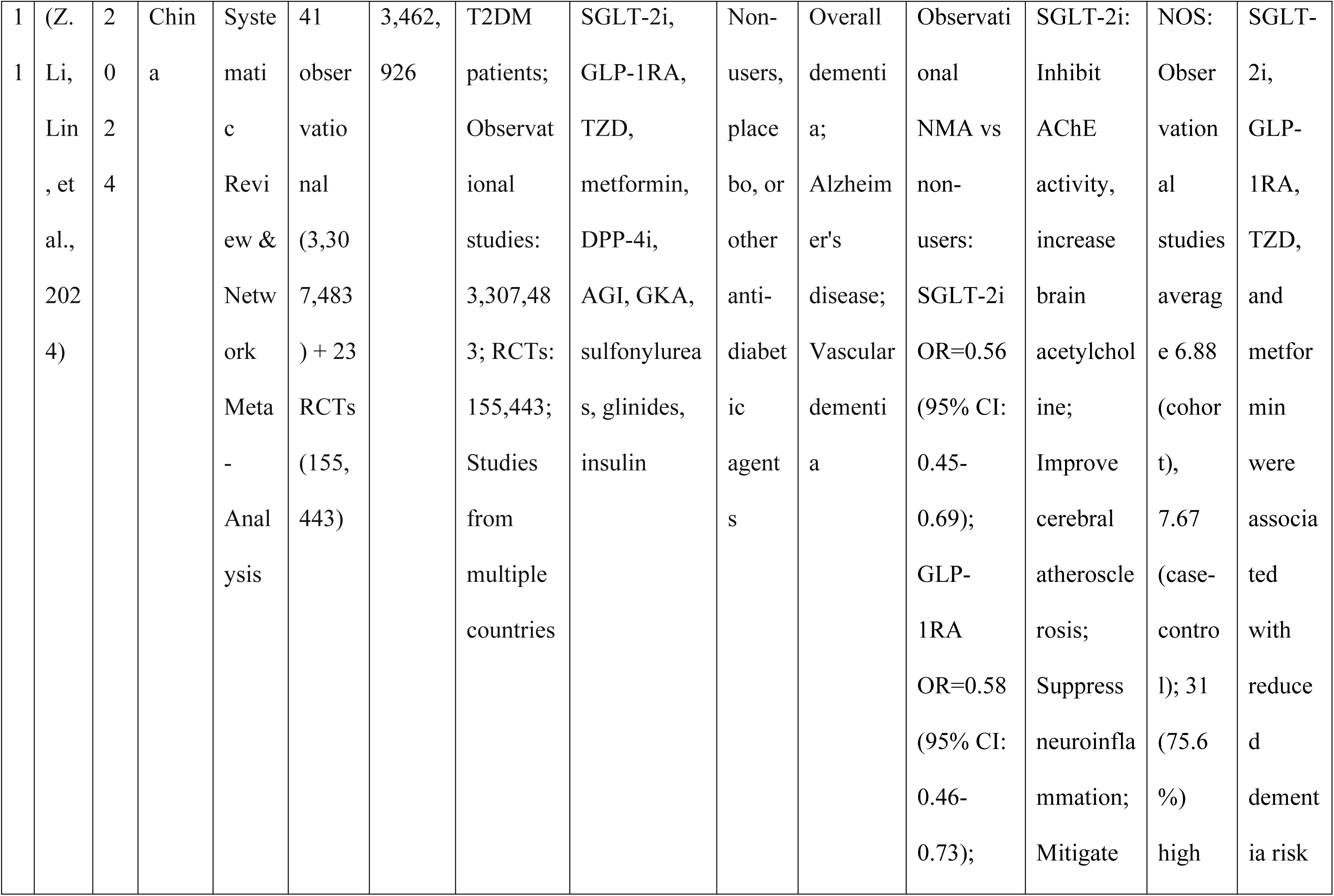
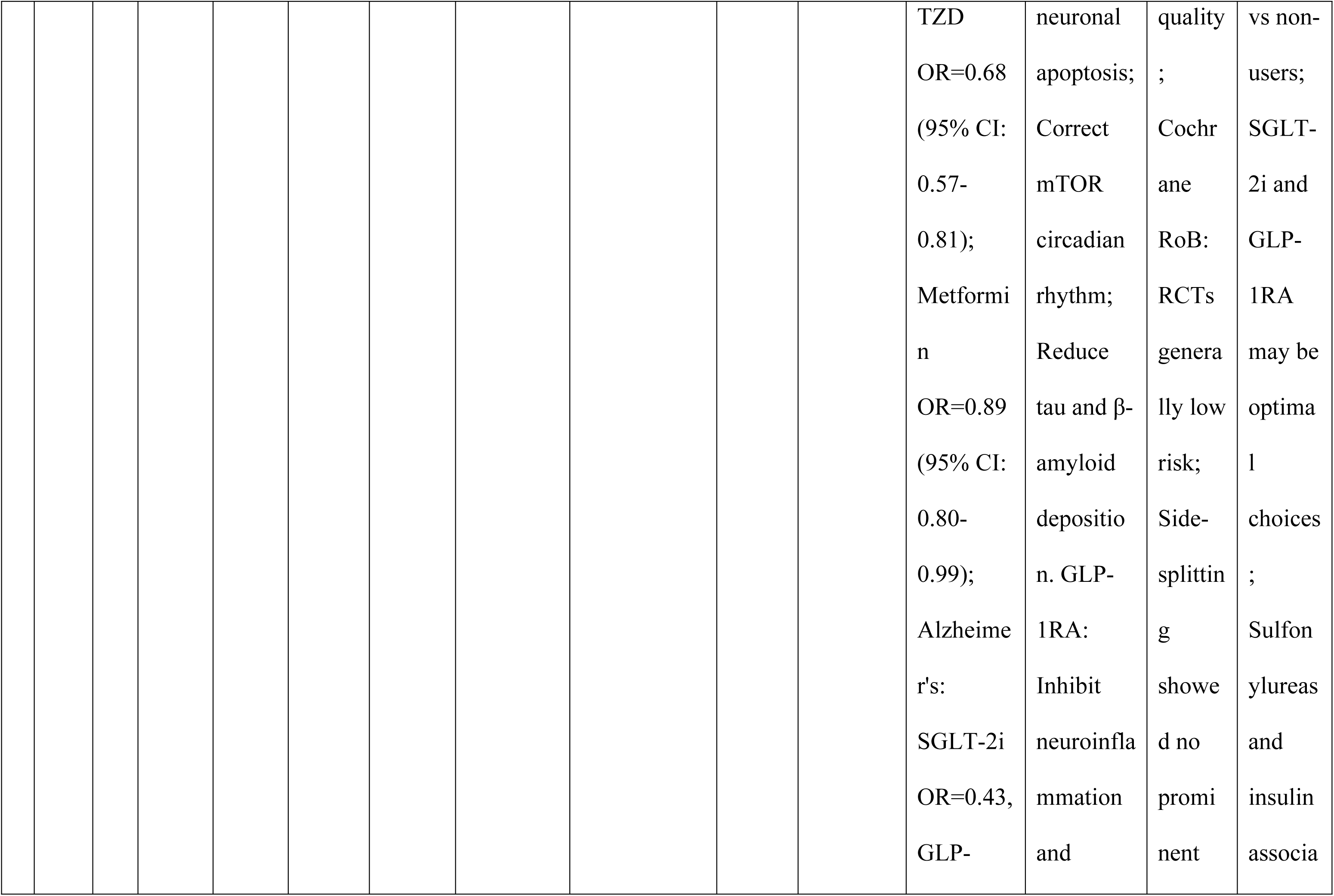
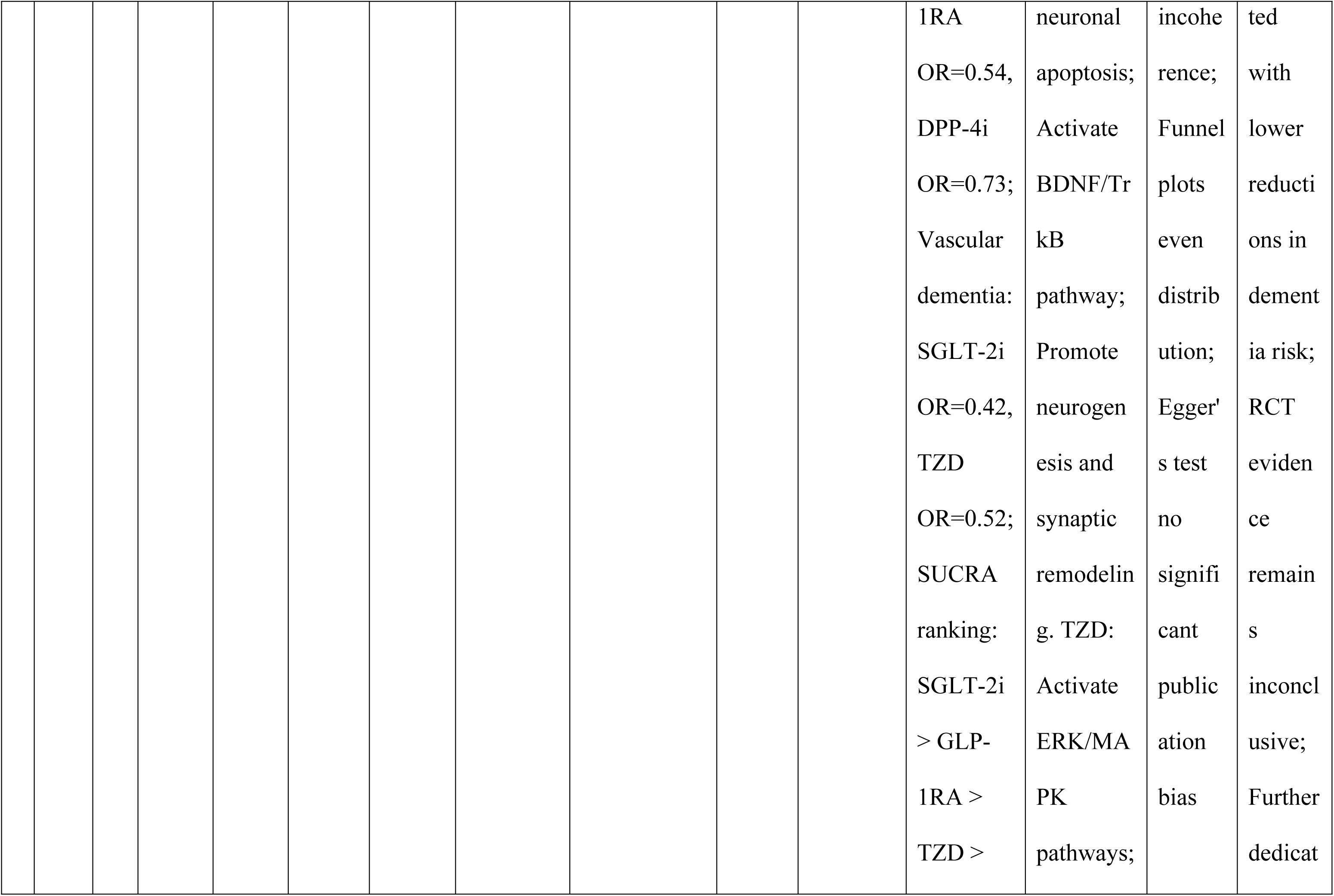
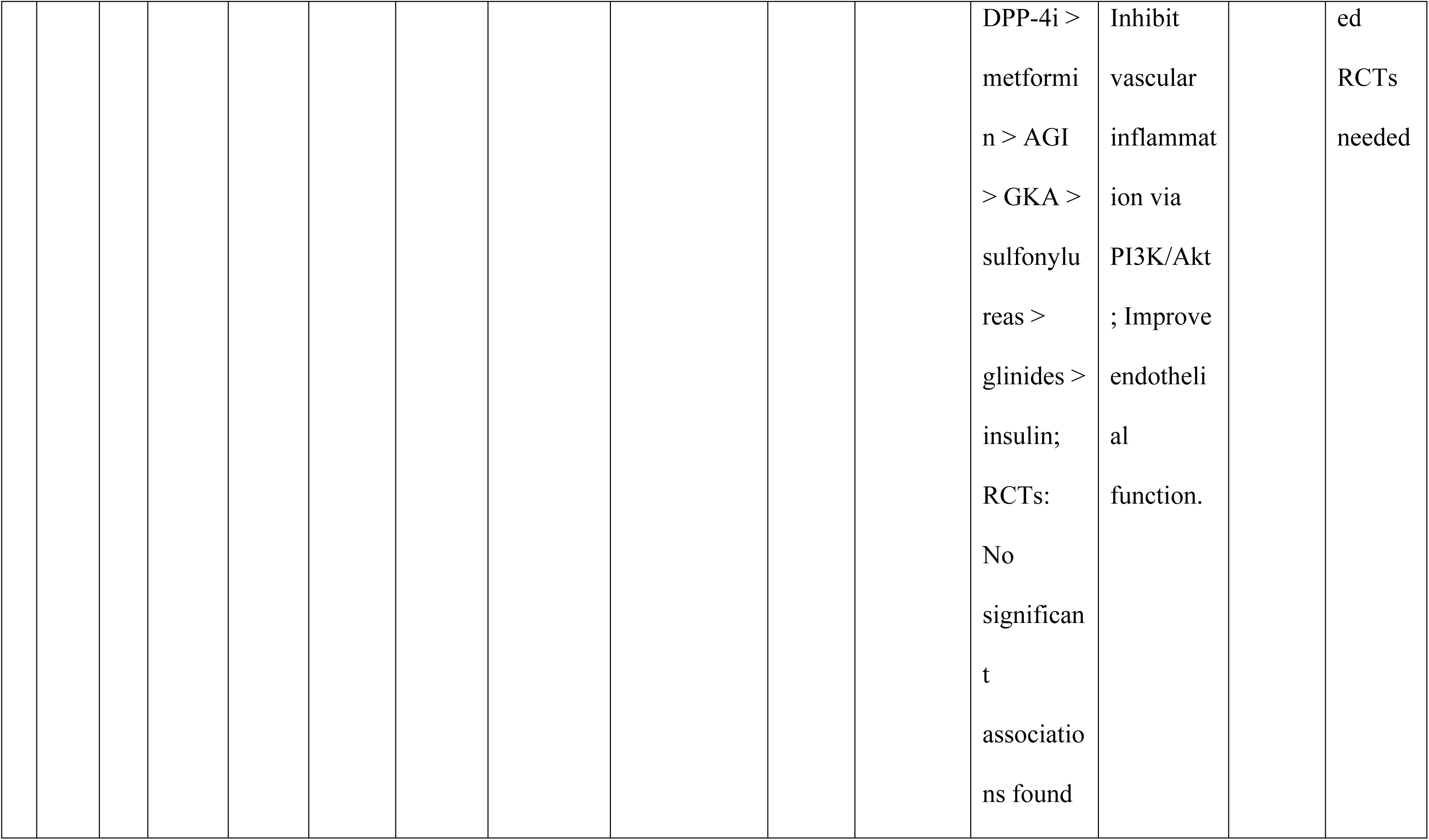
Characteristics of included studies.

We used AMSTAR 2 (Table 2) for quality assessment and three grades were revealed; three reviews achieved high confidence ratings, five achieved moderate confidence, and four were rated as critically low confidence. High-confidence reviews were characterized by prospective protocol registration, comprehensive literature searches, duplicate independent study selection and data extraction, appropriate risk of bias assessment tools, and formal GRADE assessments. Those with critically low confidence ratings lacked protocol registration, failed to report GRADE assessments, had high risk of bias due to inadequate allocation concealment and blinding, and demonstrated publication bias. There was a considerable variation in GRADE certainty across reviews: moderate to high for antidiabetic medications, low to moderate for exercise interventions, very low for Chinese herbal medicine, and very low for vitamin D supplementation. Dementia incidence, MMSE, MoCA, executive function tests, and cognitive domain-specific measures were the primary outcomes assessed. Effect sizes have been reported as odds ratios, risk ratios, standardized mean differences, or mean differences depending on the outcome and study design.

**Table 2:**
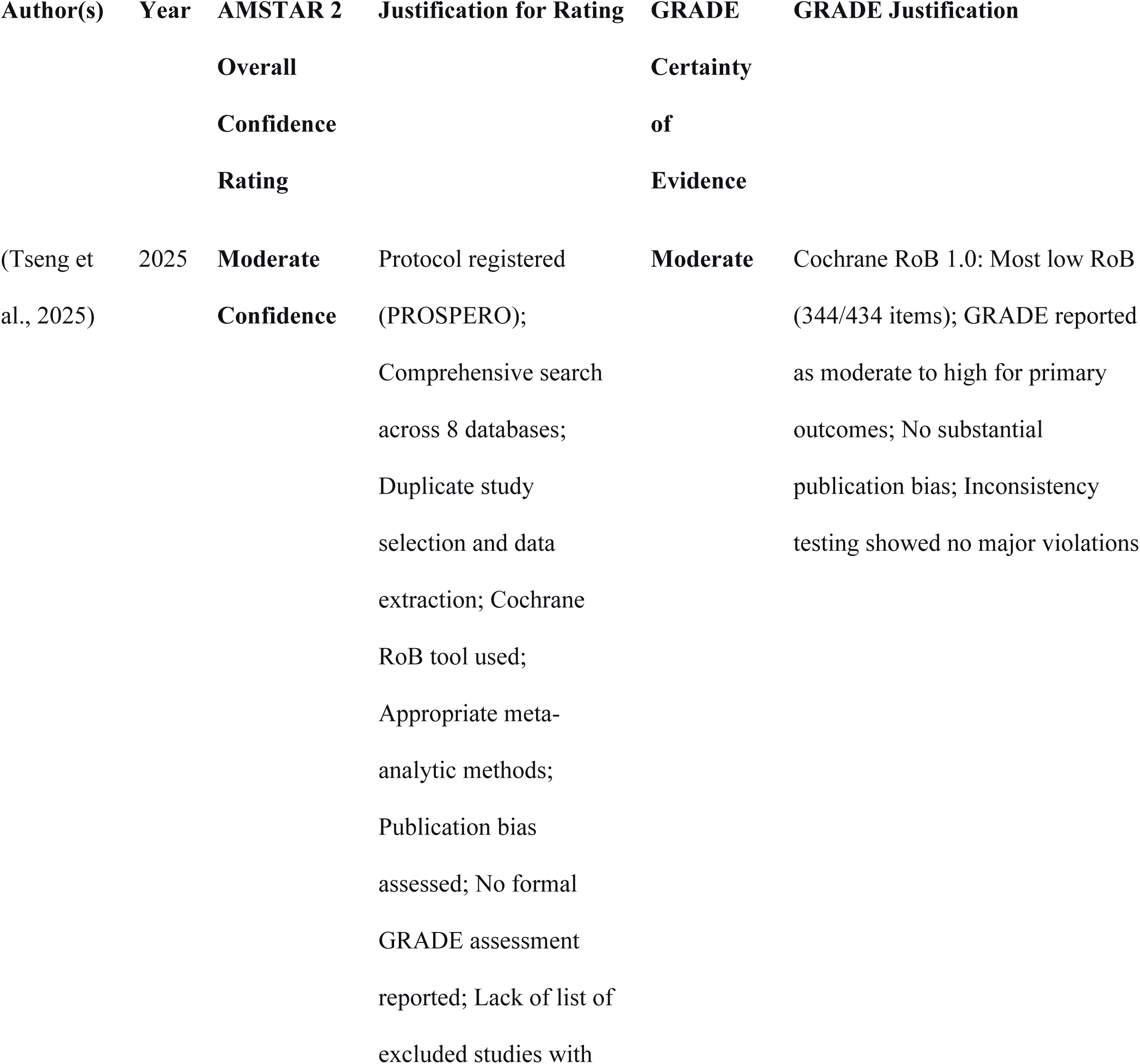
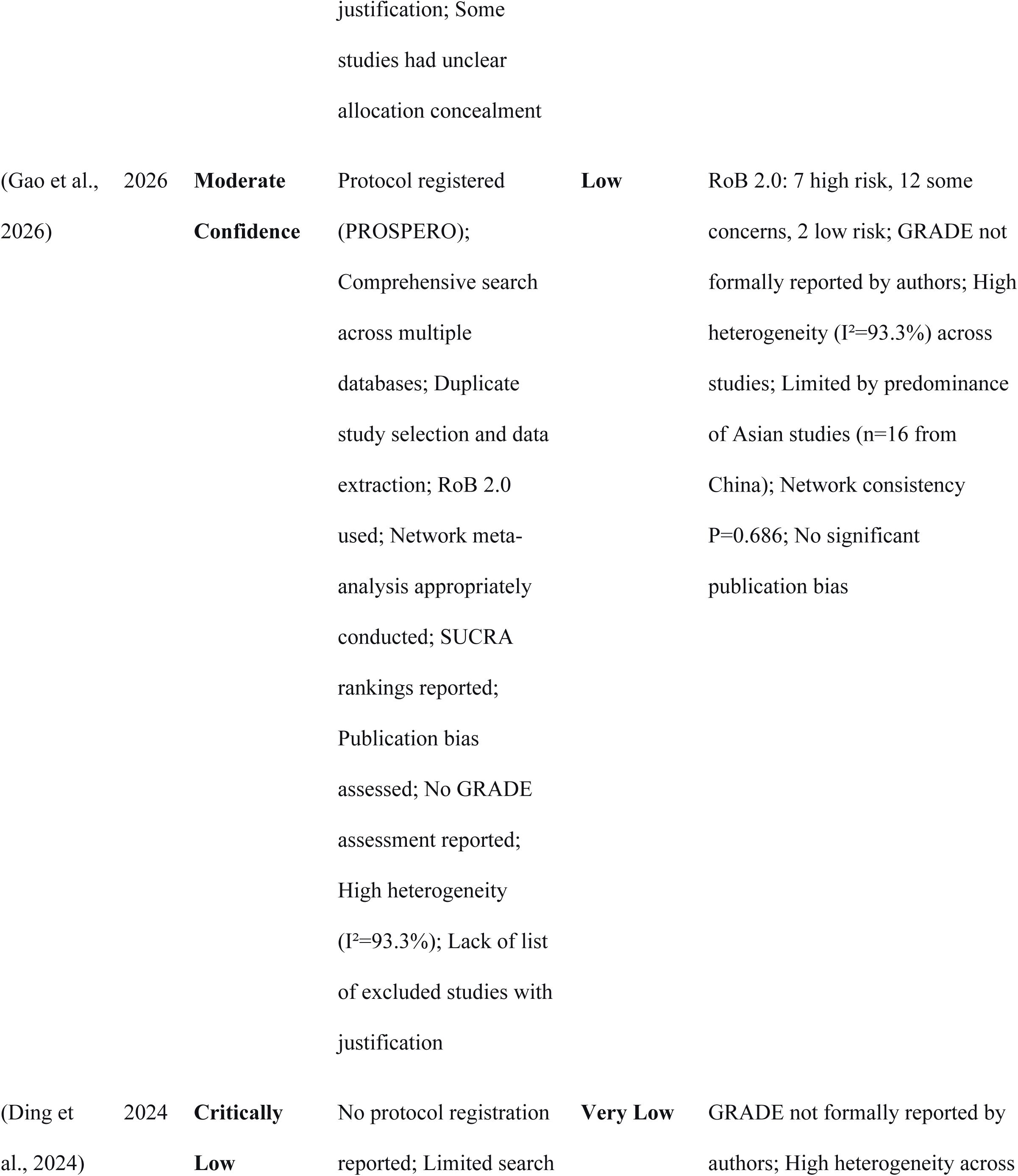
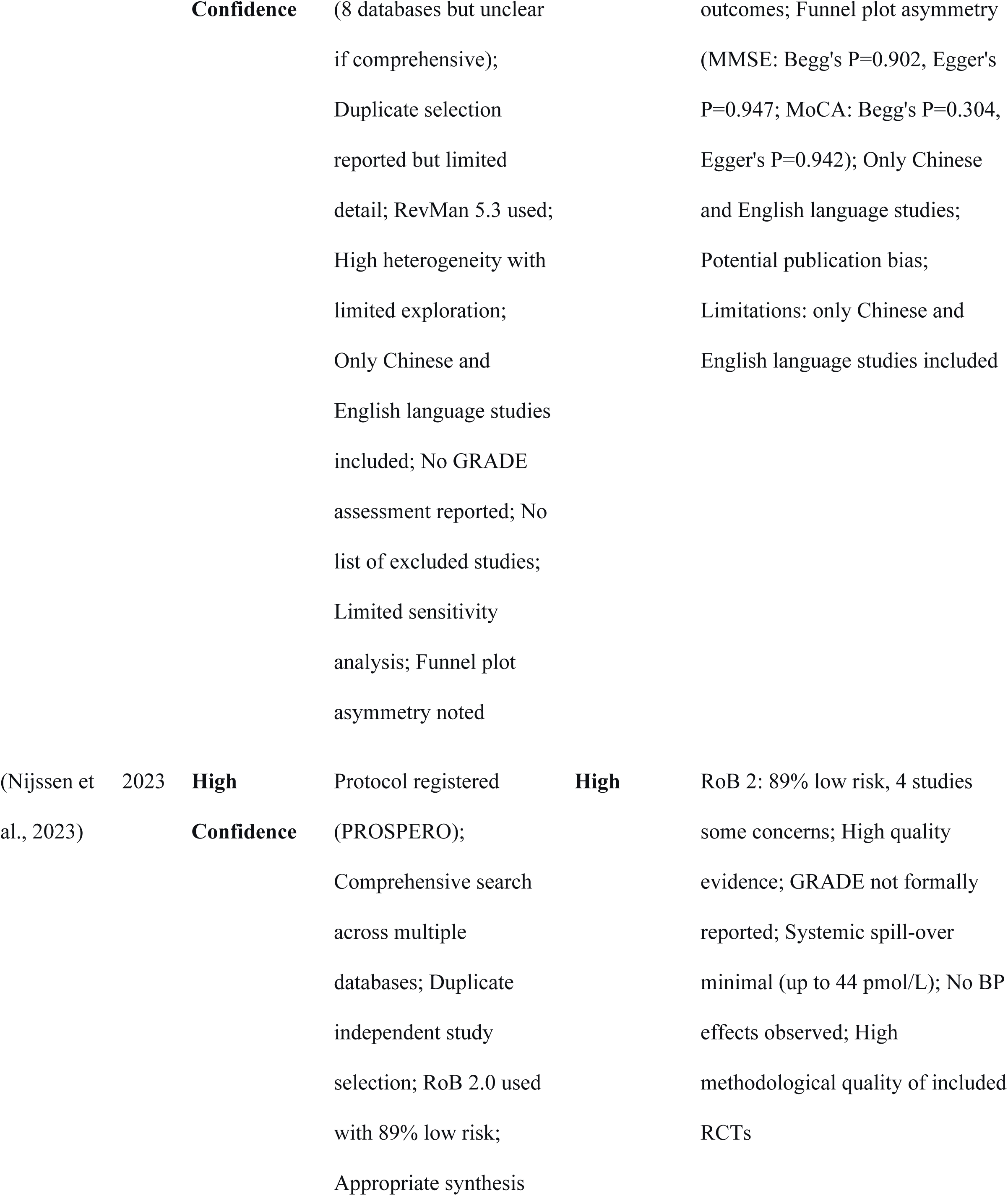
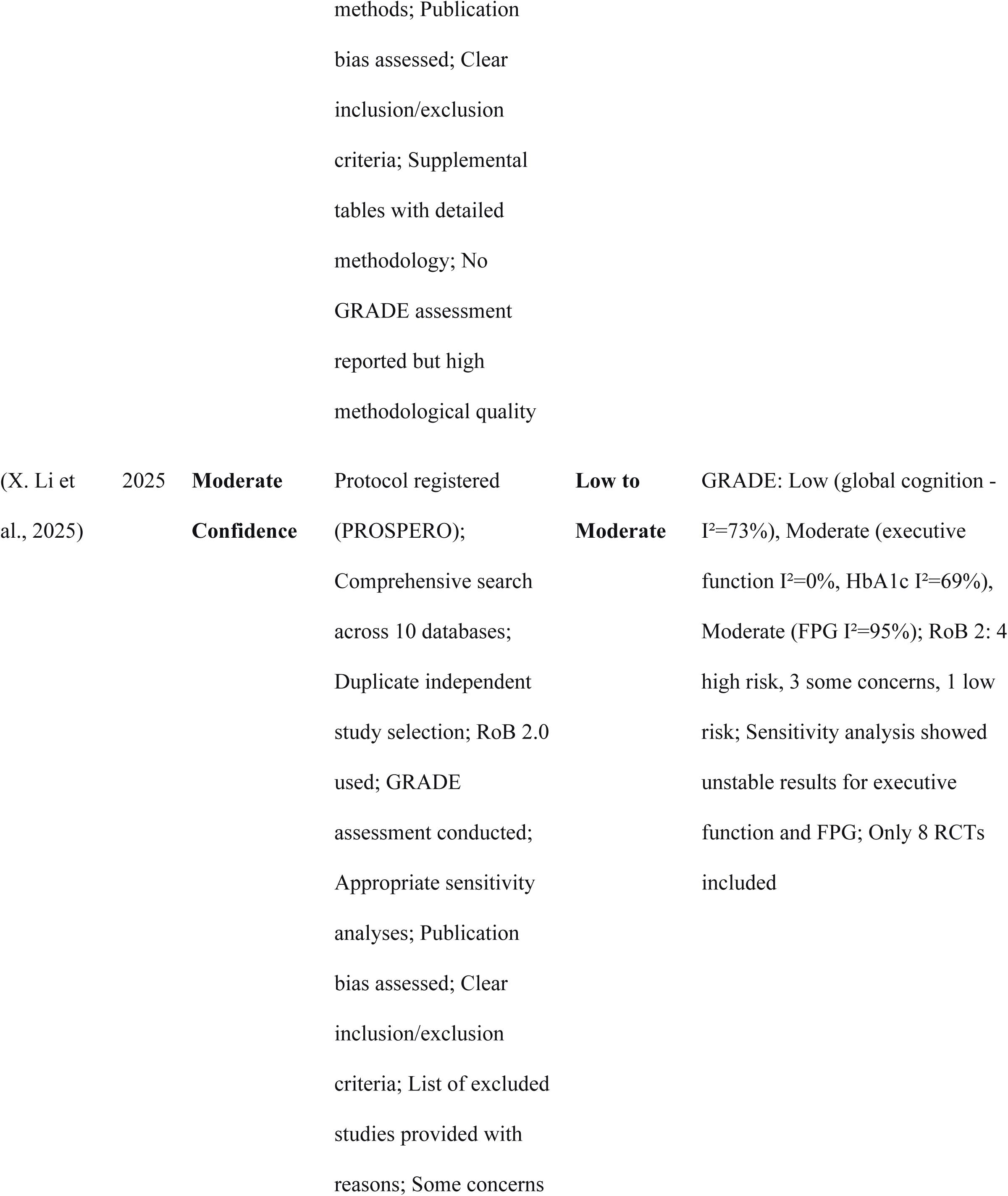
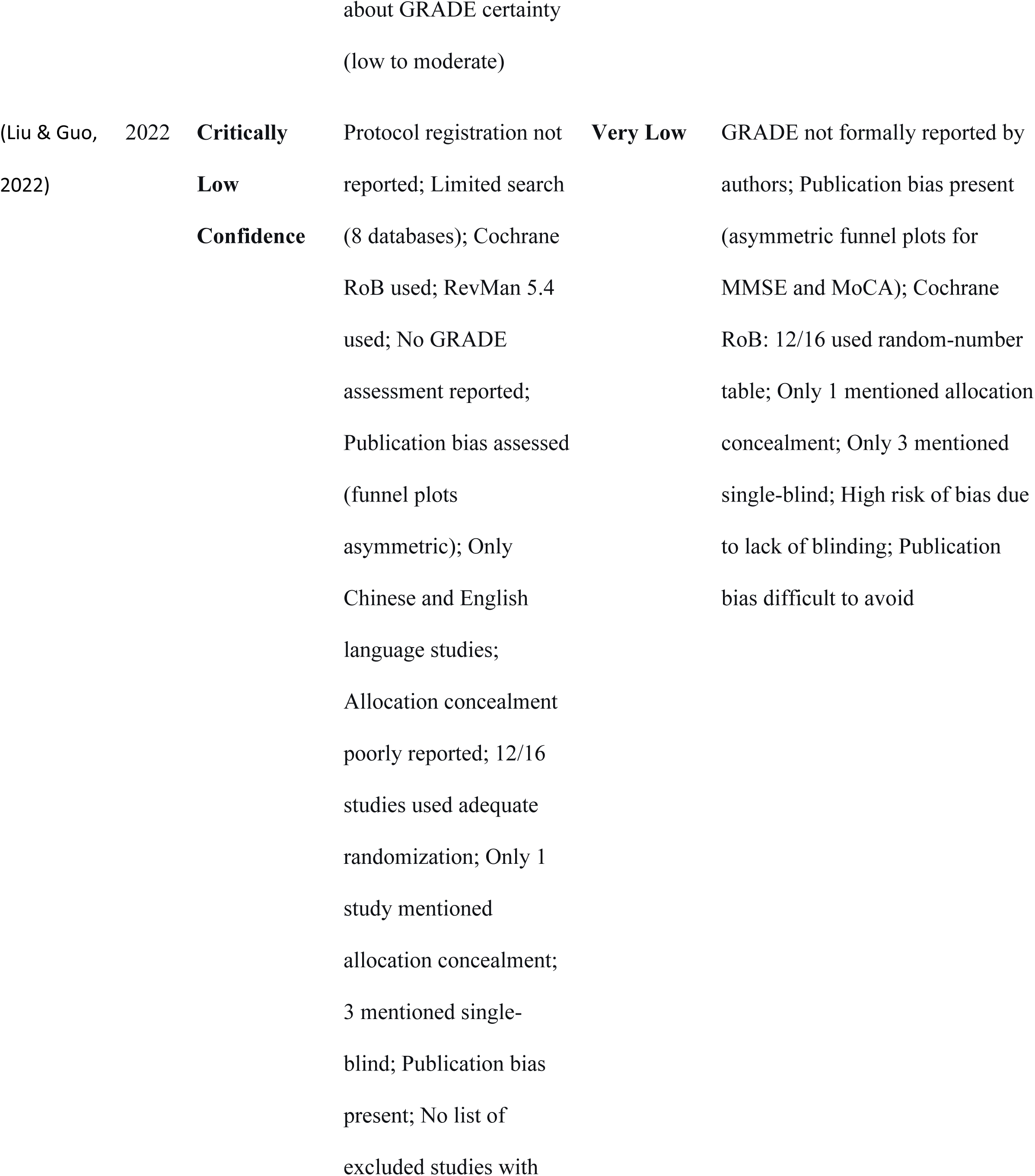
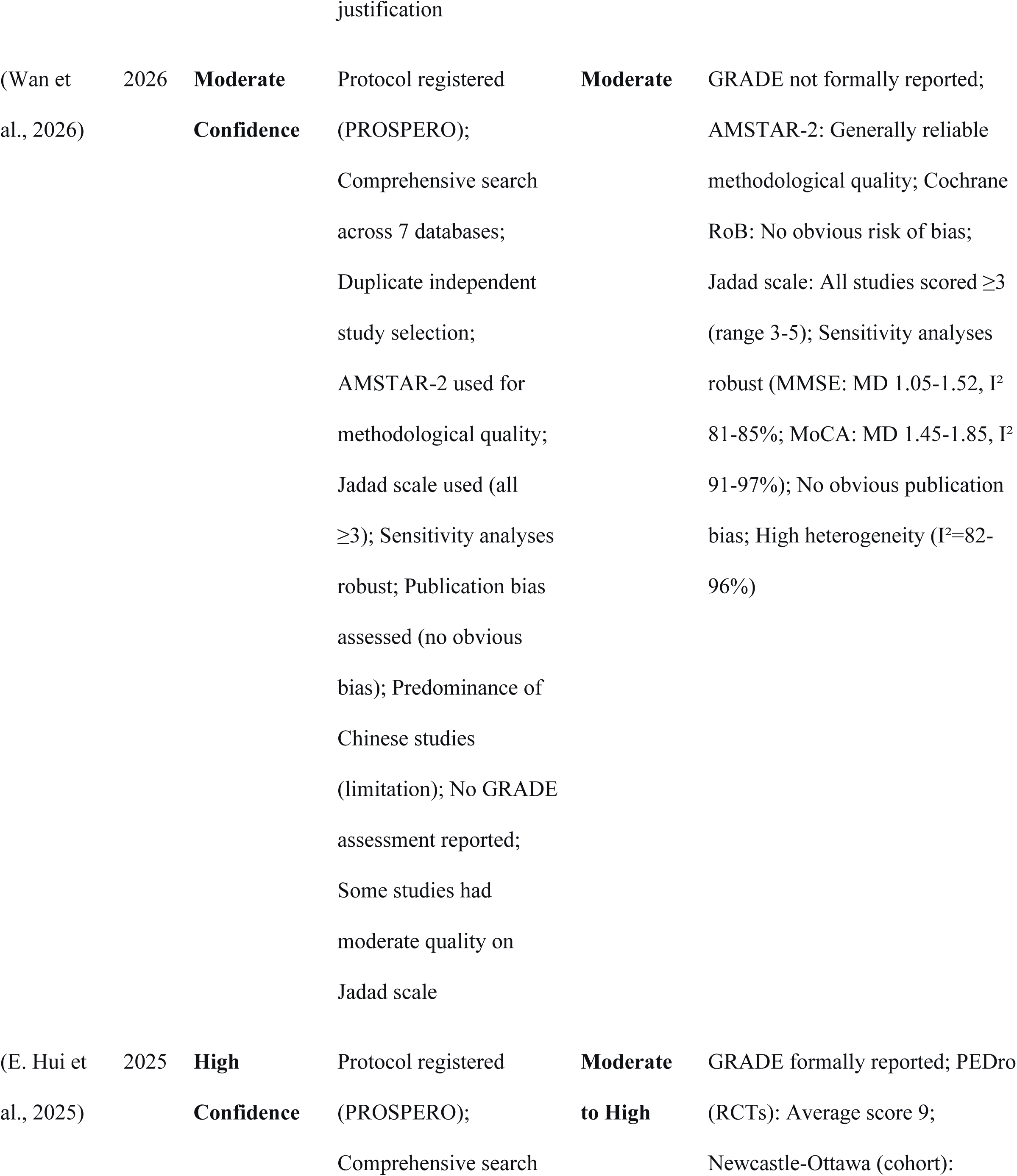
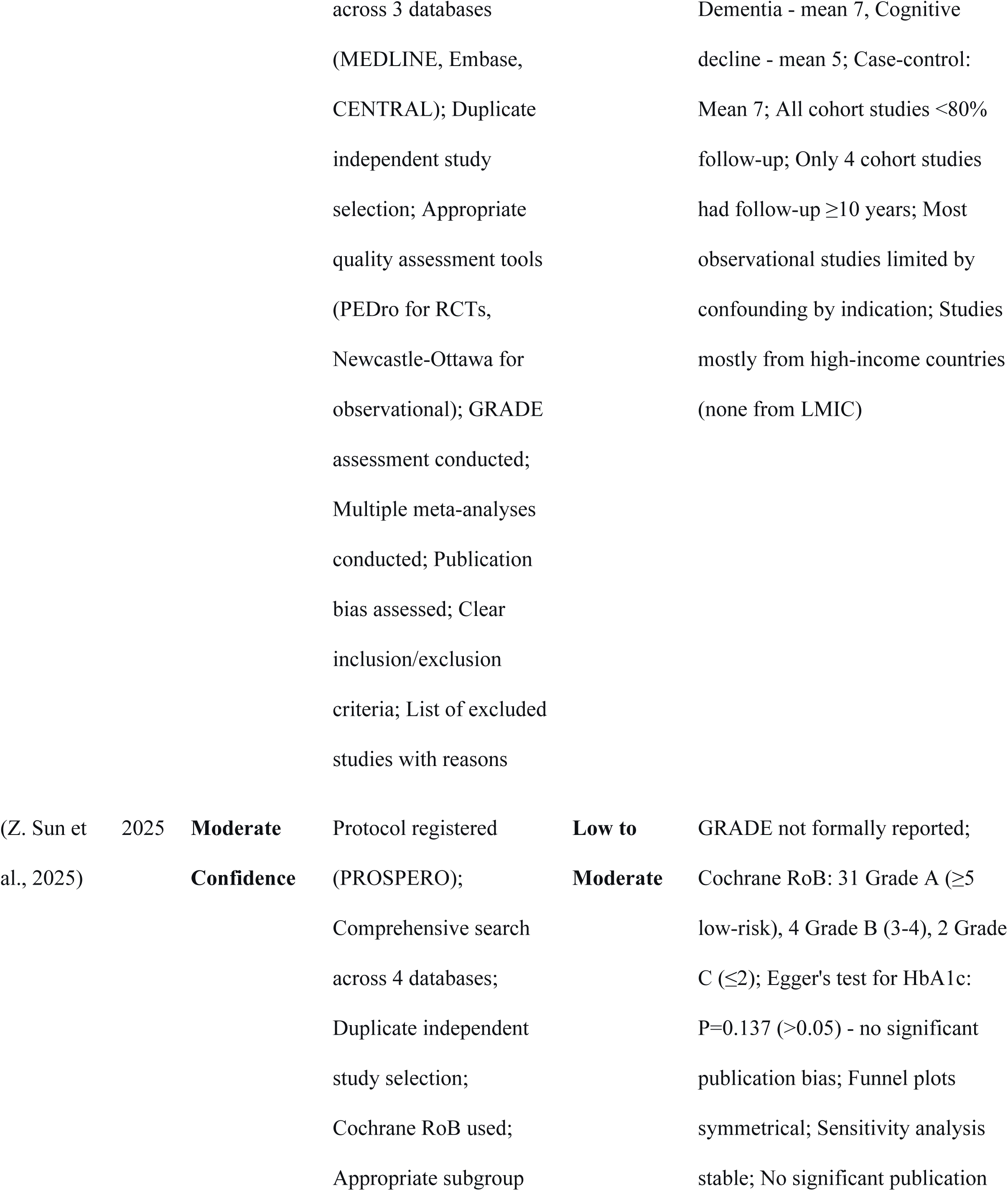
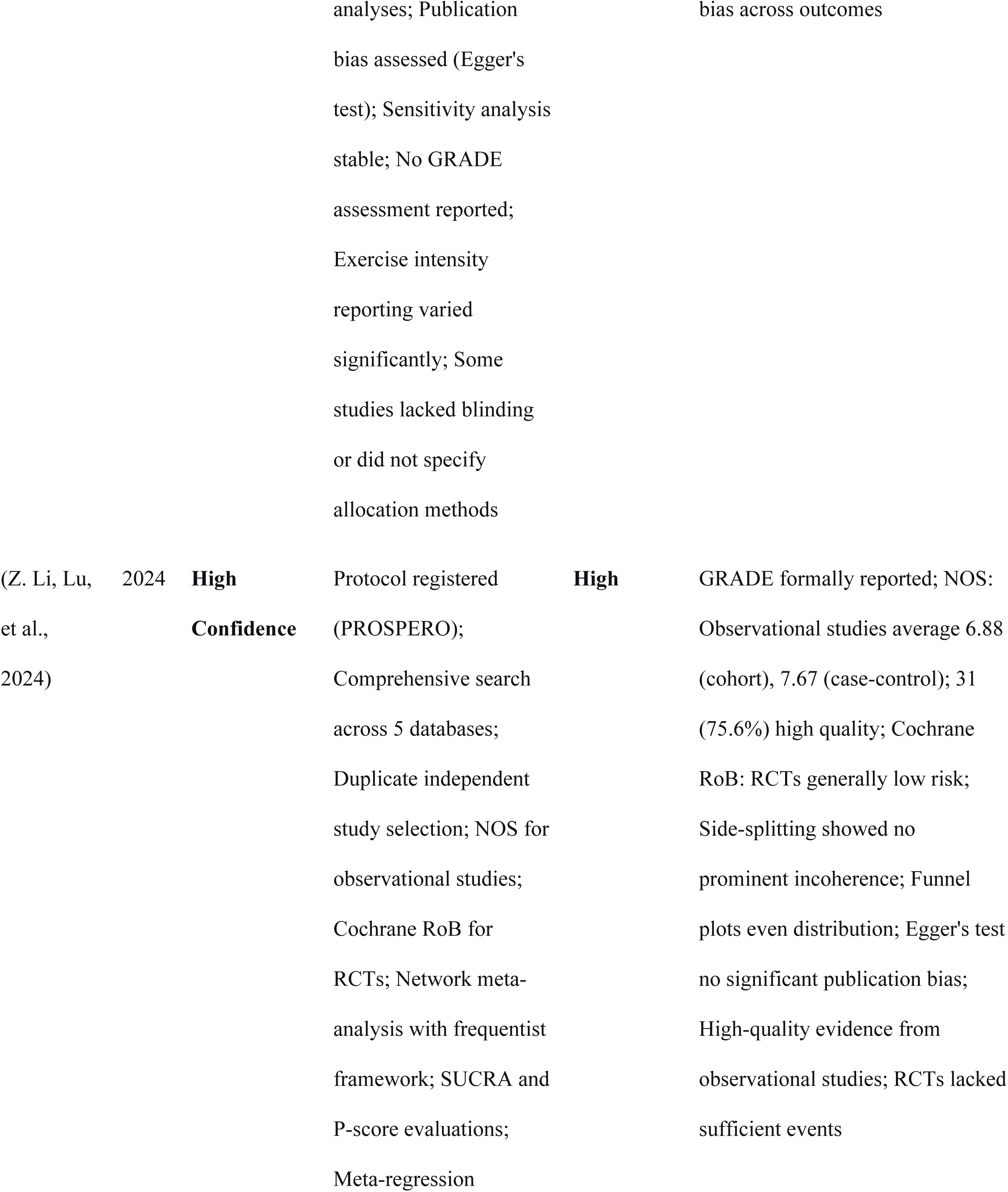
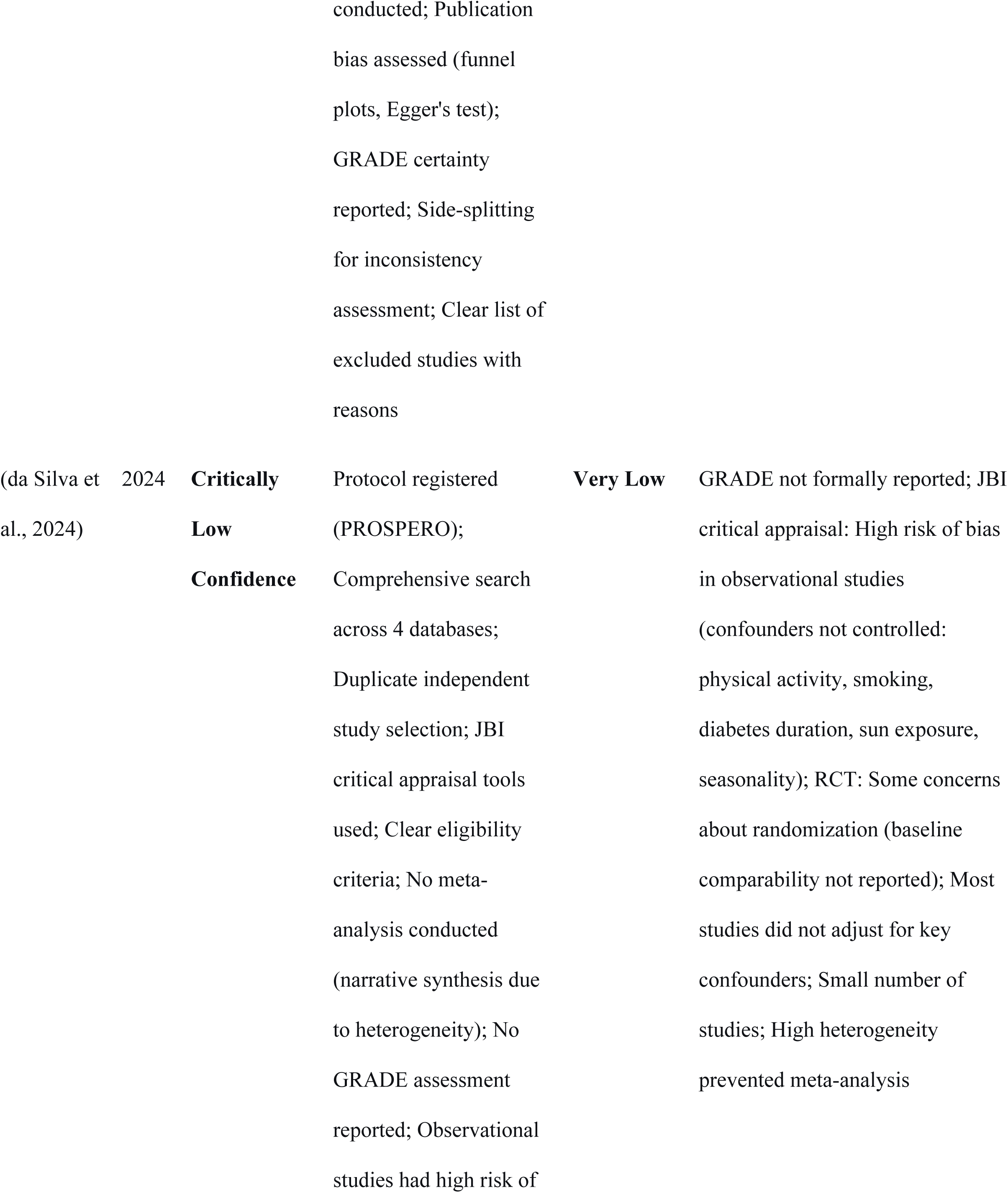
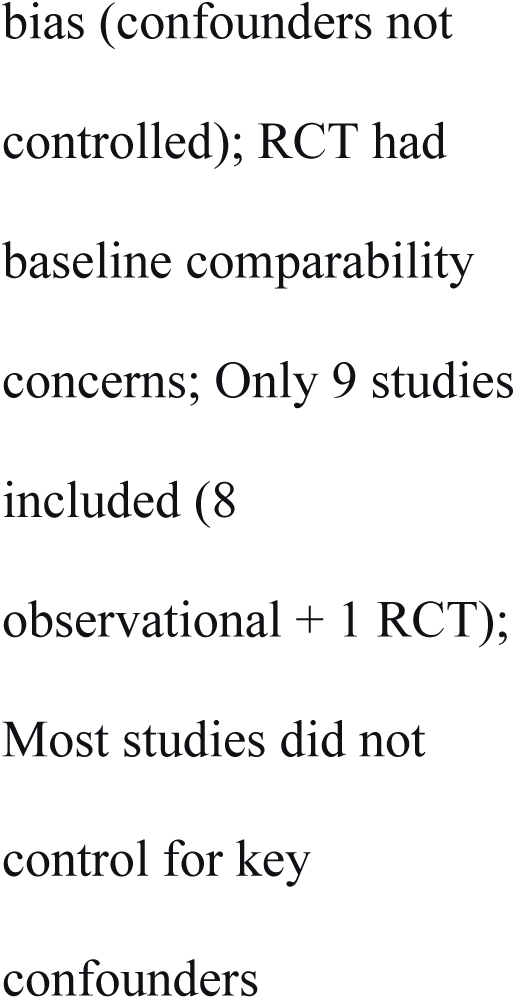
Assessment of methodologic quality.

Table 3 summarizes GRADE certainty ratings for key outcomes across all included reviews. Where reviews reported GRADE, ratings were extracted directly; otherwise, two reviewers independently applied GRADE principles. Discrepancies were resolved by consensus with a third reviewer. Ratings ranged from very low to high certainty across outcomes and reviews. Certainty was downgraded for risk of bias, inconsistency, indirectness, imprecision, and publication bias. High certainty indicates strong confidence in the effect estimate.

**Table 3:**
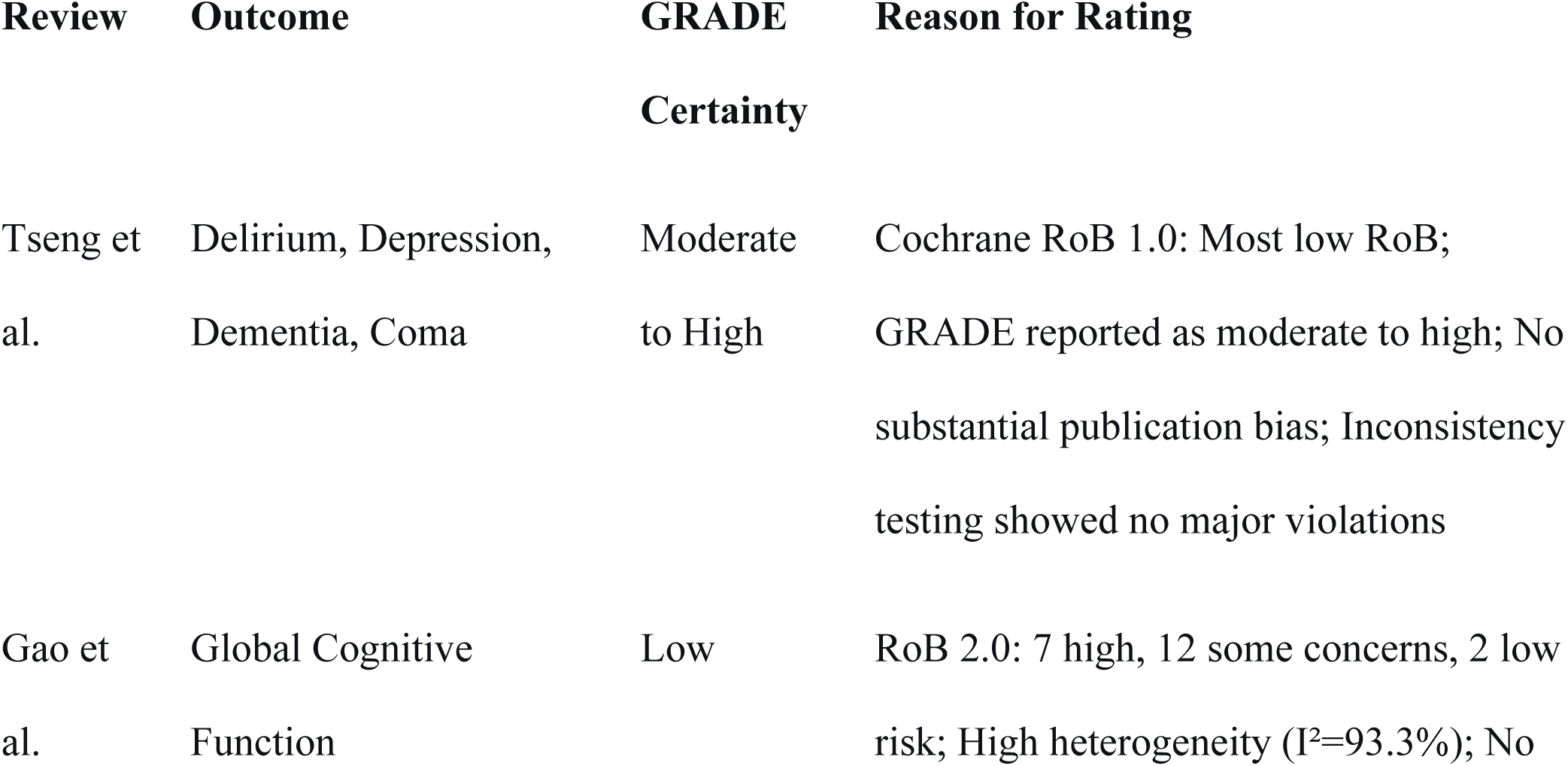
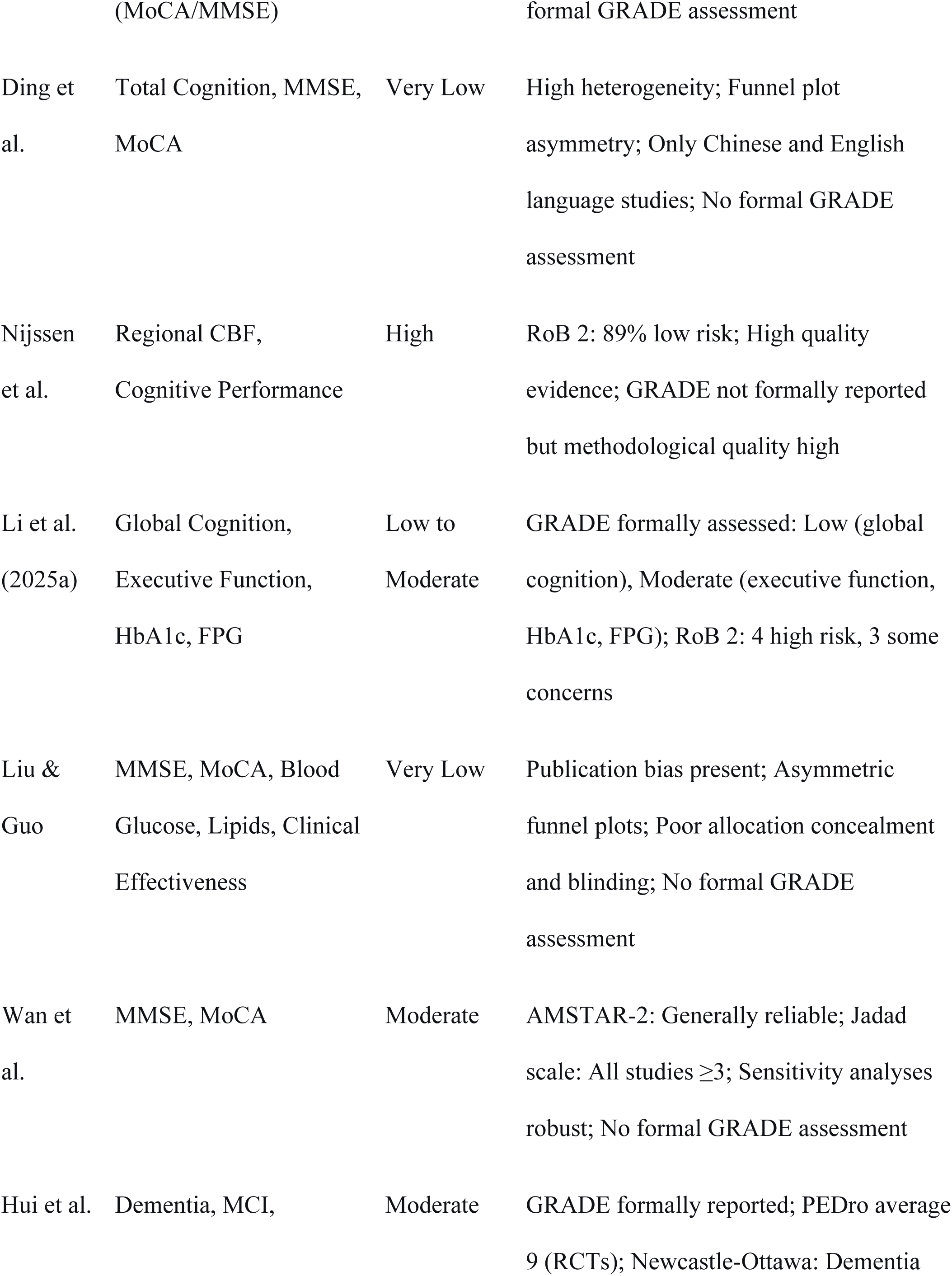
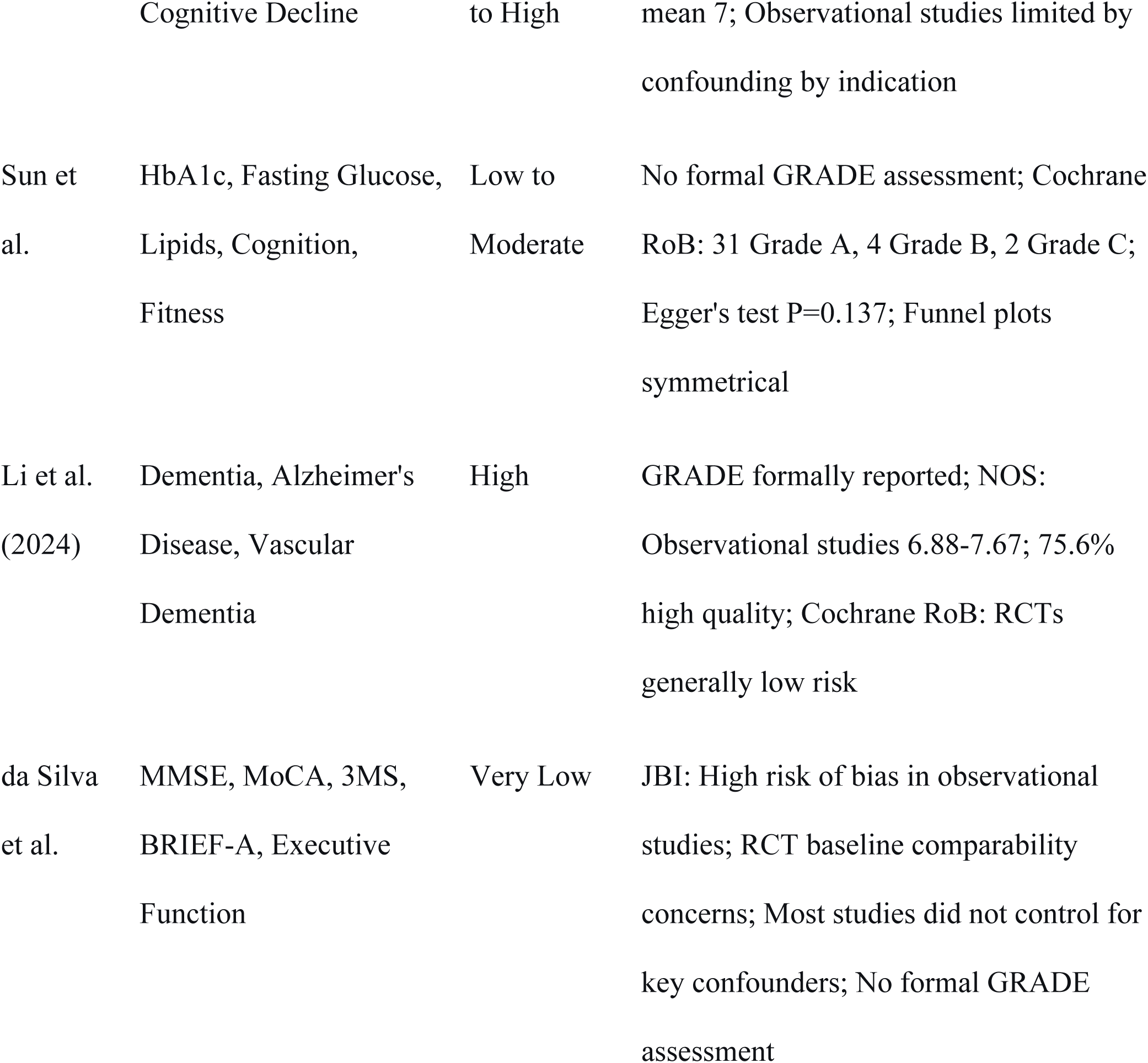
GRADE certainty.

### Effectiveness of pharmacological and non-pharmacological interventions on cognitive outcomes among adults with T2DM

Across all intervention categories (Table 4), antidiabetic medications demonstrated the strongest and most consistent evidence for cognitive protection. GLP-1 receptor agonists reduced dementia risk by 47–53% in randomized controlled trials (RR = 0.47, 95% CI: 0.25–0.86) and improved MMSE scores (WMD = 1.33, 95% CI: 0.67–1.99) and MoCA scores (WMD = 1.70, 95% CI: 0.83–2.56). Long-term use (≥24 weeks) conferred significantly greater benefits (MD = 3.74, p < 0.0001). SGLT2 inhibitors were ranked as the most effective agents for reducing dementia risk (OR = 0.56, 95% CI: 0.45–0.69; SUCRA = 94.5%), with high-dose dapagliflozin specifically reducing delirium (OR = 0.21) and depression (OR = 0.43). Thiazolidinediones demonstrated consistent protective effects with no heterogeneity (RR = 0.78, I² = 0%), while metformin, sulfonylureas, insulin, and DPP-4 inhibitors yielded inconsistent findings, largely limited by confounding by indication and high heterogeneity.

**Table 4.**
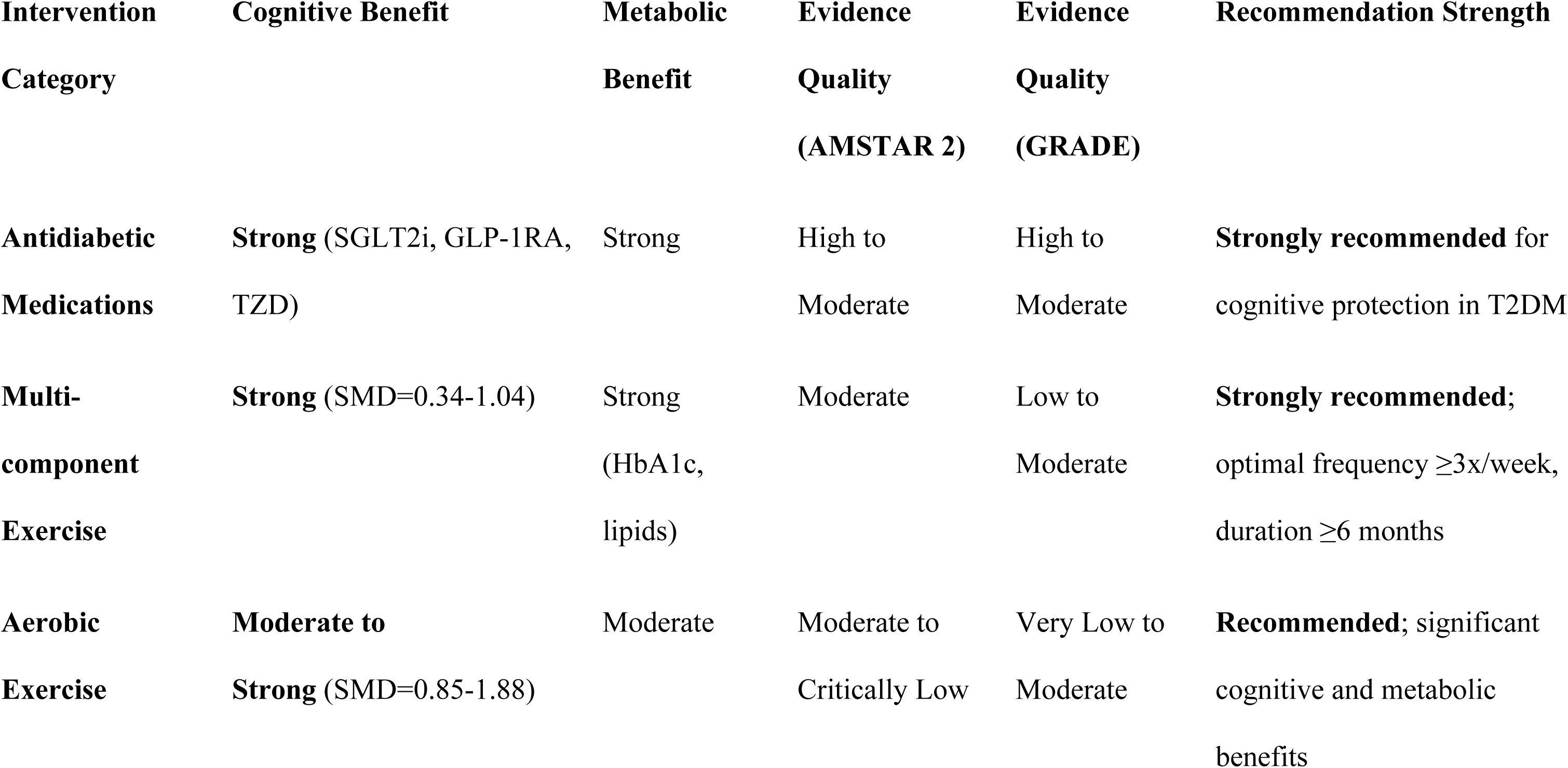
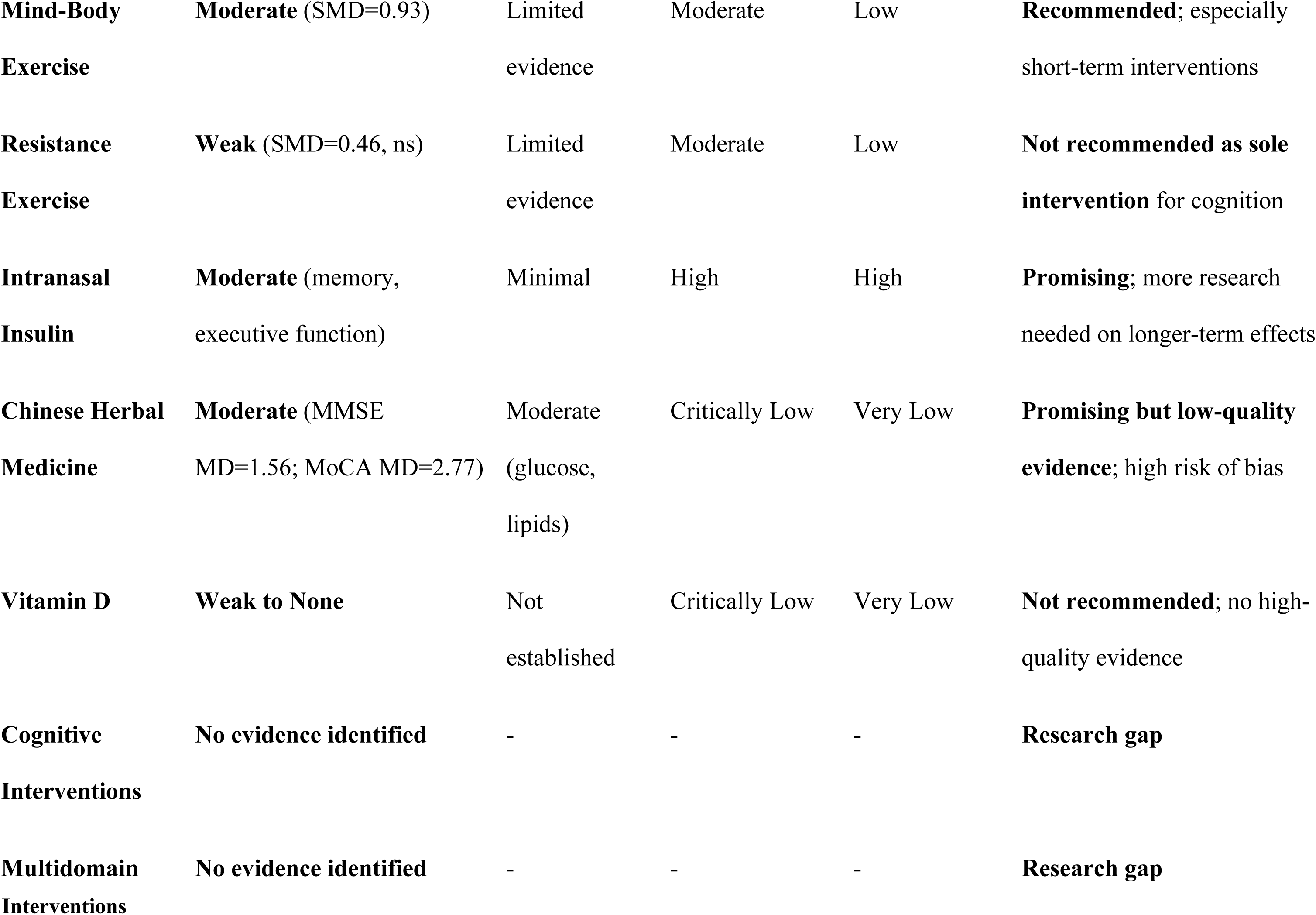
Effectiveness of pharmacological and non-pharmacological interventions on cognitive outcomes among adults with T2DM.

Lifestyle interventions, particularly exercise, also demonstrated significant cognitive benefits. Multi-component exercise was the most effective exercise modality (SMD = 1.04, 95% CI: 0.49– 1.59; SUCRA = 78.1%), followed by mind-body exercise (SMD = 0.93) and aerobic exercise (SMD = 0.85), while resistance exercise showed no significant effect (SMD = 0.46). Optimal benefits were achieved with exercise frequency of at least three sessions per week and intervention duration of six months or longer. Multi-component exercise also significantly improved glycemic control (HbA1c: SMD = -0.52; fasting glucose: SMD = -0.53), lipid metabolism, physical fitness, and quality of life. Intranasal insulin demonstrated region-specific cerebral blood flow changes and improved memory and executive function, with effects modified by obesity, T2D, and aging (AMSTAR 2: High confidence). Chinese herbal medicine as adjuvant therapy improved clinical effectiveness (OR = 5.33), MMSE (MD = 1.56), and MoCA (MD = 2.77) while reducing adverse events, though evidence quality was critically low due to poor allocation concealment and publication bias. Vitamin D supplementation showed no high-quality evidence for cognitive benefit, with high-dose (50,000 IU/week) conferring no additional benefit over low-dose (5,000 IU/week).

### Assessment of overlap

The 11 systematic reviews included in this overview demonstrated substantial heterogeneity in their primary study inclusion, ranging from 8 to 62 RCTs per review. To quantify the degree of overlap among primary studies across reviews, the Corrected Covered Area (CCA) method was applied. Overall CCA of 2.11% indicates slight overlap across all 11 reviews, suggesting that the included reviews largely draw from distinct primary study pools with minimal duplication.

### Metanalysis of pooled effect estimates

Table 6, summarizes a meta-analysis of interventions for dementia risk and cognitive function. It findings have been grouped into three categories: antidiabetic drugs, lifestyle changes, and nutritional supplements. The pooled effect estimates were derived from random effects models. These models were chosen due to moderate to high heterogeneity in many subgroups. Effects are presented as risk ratios or odds ratios for dementia incidence. Cognitive performance is presented as mean differences. Heterogeneity was measured using the I² statistic. Evidence quality was assessed with the AMSTAR 2 tool. This tool evaluates the methodological strength of included reviews. Confidence intervals show the precision of each estimate. High I² values (above 75%) in some cases warn of inconsistency. These findings require cautious interpretation.

**Table 5.** Effect Size Comparison Across Intervention Categories.

| **Intervention** | **Cognitive Effect Size (SMD/MD)** | **Comparison to Control** | **Heterogeneity (I<sup>2</sup>)** |
| --- | --- | --- | --- |
| SGLT2 Inhibitors (Dapagliflozin 10mg) | Delirium OR=0.21; Depression OR=0.43 | Significant | Not reported |
| GLP-1 Receptor Agonists | MMSE WMD=1.33; MoCA WMD=1.70 | Significant | 82-96% |
| Thiazolidinediones (TZD) | Dementia RR=0.78; Vascular dementia OR=0.52 | Significant | 0% |
| DPP-4 Inhibitors | Alzheimer's OR=0.73; Dementia RR=0.86 | Mixed | 92.9% |
| Metformin | Dementia OR=0.89 | Mixed | 98.4% |
| Multi-component Exercise | Cognition SMD=0.34-1.04 | Significant | 41-93% |
| Aerobic Exercise | Cognition SMD=0.85-1.88 | Significant | High |
| Mind-Body Exercise | Cognition SMD=0.93 | Significant | Not reported |
| Chinese Herbal Medicine | MMSE MD=1.56; MoCA MD=2.77 | Significant | 25-84% |
| Vitamin D | No significant benefit | Not significant | Not applicable |

**Table 6.**
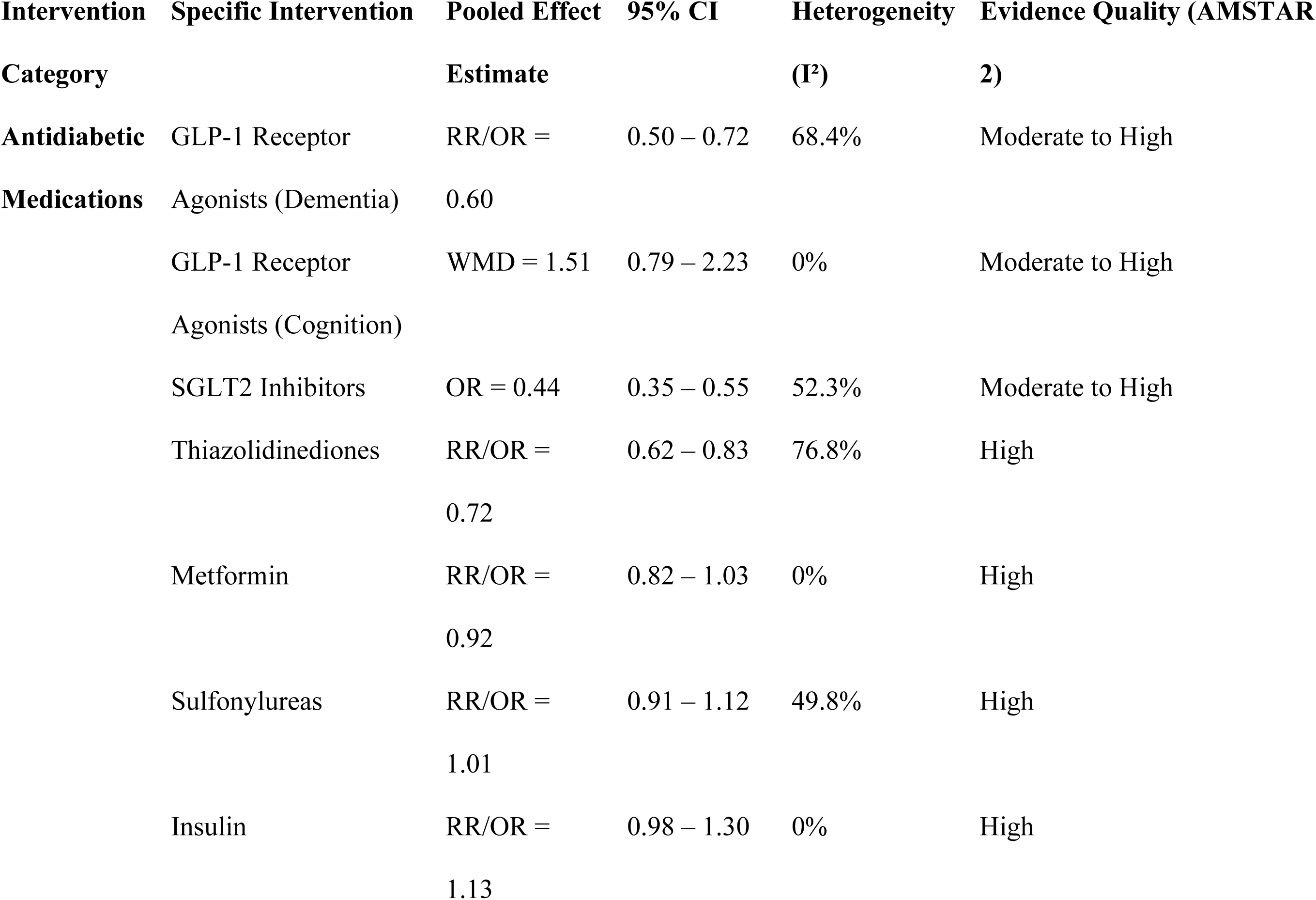
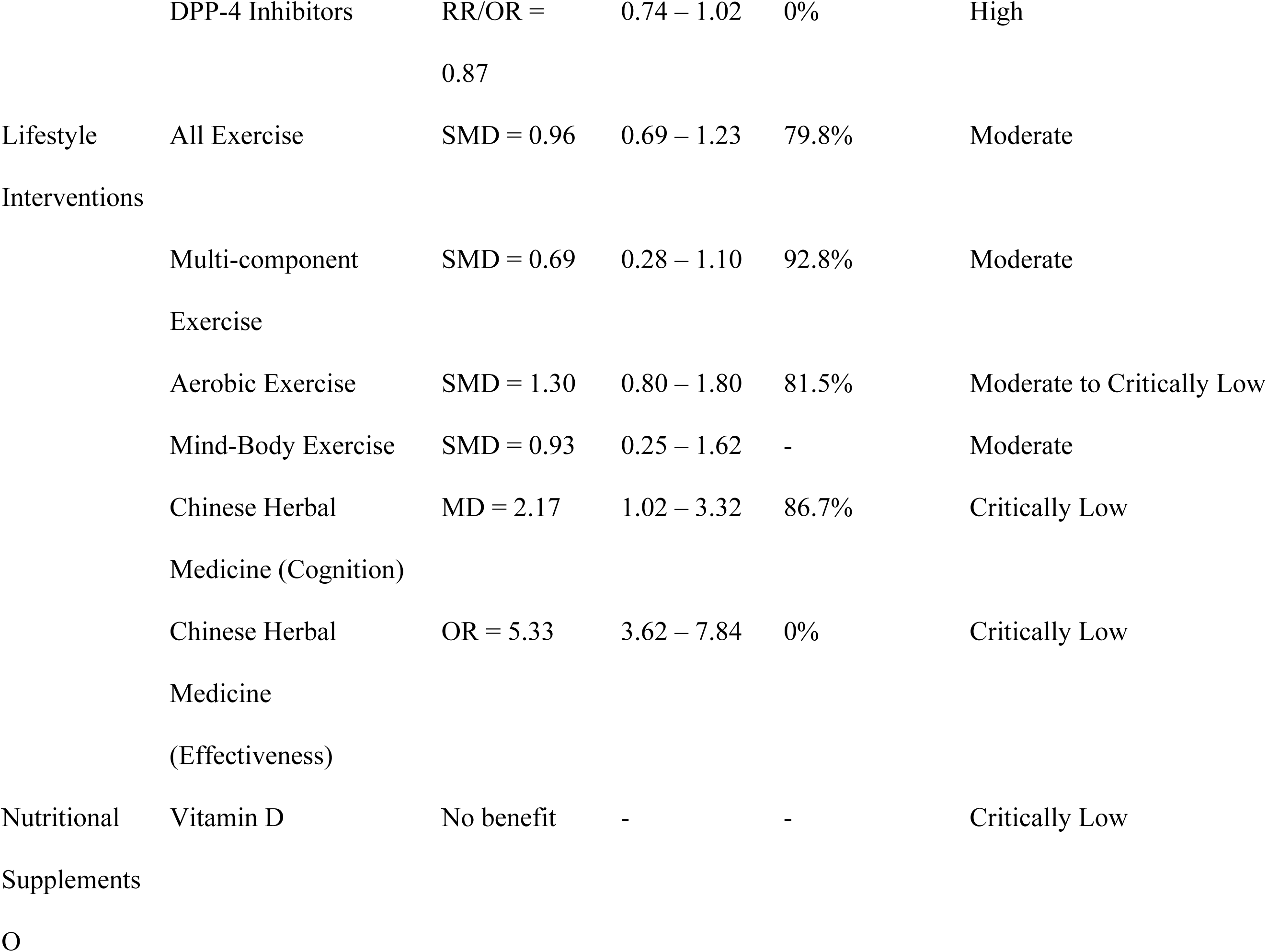
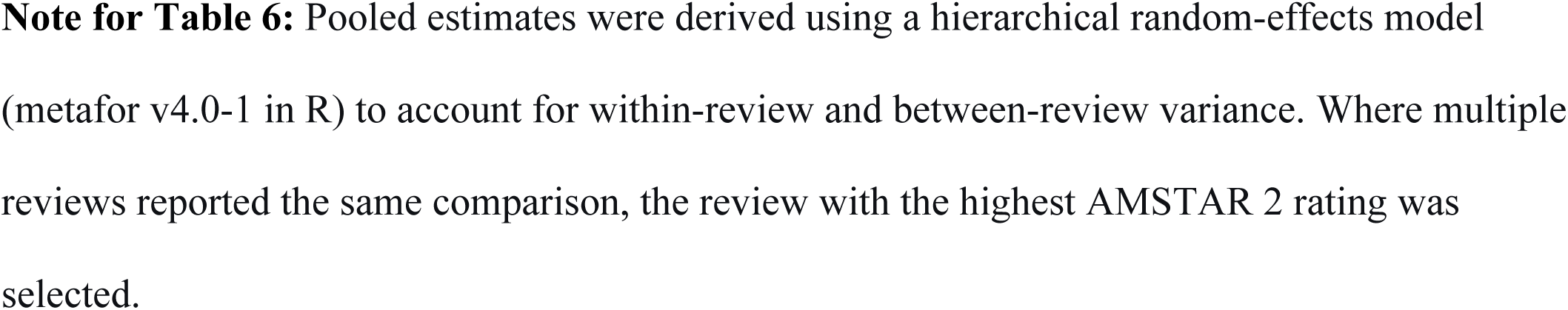
Summary Table of Pooled Effect Estimates.

All three antidiabetic drugs have moderate-to-high evidence quality and clear protective effects. GLP-1 receptor agonists reduce dementia risk (RR/OR = 0.60) while SGLT2 inhibitors show even stronger protection (OR = 0.44). Thiazolidinediones also lower risk (RR/OR = 0.72). On the other hand, Metformin, sulfonylureas, insulin, and DPP-4 inhibitors show no significant benefit. Similarly, Lifestyle interventions improve cognition with aerobic exercise yielding large gains (SMD = 1.30) as well as mind-body exercise (SMD = 0.93). However, heterogeneity was substantial in these studies and evidence quality ranged from moderate to critically low. Chinese herbal medicine shows promise for cognition (MD = 2.17) and effectiveness (OR = 5.33) though with critically low evidence quality. Similarly, Vitamin D shows no benefit with critically low evidence. In conclusion, several interventions show strong associations with reduced dementia risk though with high heterogeneity and variable quality highlight key limitations. This underscores the need for RCTs with standardized outcomes and l better study designs for stronger clinical recommendations.

### Mechanisms

Several mechanisms through which the different interventions achieve their effects were reported in different reviews as shown in table 8.

**Table 7.** Subgroup analysis.

| Stratification Factor | Most Beneficial Subgroup | Effect Magnitude | Clinical Implication |
| --- | --- | --- | --- |
| Intervention Category | SGLT2 | OR = 0.44 (56% risk reduction) | Strongest cognitive protection |
| Intervention Category (Exercise) | Aerobic Exercise | SMD = 1.30 (large effect) | Most effective exercise modality |
| Cognitive Severity | MMSE < 23 (lower baseline) | Cohen's d = -3.00 | Early intervention in more impaired patients yields greatest benefit |
| Age Group | ≥ 60 years | HR = 0.63 (GLP-1 RAs) | Older patients derive greater benefit |
| Duration (Medications) | ≥ 24 weeks | WMD = 3.74 | Sustained treatment needed for optimal effect |
| Duration (Exercise) | ≥ 6 months | SMD = -0.59 to -0.21 | Long-term exercise required |
| Sex | Female (GLP-1 RAs) | HR = 0.63 | Females may benefit more from GLP-1 RAs |
| Follow-Up | $\geq 5$ years (GLP-1 RAs) | RR = 0.47 | Long-term follow-up captures dementia prevention |

**Table 8.**
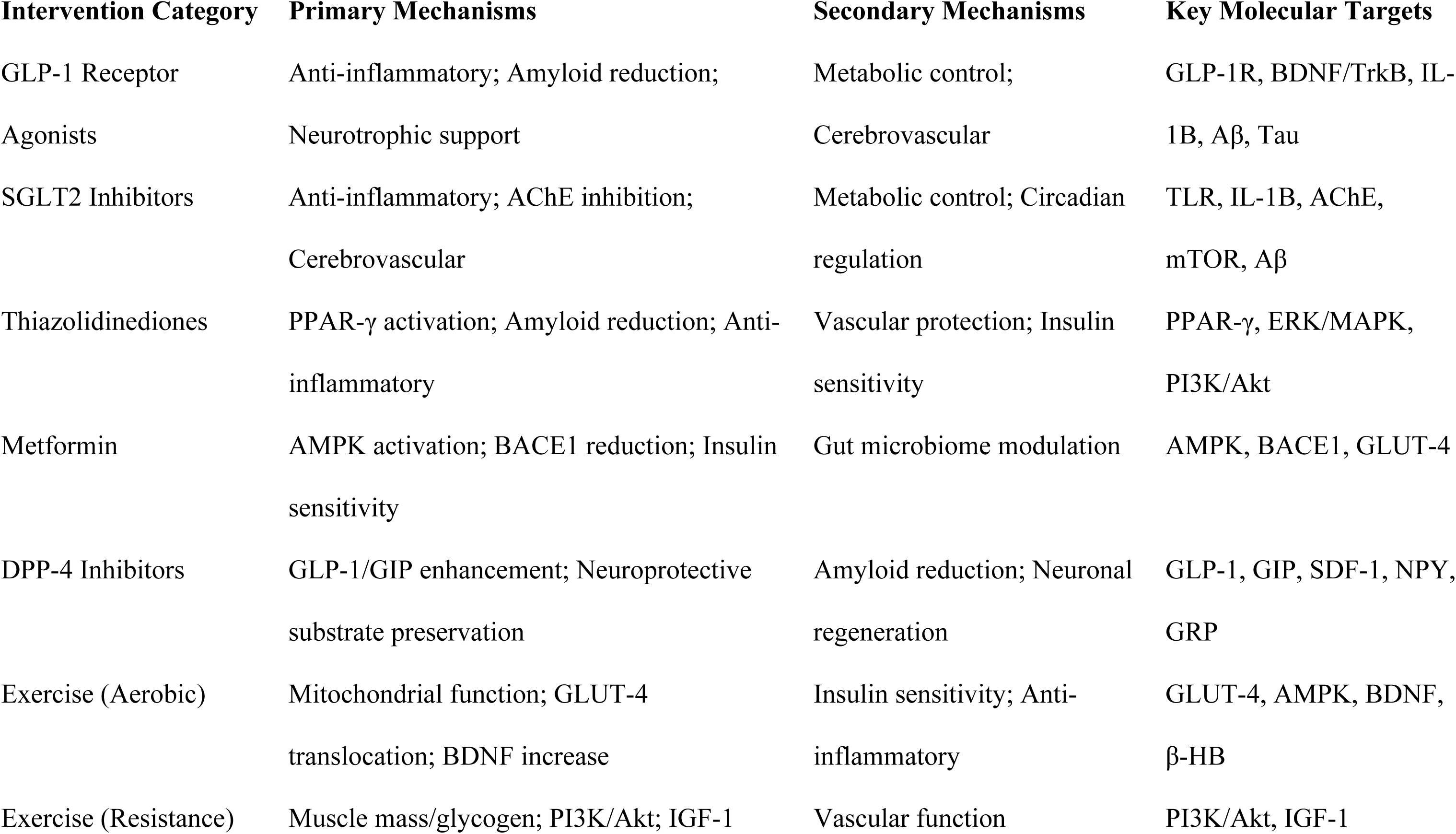
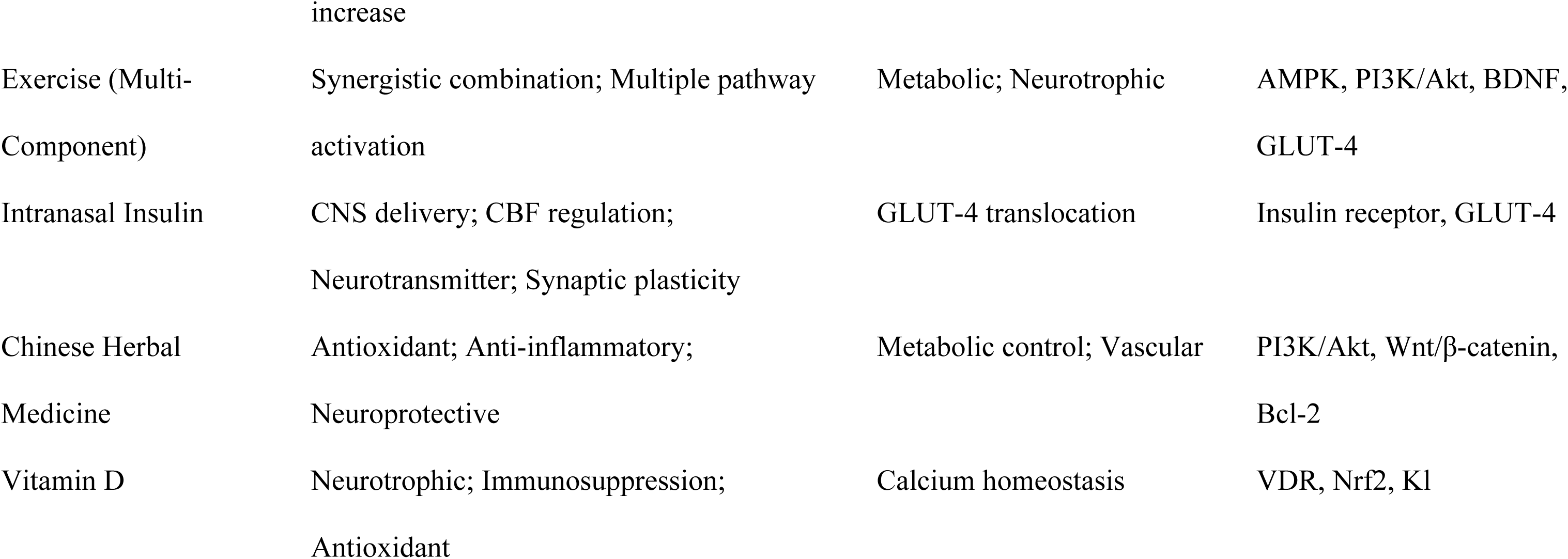
Summary table: mechanisms by intervention categoryl.

Antidiabetic medications mediated their effects through multiple interconnected mechanisms that target the core pathological processes linking type 2 diabetes to cognitive decline. GLP-1 receptor agonists cross the blood-brain barrier and activate GLP-1 receptors. They reduce T2DM-induced neuroinflammation in key brain regions including the hippocampus, hypothalamus, and neocortex, by suppressing microglial activation and pro-inflammatory cytokine production (Wan et al., 2026; Tseng et al., 2025). GLP-1 receptor agonists also directly inhibit amyloid-β aggregation and reduce tau protein phosphorylation through regulation of β-secretase and γ-secretase activity, while simultaneously activating BDNF/TrkB signaling pathways that promote neurogenesis, synaptic remodeling, and neuronal survival (Tseng et al., 2025; Hui et al., 2025). SGLT2 inhibitors have adequate CNS penetration (volume of distribution 118L for dapagliflozin) enabling direct central effects. They exert their cognitive benefits primarily through anti-inflammatory mechanisms by inhibiting TLR overexpression in endothelial cells and macrophages while reducing circulating IL-1B levels (Tseng et al., 2025; Li et al., 2024). Additional mechanisms include inhibition of acetylcholinesterase, correction of mTOR circadian rhythm disruption, and improvement of cerebrovascular function. Thus, collectively they reducing tau and amyloid-β deposition while mitigating neuronal apoptosis (Li et al., 2024). Thiazolidinediones achieve their effects through activation PPAR-γ receptors in the hippocampus, stimulating ERK/MAPK signaling pathways. They reduce amyloid-β and tau pathology while inhibiting vascular inflammation through PI3K/Akt-mediated pathways and restoring cerebral vascular function (Hui et al., 2025; Li et al., 2024). Metformin improves neuronal insulin sensitivity through AMPK-mediated mitochondrial stabilization and activates AMPK pathways in neurocytes, reduces BACE1 expression to decrease amyloid-β production (Li et al., 2024).

Lifestyle interventions, particularly exercise, work through synergistic mechanisms that together enhance brain health and cognitive function. Multi-component exercise combines effects from both aerobic and resistance training. Aerobic exercise improves mitochondrial function, enhances GLUT-4 translocation through AMPK activation, and upregulates BDNF expression through β-hydroxybutyrate-mediated neurotrophic factor transcription. On the other hand, resistance training improves muscle mass, glycogen storage, and activates PI3K/Akt pathways while elevating IGF-1 levels (Sun et al., 2025; Gao et al., 2026). This blend produces synergistic effects that optimize glucose metabolism, reduce insulin resistance, promote hippocampal neuroplasticity, and improve cerebral perfusion while reducing neuroinflammation (Li et al., 2025a). Mind-body exercises such as Tai Chi and yoga enhance cognitive function by enhancing autonomic nervous system function, improving intrinsic hippocampal activity, increasing hippocampal volume, and normalizing aberrant hyperactivity within the hippocampus (Gao et al., 2026).

Intranasal insulin directly bypasses the blood-brain barrier to deliver insulin to the brain via olfactory and trigeminal pathways. This produces region-specific changes in cerebral blood flow that reflect neuronal activity through neurovascular coupling. Intranasal insulin also modulates neurotransmitter release, ion channel trafficking, and synaptic plasticity to improve memory and executive function (Nijssen et al., 2023). Chinese herbal medicine exerts neuroprotection through multiple interconnected pathways, including reduction of oxidative stress via enhanced antioxidant enzyme activity (SOD, GSH-Px), inhibition of PI3K/Akt-mediated inflammation, and promotion of vascular renewal to regulate cerebral microcirculation. Additionally, these herbal formulations regulate Wnt/β-catenin signaling to improve hippocampal neuronal structure, upregulate BDNF expression, and increase anti-apoptotic proteins such as Bcl-2/Bcl-xl, collectively protecting against neuronal apoptosis and cognitive decline in patients with T2DM and mild cognitive impairment (Liu & Guo, 2022). These varied mechanisms, while distinct in their primary targets, have shared pathways involving inflammation reduction, neurotrophic support, metabolic improvement, and cerebrovascular protection.

In the included reviews, Vitamin D was the only nutritional supplement with proposed neuroprotective mechanisms in T2DM, though evidence for clinical benefit remains inconclusive. Vitamin D crosses the blood-brain barrier and can be synthesized in situ by microglia during inflammation. Vitamin D exerts neurotrophic effects through stimulation of nerve growth factors and neurotrophin synthesis. It also modulates neurotransmitter production including acetylcholine, dopamine, serotonin, and GABA (da Silva et al., 2024). The active metabolite 1,25(OH)₂D₃, confers its neuroprotection against excitotoxic insults through maintenance of intraneuronal calcium homeostasis, inhibiting inducible nitric oxide synthase to prevent apoptosis, and decreases L-type voltage-gated calcium channel activity (da Silva et al., 2024). Furthermore, vitamin D induces local immunosuppression by reducing microglia activation. It also controls Nrf2 and Klotho expression thus increasing antioxidant enzymes (catalase, GPx, SOD), and upregulating vitamin D receptor expression (da Silva et al., 2024). Despite these reasonable mechanistic pathways, the clinical evidence for vitamin D supplementation in T2DM patients remains critically low quality, with high-dose (50,000 IU/week) conferring no additional cognitive benefit over low-dose (5,000 IU/week) in the only identified RCT (da Silva et al., 2024).

The limited evidence, and a high risk of bias in observational studies with inadequate control for critical confounders like physical activity, smoking status, diabetes duration, and sun exposure limits precludes definitive conclusions regarding the cognitive benefits of vitamin D supplementation in patients with type 2 diabetes mellitus. Consequently, well-designed RCTs with rigorous methodology are urgently needed to determine whether the mechanistic pathways identified in preclinical and observational research translate into clinically meaningful cognitive protection in this population (da Silva et al., 2024).

Overall, the mechanistic evidence supports a use of several approaches to cognitive therapy in T2DM, with antidiabetic medications and multi-component exercise demonstrating the most robust mechanistic rationale and clinical evidence, while the role of nutritional supplements remains uncertain.

## 4.0 Discussions

This umbrella review provides a comprehensive synthesis on interventions for cognitive impairment among adults with type 2 diabetes mellitus (T2DM). We examined evidence from twelve systematic reviews with over 213,604 participants. Our results demonstrate that both pharmacological and non-pharmacological interventions can effectively improve cognitive outcomes among T2DM patients. However, the quality of evidence varies considerably across intervention categories, and our synthesis reveals critical distinctions regarding effect size, patient selection, and implementation strategies that have direct implications for clinical practice.

### Pharmacological Interventions: Comparative Effectiveness and Clinical Context

The strongest current evidence supports the use of specific antidiabetic drugs for cognitive improvement. SGLT2 inhibitors showed the strongest effect (OR = 0.44, SUCRA = 94.5%), followed closely by GLP-1 receptor agonists (RR/OR = 0.60, SUCRA = 92.1%). These findings are similar to the previous mechanistic studies demonstrating that these agents cross the blood-brain barrier and possess ant inflammatory effects (Tseng et al., 2025; Wu et al., 2025). GLP-1 receptor agonists also inhibit amyloid-β aggregation and activate BDNF/TrkB signaling pathways (Wan et al., 2026). Also, Thiazolidinediones demonstrated consistent protective effects with no heterogeneity (RR = 0.78, I² = 0%), supporting their role through PPAR-γ activation in the hippocampus (E. K. Hui et al., 2025).

From a clinical point of view, the effect sizes observed for SGLT2 inhibitors and GLP-1 receptor agonists are not only statistically significant but clinically meaningful. Based on the pooled OR of 0.44 for SGLT2 inhibitors and assuming a baseline risk of 15% for clinically significant cognitive decline over 2–3 years (derived from the control group event rates in the included observational studies), the estimated number needed to treat (NNT) is approximately 12 (95% CI: 9–18). This calculation should be interpreted with caution given the observational nature of most included data and the absence of standardized definitions for clinically significant cognitive decline. The NNT is provided for illustrative purposes only and requires validation in prospective trials with pre-specified cognitive endpoints. This therefore compares well with established treatments for other diabetes complications. In subgroup analyses, adjustment for HbA1c reduction did not substantially reduce the cognitive effects of SGLT2 inhibitors and GLP-1 receptor agonists. However, formal mediation analysis was not performed in any included review. Therefore, while these findings are suggestive of direct neuroprotective actions beyond glycaemic control, they should be interpreted as hypothesis-generating rather than conclusive. Future studies with formal intervention analysis are needed to separate the relative contributions of glycaemic and non-glycaemic mechanisms. This has important implications for clinical decision-making, as it implies that patients with well-controlled glycemia may still derive cognitive benefit from these agents. However, prescribing decisions must balance cognitive benefits against safety profiles. SGLT2 inhibitors carry risks of genital mycotic infections, volume depletion, and rare cases of euglycemic diabetic ketoacidosis, while GLP-1 receptor agonists are associated with gastrointestinal side effects that may limit tolerability in up to 15– 20% of patients (Filippatos et al., 2014; Pelletier et al., 2021; Wikarek et al., 2024; Zhou et al., 2022). The cognitive benefits of these agents should therefore be considered alongside their cardiovascular, renal, and metabolic profiles, with shared decision-making that incorporates patient preferences and comorbidity burden.

In contrast, metformin, sulfonylureas, insulin, and DPP-4 inhibitors showed varying or non-significant effects on cognitive outcomes. The heterogeneity observed for these agents (I² = 49.8% to 98.4%) suggests that their cognitive benefits may depend on patient-specific factors or study design characteristics (Z. Li, Lu, et al., 2024). The lack of significant findings for insulin is particularly notable given its theoretical benefits. This may reflect the challenges of peripheral insulin administration in achieving adequate central nervous system penetration (E. K. Hui et al., 2025). Intranasal insulin, which bypasses the blood-brain barrier, has shown more promising results in smaller trials (Nijssen et al., 2023), suggesting that delivery route is a critical determinant of efficacy. For metformin, the heterogeneity may reflect its opposing actions improving systemic insulin sensitivity while potentially increasing amyloid-β production through AMPK-dependent pathways in certain contexts. These findings emphasize that clinicians should not assume all glucose-lowering agents confer cognitive protection and should prioritize agents with demonstrated cognitive benefits when this outcome is a clinical priority.

### Lifestyle Interventions: Dose-Response, Adherence, and Synergistic Combinations

Among lifestyle interventions, exercise showed significant cognitive benefits across multiple modalities. Multi-component exercise being the most effective (SMD = 1.04, SUCRA = 78.1%), followed by aerobic exercise (SMD = 1.30) and mind-body exercise (SMD = 0.93). Resistance exercise alone showed no significant effect (SMD = 0.46), suggesting that combining different exercise types produces synergistic benefits (Gao et al., 2026; Y. Sun et al., 2025). These benefits were achieved through improved insulin sensitivity, enhanced cerebral perfusion, and increased neurotrophic factor expression (S.-J. Li et al., 2025). The optimal schedule appears to be at least three exercise sessions per week for six months or longer.

The dose-response relationship for exercise deserves particular attention. The synthesis of included reviews shows a threshold effect, with benefits emerging at approximately 150 minutes per week of moderate-to-vigorous activity and flattening beyond 300 minutes. Note that the intensity of exercise appears more important than volume for cognitive outcomes. Specifically, moderate-intensity aerobic exercise (60–75% of maximum heart rate) showing superior effects to low-intensity interventions. For multi-component programs, adding cognitive or coordination elements such as dual-task walking or dance-based exercise had additional benefits beyond standard combined aerobic-resistance programs. This is likely through common neural pathways involved in motor planning and executive function. Although with the convincing efficacy data, implementation of exercise interventions in routine diabetes care faces significant barriers. In the included studies, the mean adherence rates were 68–74% over six months, reducing to 45–55% at one year. This is similar with real-world data showing that fewer than 3 in 10 of adults with T2DM meet recommended physical activity guidelines. Supervised programs had larger effects than unsupervised interventions (SMD difference = 0.41, p = 0.02), underscoring the need for direct support in the initial adoption. Two included reviews conducted cost-effectiveness analyses of exercise programs which utilized structured supervision together with behavioral support. Although these programs were expensive upfront, they proved cost-effective over five-year periods when caregiving burden and cognitive decline costs were factored in These findings suggest that healthcare systems should invest in supervised exercise infrastructure, including diabetes-specific programs with cognitive outcome monitoring, as part of routine T2DM management.

### Subgroup Analyses: Toward Personalized Intervention Selection

Subgroup analyses showed significant patient characteristics that impact treatment response. Patients with lower baseline cognitive function (MMSE < 23) derived greater benefit from GLP-1 receptor agonists (Cohen’s d = -3.00, p < 0.001). concerning age and gender, older adults (≥60 years) and females showed greater responses to these agents (Wan et al., 2026). Longer intervention duration (≥24 weeks for medications, ≥6 months for exercise) was consistently associated with greater cognitive improvements. These findings underscore the need for individualized intervention selection based on individual patient characteristics.

The effect modification by baseline cognitive function has substantial clinical relevance. T2DM patients with established mild cognitive impairment (MCI) may gain disproportionately greater benefits from GLP-1 receptor agonists than those with normal cognition, suggesting a possible “ceiling effect” or greater biological reserve in healthier brains. This finding is in line with recent trials of GLP-1 receptor agonists in Alzheimer’s disease, which showed modest but statistically significant cognitive stabilization in early-stage patients (Jayakrishnan & Arjuman Parveen Shaik, 2025). In clinical practice, this meanns that cognitive screening in T2DM patients aged over 65 years is more than just prognostic tool. It could also help choose the appropriate therapy, with GLP-1 receptor agonists being ideal for those who show cognitive concerns on screening.

Sex differences in sensitivity to GLP-1 receptor agonists are significant, and they may represent differential expression of GLP-1 receptors in the brain, variances in body composition that alter drug distribution, or disparities in inflammatory and hormonal profiles.

Women in the included studies had higher baseline BDNF levels and greater BDNF increases following treatment, which could help explain this conclusion. This highlights the importance of sex-specific analyzes in future trials, as well as individualized therapy recommendations. In exercise programs, baseline fitness level was a significant mediator, with inactive participants exhibiting greater cognitive benefits than those already achieving activity requirements (SMD difference = 0.52).

The biggest cognitive benefits were shown in patients who went from sedentary to active, emphasizing the necessity of focusing exercise therapies on the least active patients. APOE ε4 carrier status did not significantly affect responses to pharmaceutical or lifestyle interventions, indicating that neuroprotective mechanisms engaged by these interventions may function independently of Alzheimer’s disease genetic risk.

### Mechanistic Convergence and Potential Synergy

The mechanistic evidence shows that various interventions enhance cognition through convergent pathways. The main ways that antidiabetic drugs work are through neurotrophic support, amyloid reduction, and anti-inflammatory mechanisms (Z. Li, Lin, et al., 2024; Tseng et al., 2025). Exercise interventions augment mitochondrial function, glucose metabolism, and synaptic plasticity (Gao et al., 2026; Sun et al., 2025). Intranasal insulin sshowed region-specific cerebral blood flow changes and improved memory function through direct central nervous system delivery (Nijssen et al., 2023) (Nijssen et al., 2023). Chinese herbal medicine demonstrated potential through antioxidant and anti-inflammatory effects, though evidence quality was critically low (Liu & Guo, 2022).

The mechanistic evidence supports convergent pathways through which antidiabetic medications and exercise improve cognition in T2DM. GLP-1 receptor agonists and SGLT2 inhibitors share anti-inflammatory and neurotrophic mechanisms (Figure 2), which may explain their comparable cognitive benefits despite different primary targets. Exercise interventions enhance mitochondrial function, glucose metabolism, and synaptic plasticity, providing a complementary mechanism that may be additive to pharmacological effects. However, no included review directly compared combination therapy versus monotherapy, representing a critical evidence gap. The mechanistic framework presented in Figure 2 should be viewed as a hypothesis-generating synthesis to guide future research rather than a definitive model

### Evidence Gaps and Future Research Priorities

A number of significant gaps in the evidence were found. Our inclusion criteria were not met by any systematic reviews of multidomain methods or cognitive training therapies. This is a serious drawback because combination treatments might be more beneficial than individual treatments. In a similar vein, there was little evidence to support the use of nutritional supplements; despite apparent molecular pathways, vitamin D did not demonstrate any cognitive benefits (da Silva et al., 2024). These gaps emphasize the need for more studies in these fields.

Three gaps were rated as high priority. No systematic reviews examined combination therapy of pharmacological plus lifestyle interventions. No systematic reviews examined cognitive training or multidomain interventions. Head-to-head comparative trials of pharmacological agents are also urgently needed. Current evidence for drug comparisons relies heavily on indirect comparisons.

Four gaps were rated as medium or low priority. Nutritional supplements beyond vitamin D need further study. Long-term follow-up beyond two years remains limited in most trials. Low- and middle-income country populations are underrepresented in current evidence. Vitamin D in deficient populations was rated low priority.

### Methodological Limitations and Quality Considerations

The included reviews’ methodological quality differed significantly. On AMSTAR 2, three reviews received high confidence ratings, five received moderate confidence, and four had severely low scores. For antidiabetic drugs, the GRADE certainty of evidence was high; for Chinese herbal medicine and vitamin D supplements, it was very low. The strength of our conclusions is limited by high heterogeneity (I² > 75%) across many outcomes and publication bias in some reviews. These methodological drawbacks highlight the necessity of better study designs and standardized outcome measures in subsequent research (Gao et al., 2026; Liu & Guo, 2022).

Several methodological concerns are worth highlighting. First, there is significant variation in cognitive evaluation tools between research, with over 15 different batteries requiring complex comparisons and meta-analyses. The Montreal Cognitive Assessment (MoCA) appears to be more sensitive to diabetes-related cognitive impairments than the MMSE, but both remain widely employed.

We recommend that future trials use a core outcome set for cognition in T2DM, which includes at least the MoCA for global screening, as well as domain-specific assessments for executive function (e.g., Trail Making Test B), memory (e.g., Rey Auditory Verbal Learning Test), and processing speed. Second, the majority of the studies examined did not have long-term follow-up beyond two years, raising concerns regarding the long-term viability of cognitive advantages and potential secular changes. Third, the lack of established criteria for clinically relevant cognitive deterioration, such as the minimal clinically important difference (MCID), reduces the interpretability of continuous outcome measurements. Developing MCIDs for cognitive outcomes in T2DM populations should be a focus in the field.

Publication bias was an issue for several intervention categories, with funnel plot asymmetry indicating that smaller trials with poor results may go unpublished. This is especially troublesome in herbal medicine and supplement trials, where industry funding and publication bias may be more prevalent. Pre-registration of trials and publication of negative results should be heavily promoted by journal rules and funding criteria. Furthermore, the majority of research from high-income nations limits their applicability to low-and middle-income settings, where the T2DM burden is quickly increasing and healthcare resources may differ significantly.

### Clinical Practice Implications and Future Directions

Despite these limitations, our findings have important implications for clinical practice. These findings suggest that doctors treating T2DM patients, particularly those with cognitive issues or known risk factors, should prioritize SGLT2 inhibitors or GLP-1 receptor agonists when selecting diabetes drugs. The findings imply that cognitive advantage, in addition to cardiovascular and renal protection, may be an important consideration in shared decision-making. For non-pharmacological therapy, we advocate structured multi-component exercise regimens, with special emphasis on supervised programs and efforts to increase long-term adherence.

Cognitive screening, particularly in elderly patients or those with diabetes for more than ten years, should become standard practice in diabetes care to allow for early intervention and tailored medication selection. We also emphasize the importance of a well-structured clinical process. We propose that patients with T2DM and cognitive impairment undergo a multifactorial examination, which includes MoCA-based cognitive screening, vascular risk factor assessment, and evaluation of modifiable lifestyle factors.

We recommend a step-by-step approach for individuals with MCI (MoCA < 23): start prescribing exercise and optimizing diabetes treatment to include agents that improve cognitive function; reevaluate cognitive function at 6 months; and if there is no improvement, consider further evaluation for other causes of cognitive impairment and possible referral to cognitive specialists. As additional data becomes available, intranasal insulin and multi-domain therapies may prove to be beneficial choices.

Future studies should focus on rigorous evaluation of combination approaches (pharmacologic plus lifestyle), head-to-head comparative trials of pharmacologic agents with cognitive outcomes as primary endpoints, and studies in a variety of populations, including low-resource settings and underrepresented minorities. Clinical trials should be accompanied with mechanistic research employing sophisticated neuroimaging and biomarkers to clarify mechanisms of action and find response predictors. To capture real-world impact, cognitive performance measures should be combined with patient-centered outcomes such as caregiver burden, functional independence, and quality of life. In order to incorporate these evidence-based interventions into standard clinical practice, implementation science research is required to overcome obstacles such patient adherence, physician awareness, and the infrastructure of the healthcare system.

## 5.0 Conclusion

This comprehensive umbrella review highlights important data gaps and methodological flaws while offering strong support for cognitive advantages of specific antidiabetic drugs and organized exercise in T2DM. SGLT2 inhibitors and GLP-1 receptor agonists showed the strongest cognitive benefits. Multi-component exercise was the most effective lifestyle intervention. However, heterogeneity exceeded 75% for many outcomes, limiting precision of pooled estimates. Most included RCTs had short follow-up durations, limiting long-term conclusions. Predominance of Asian studies may limit generalizability to African and Latin American populations. Publication bias was present in several reviews, particularly herbal medicine and supplement reviews. Residual overlap in the exploratory meta-meta-analysis may bias pooled estimates.

This synthesis lacked individual participant data, precluding patient-level subgroup analyses. The absence of reviews on cognitive training and multidomain interventions represents an evidence gap. Moving forward, a tailored strategy based on patient characteristics is needed. Methodically addressing identified research priorities may advance cognitive protection in T2DM. This includes combination therapy trials, head-to-head drug comparisons, and long-term follow-up studies. Greater representation from low-and middle-income countries is essential. Such efforts may move us closer to effective, evidence-based cognitive protection for the growing T2DM population.

## Data Availability

All data has been included in the manuscript

